# Understanding pediatric long COVID using a tree-based scan statistic approach: An EHR-based cohort study from the RECOVER Program

**DOI:** 10.1101/2022.12.08.22283158

**Authors:** Vitaly Lorman, Suchitra Rao, Ravi Jhaveri, Abigail Case, Asuncion Mejias, Nathan M. Pajor, Payal Patel, Deepika Thacker, Seuli Bose-Brill, Jason Block, Patrick C. Hanley, Priya Prahalad, Yong Chen, Christopher B. Forrest, L. Charles Bailey, Grace M. Lee, Hanieh Razzaghi

## Abstract

**Objectives:** Post-acute sequalae of SARS-CoV-2 infection (PASC) is not well defined in pediatrics given its heterogeneity of presentation and severity in this population. The aim of this study is to use novel methods that rely on data mining approaches rather than clinical experience to detect conditions and symptoms associated with pediatric PASC.

**Materials and Methods:** We used a propensity-matched cohort design comparing children identified using the new PASC ICD10CM diagnosis code (U09.9) (N=1309) to children with (N=6545) and without (N=6545) SARS-CoV-2 infection. We used a tree-based scan statistic to identify potential condition clusters co-occurring more frequently in cases than controls.

**Results:** We found significant enrichment among children with PASC in cardiac, respiratory, neurologic, psychological, endocrine, gastrointestinal, and musculoskeletal systems, the most significant related to circulatory and respiratory such as dyspnea, difficulty breathing, and fatigue and malaise.

**Discussion:** Our study addresses methodological limitations of prior studies that rely on pre-specified clusters of potential PASC-associated diagnoses driven by clinician experience. Future studies are needed to identify patterns of diagnoses and their associations to derive clinical phenotypes.

**Conclusion:** We identified multiple conditions and body systems associated with pediatric PASC. Because we rely on a data-driven approach, several new or under-reported conditions and symptoms were detected that warrant further investigation.

**LAY SUMMARY:** Pediatric long COVID in children does not currently have a precise clinical definition, in part due to its widely varying presentation in kids. By comparing children diagnosed with long COVID to children who had COVID-19 but were not diagnosed with long COVID, this study identified several groups of symptoms and conditions that are associated with pediatric long COVID. These findings can be used towards developing a precise definition of long COVID in children for use in future studies.

## BACKGROUND AND SIGNIFICANCE

The National Institutions of Health (NIH) launched the new RECOVER initiative in 2021 (1) to leverage electronic health record (EHR) data to better identify and characterize patients with post-acute sequelae of SARS-CoV-2 infection (PASC), defined by the NIH as failure to recover from COVID-19, or those persistently symptomatic for >30 days.(2) In the adult literature, advances have been made to predict PASC among COVID-19 affected patients (3) and to describe etiology, risk factors, and outcomes (4–7). In contrast, there is currently a paucity of rigorous studies that accurately describe PASC in children (8). Research attempting to better elucidate PASC in children has been limited by small sample sizes (9–11), lack of a control group (12–14), or restrictions in source population (15) that limit generalizability to a broader cohort. (16)

Access to EHR data in a large population of children offers an opportunity to better understand the spectrum of PASC across a wide range of demographics and clinical trajectories. A recent exploratory analysis using EHR data to characterize pediatric PASC examined symptoms, diagnoses, and medications occurring more frequently in a large cohort of SARS-CoV-2 viral test-positive patients when compared with SARS-CoV-2-negative controls. Similar studies have also been carried out in adult populations(7). However, these studies are limited in two important ways. First, SARS-CoV-2 positive patients are heterogeneous and therefore analyses are at risk for spurious associations attributable to residual confounding rather than significant clinical findings. Second and relatedly, studies of PASC-associated features employ outcomes defined by clusters of clinically similar diagnosis codes. Clustering clinical codes in this way may bias findings that confirm clinical experience. The rapidly evolving nature of the pandemic and the lack of consensus of specific symptoms that define PASC in children necessitates a more data-driven approach for knowledge discovery.(17)

In this study, we explored syndromic and systemic features associated with a clinical diagnosis of PASC compared to children with and without SARS-CoV-2 infection. The diagnosis code for PASC was established in October 2021 — *U09.9, post COVID-19 condition, unspecified.* Prior to this, the non-specific code, *B94.8, Sequelae of other specified infectious and parasitic diseases,* was proposed as a temporary alternative (18). While these codes reflect clinician judgment in diagnosing patients who may suffer from PASC, they are likely to have a higher positive predictive value than identifying cases using COVID-19-alone. To identify clusters of PASC-associated diagnoses from tens of thousands of diagnosis codes, we used a tree-based scan statistic, a data mining tool which detects signals from hierarchical structures without relying on pre-specifying the clusters of codes of interest. This data-driven technique is especially advantageous in EHR research where mining a large corpus of data is feasible.

## MATERIALS AND METHODS

### Study Population and Design

The RECOVER PEDSnet EHR population includes EHR data from nine children’s hospitals: Children’s Hospital of Philadelphia, Cincinnati Children’s Hospital Medical Center, Children’s Hospital of Colorado, Ann & Robert H. Lurie Children’s Hospital of Chicago, Nationwide Children’s Hospital, Nemours Children’s Health System (in Delaware and Florida), Seattle Children’s Hospital, and Stanford Children’s Health. The Children’s Hospital of Philadelphia’s institutional review board designated this study as not human subjects research and the need for consent was waived. The PEDSnet COVID-19 Database Version week 141 was used, which includes clinical histories through December 1, 2022 of patients who have been tested for SARS-CoV-2, diagnosed with COVID-19, or received a COVID-19 vaccine^1^. This study is part of the NIH Researching COVID to Enhance Recovery (RECOVER) Initiative, which seeks to understand, treat, and prevent the post-acute sequelae of SARS-CoV-2 infection (PASC). For more information on RECOVER, visit https://recovercovid.org/.

Our primary analyses focused on two comparisons of interest. Cases consisted of patients with evidence of PASC; the two comparator cohorts were patients with and without SARS-CoV-2 infection. Evidence for PASC comprised a U09.9 diagnosis code or an interface terminology (IMO) term in the EHR with the following strings: (‘post’ and ‘acute’ and ‘covid’) or (‘complication’ and ‘covid’). As evidence for PASC, we also admitted a B94.8 (Sequelae of other specified infectious and parasitic diseases) diagnosis code; because the B94.8 code is more general, we did not include such diagnoses where the IMO term indicated something other than PASC. We excluded patients who had a MIS-C diagnosis at any point in time based on the presence of the M35.81, U10, and U10.9 ICD10CM diagnostic codes; therefore, we use ‘PASC’ to refer specifically to non-MIS-C variants. Patients were considered *SARS-COV-2 infected* if they had a diagnostic test that was positive for infection (Polymerase Chain Reaction (PCR), antigen, or serology). Serology tests included IgM, IgG anti-N antibodies, IgG anti-S or receptor binding domain (RBD) antibodies, and IgG and IgA undifferentiated antibodies based on criteria from Mejias et al. for positivity due to infection(19). Patients were also considered SARS-COV-2 infected if they had a diagnosis code for COVID-19 in the inpatient or emergency department (ED) setting. We excluded diagnosis codes for COVID-19 in the outpatient setting because of the possibility that the code was assigned as a rule-out diagnosis during a patient evaluation. Further, the observation period in the study aligned with the phase of the pandemic when viral testing was still widely performed in the healthcare setting. Patients were considered *SARS-COV-2 uninfected* if (1) all available diagnostic tests (PCR, Ag, serology) during the study period were negative and (2) the patient did not have any diagnosis codes indicating SARS-COV-2 infection, MIS-C, or PASC in any setting.

Cohort entry date for incident PASC cases was defined as: (1) the date of the first positive PCR or antigen test, or (2) 4 weeks before the date of the first positive serology test, or (3) 4 weeks before the first occurrence of a diagnosis of PASC if no preceding confirmatory test was available. We recognize that imposing a date of infection may misclassify the risk window between initial infection and PASC diagnosis if no testing data is available but chose to be overly inclusive of patients rather than exclude patients with the diagnosis based on unavailability of testing data (only 533 of 1309 patients with PASC [40.7%] had evidence of SARS-CoV-2 infection prior to their earliest PASC diagnosis). The cohort entry date for non-PASC SARS-CoV-2 infected patients was defined based on the date of the earliest SARS-CoV-2 diagnostic test (unless serology positive, then 4 weeks earlier) or earliest COVID-19 diagnosis/encounter. For SARS-CoV-2 uninfected patients, cohort entry dates were chosen as the date of a random negative test. All patients across case and control definitions were < 21 years of age at the date of cohort entry. Further, to ensure that patients had a history of care at their institution, we required that all patients had at least two encounters in the health system in the 18 months prior to cohort entry.

Additionally, we conducted a sensitivity analysis in which we compared SARS-CoV-2 positive to SARS-CoV-2 negative patients during the post-acute period—a description of the methods and results of this analysis are included in the supplementary appendix.

### Matching

We extracted data for our study from the RECOVER pediatric database by identifying any patient who met study inclusion criteria between March 1, 2020, and December 1, 2022. We used a propensity score matched approach to balance covariates between cases and each of the comparison groups to minimize potential confounding (20). Propensity scores were estimated using logistic regression as the probability of a PASC diagnosis conditional on the following covariates: institution, age group (<1, 1-4, 5-11, 12-15, 16-20 years), sex, race/ethnicity (white non-Hispanic (NH), Black NH, Hispanic, Asian NH, multi-racial NH, other NH), and clinical setting of testing. Additionally, we used the Pediatric Medical Complexity Algorithm (PMCA) (21) to define for each body system an index indicating presence and complexity of chronic condition in that body system grouping and used these PMCA indices in our propensity score model. We then matched PASC patients to comparison patients (SARS-CoV-2 infected patients in the first cohort, SARS-COV-2 uninfected patients in the second cohort) using 5:1 nearest neighbor matching, additionally requiring exact matching on both cohort entry month and age group (22). We assessed the balance between the SARS-CoV-2 infected and uninfected patients via absolute standardized mean differences.

### Tree-based scan statistic

The tree-based scan statistic is a data mining tool that is well established in vaccine safety(23) and disease surveillance and characterization(23, 24), making it salient for applications to understand pediatric PASC in its early stages of characterization. This approach simultaneously evaluates outcomes at different levels of granularity in a hierarchical structure, adjusting for multiple testing using a likelihood ratio statistic. In this study, we employed the unconditional analysis based on a Bernoulli probability model at each node of the tree, as described in the TreeScan User’s Guide(25). This approach identifies branches of the tree at which outcomes belonged most disproportionately to cases as compared to controls.

As inputs, we used the ICD10CM vocabulary following the ICD10CM Tabular List of Diseases and Injuries(26) as developed by the National Center for Health Statistics (NCHS). For example, a path down the hierarchy might proceed as follows: C00-D49-> C00-C97 -> C44 -> C44.1 -> C44.10 -> C44.102 -> C44.1021. In all, the hierarchy has 7 levels, with the following counts of nodes per level starting from the top down: 22, 262, 2274, 9502, 14400, 22236, 48450.

We will alternately refer to the hierarchy as a tree, and the cluster consisting of a node together with all of its descendants will be referred to as a *branch* of the tree, or equivalently, as a *cut.* For each branch of the tree, we started with the observed count of cases who had an incident diagnosis in that branch during the outcome period. We also calculated the expected number of cases for each branch by multiplying the proportion of cases in the cohort (1/6 due to 5:1 matching) by the number of controls who had an incident diagnosis in the branch. The null hypothesis, for each cut, is that the number of observed cases equals the number of expected cases. This method adjusts for multiple testing in the sense that under the null hypothesis that the number of observed cases equals the number of expected cases at each cut, there is a 95% probability that there is no cut in the tree with a p-value <0.05. For each cut demonstrating significance at the p<0.05 threshold, we calculated percent excess cases as the difference between number of observed cases and the number of expected cases divided by the number of expected cases. Analyses were carried out using R (4.1.0) and Treescan (2.0) software. Subclassification matching was implemented using the MatchIt R package and tree visualizations were built using the collapsibleTree R package.

### Outcomes

For each patient and each diagnosis code, we calculated a binary indicator for whether the patient had an incident occurrence of that diagnosis code during the 28-179 days following cohort entry. We then used these to compute for each diagnosis code, and consequently each cluster of diagnosis codes defined by a branch of the tree, the number of cases and controls who had an incident diagnosis in that branch.

To define incident outcomes, we employed a washout period spanning from 18 months prior to cohort entry to 7 days prior to cohort entry for conditions grouped at the 3^rd^ level of the ICD hierarchy (I.e., the part of the ICD code before the decimal place) to ensure new incident diagnoses. In other words, conditions which occurred during the study period were not counted if a code in the same group occurred during the washout period.

## RESULTS

### Study population and matching

Between March 1, 2020, and June 22, 2022, there was a total of 14,399 patients identified for inclusion into the 3 cohorts, with PASC cases accounting for 1,309 patients (Table 1). Older children and females were more likely to be included in the PASC cohort than younger children and males (overall: 54.9% vs 45.1%). The most common cohort entry months were in the fall of 2021. A majority of children in the PASC cohort (55.8%) had a chronic condition. Following 5:1 nearest neighbor matching, both comparison cohorts had excellent balance (|SMD|<0.1 for all variables). (Supplementary Appendix Figures 1a-b). After matching, the demographic distributions of both COVID-positive and COVID-negative cohorts reflect those of the PASC cohort.

**Table 1.**
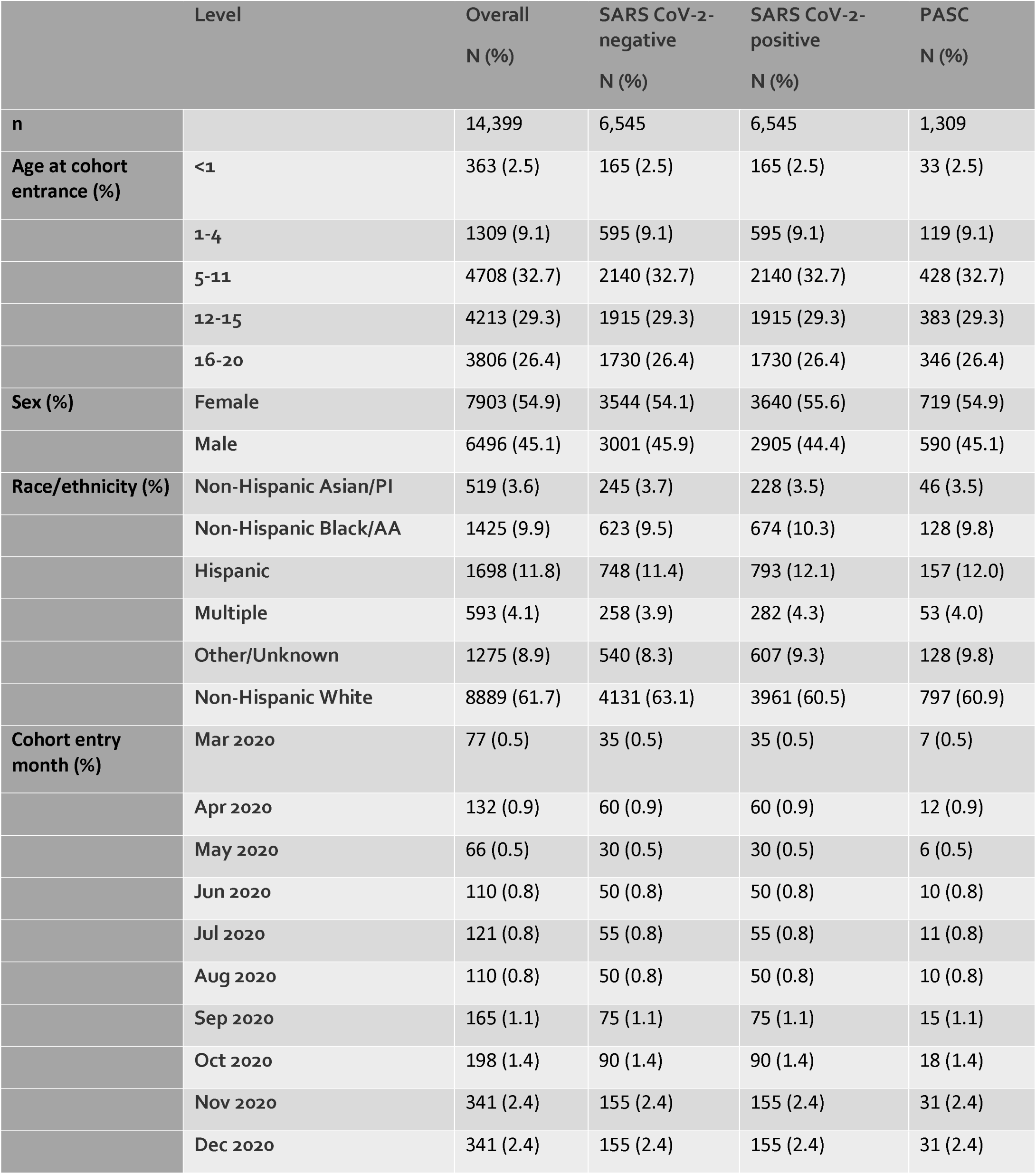
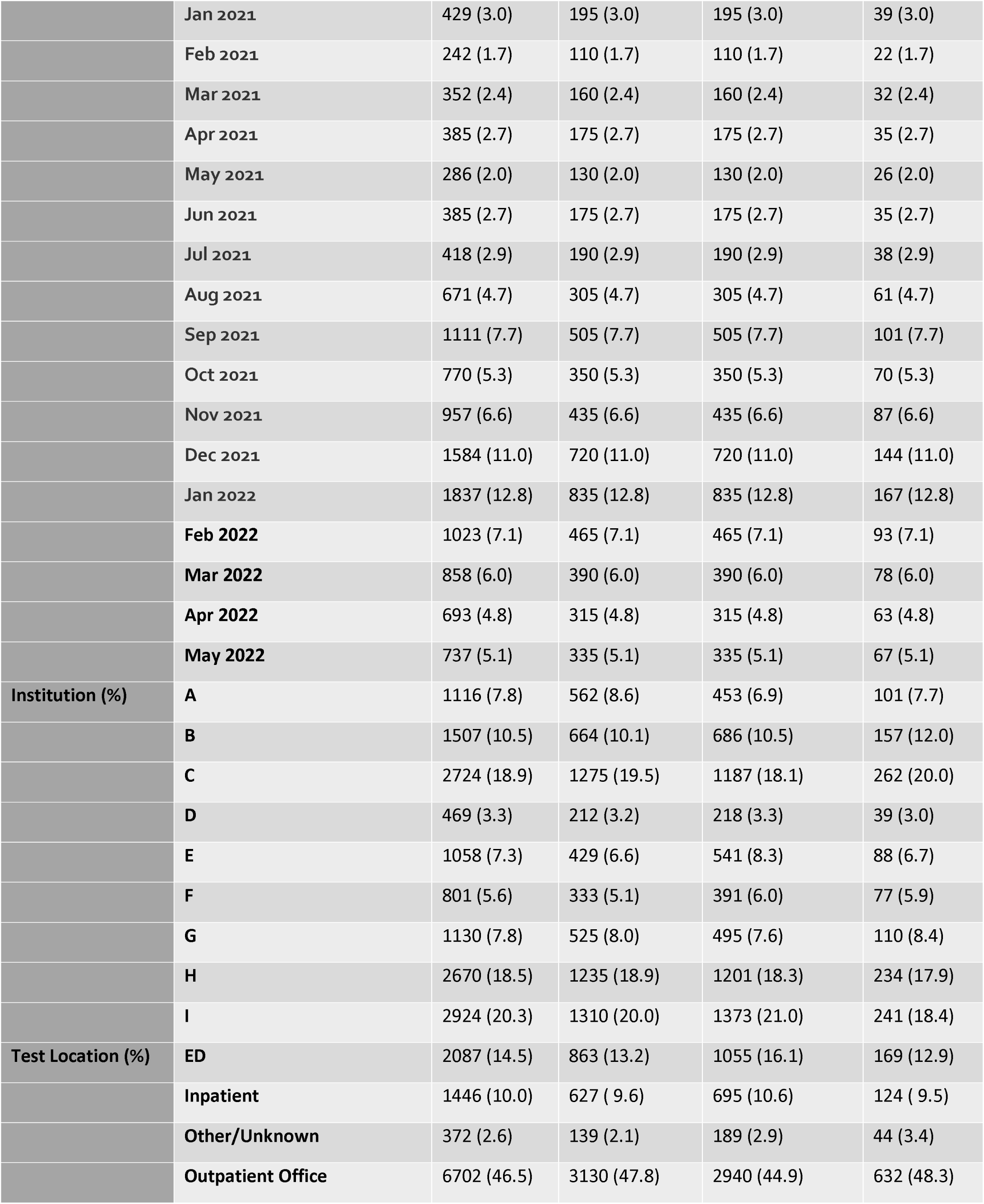
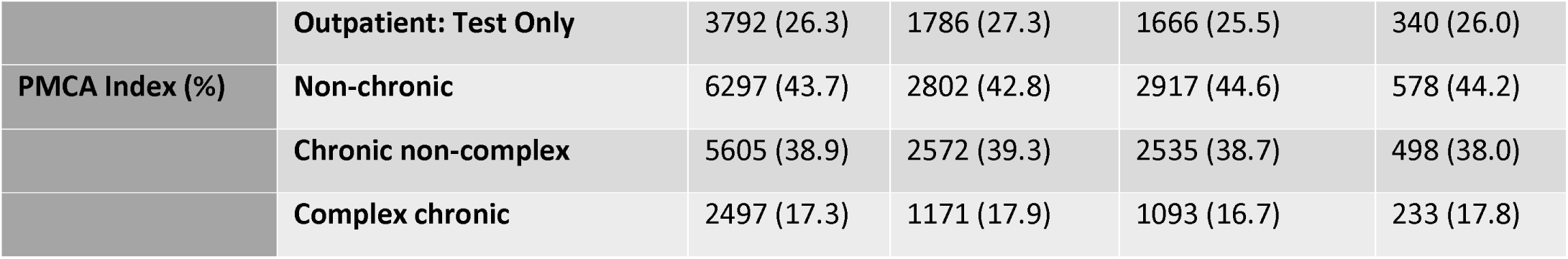
Study population characteristics after matching

### Tree-based scan statistic

When comparing patients with PASC to patients who were SARS-CoV-2 infected, we identified multiple statistical signals. At the highest level of the tree, significant cuts included R00-R99 Symptoms, signs and abnormal clinical and laboratory findings, not elsewhere classified, M00-M99 Diseases of the musculoskeletal system and connective tissue, G00-G99 Diseases of the nervous system, J00-J99 Diseases of the respiratory system, F00-F99 Mental and behavioral disorders, E00-E90 Endocrine, nutritional, and metabolic diseases, I00-I99 Diseases of the circulatory system, Z00-Z99 Factors influencing health status and contact with health services, L00-L99 Diseases of the skin and subcutaneous tissue, and K00-K93 Diseases of the digestive system.

Within the R00-R99 branch, the top three cuts were R00-R09 Symptoms and signs involving the circulatory and respiratory systems, R50-R69 General symptoms and signs and R40-R46 Symptoms and signs involving cognition, perception, emotional state and behavior. Within the R00-R09 branch, the three most significant cuts included R06.0 Dyspnea, R07 Pain in throat and chest, R09.9 Chest pain, unspecified. This subbranch of the R00-R99 branch is visualized in Figure 1.

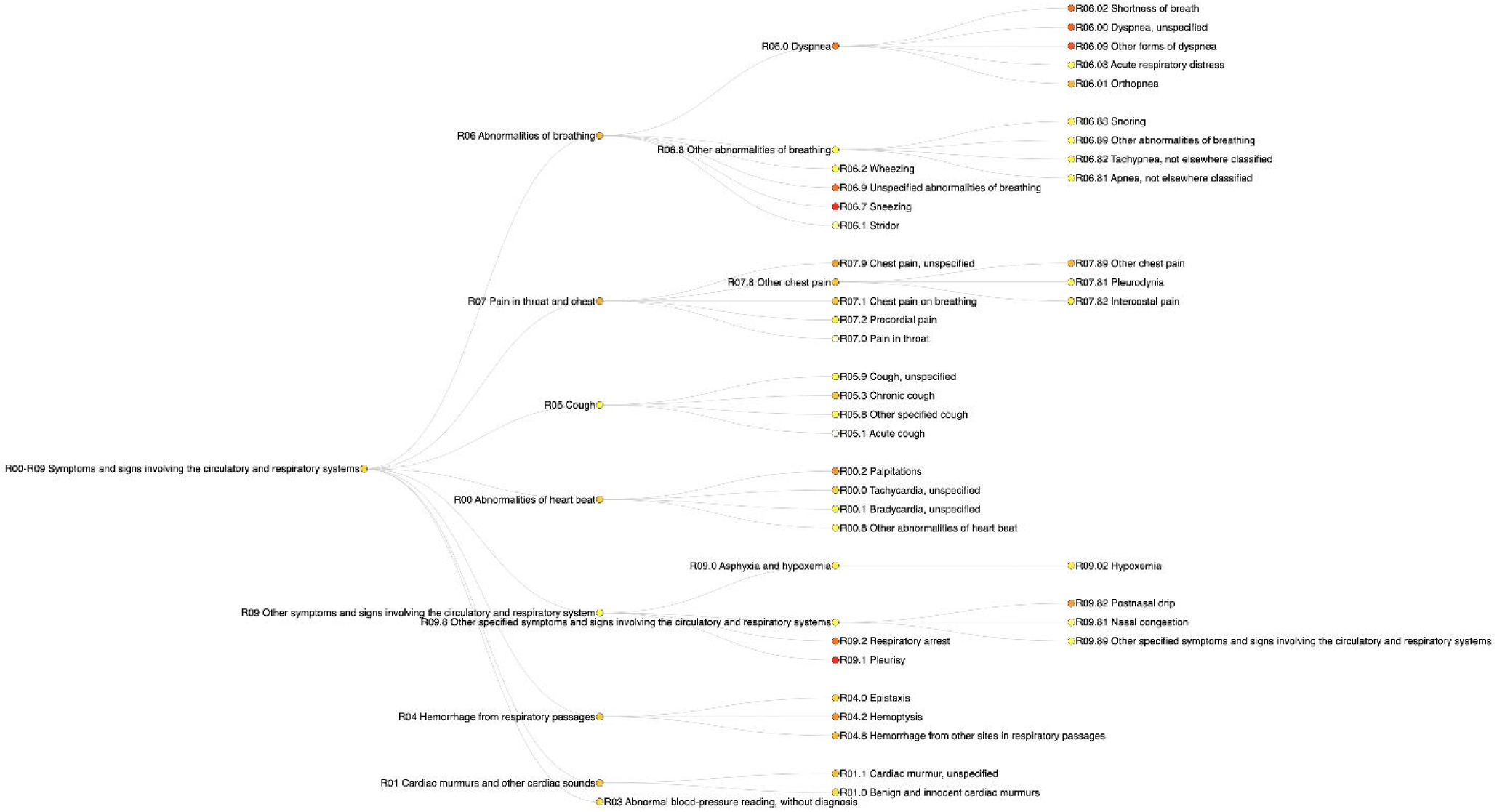

The PASC vs SARS-CoV-2 negative comparison showed significance for 304 cuts, and largely overlapped with the PASC vs SARS-CoV-2 negative comparison at the top of the tree, with the addition of H00-H59 Diseases of the eye and adnexa.

We refer the reader to Table 2 for a summary of systemic and syndromic findings collected from the two comparisons and Tables 3a and 3b for a full list of significant cuts, the likelihood ratio statistics, p-values, and percent excess cases.

**Table 2:**
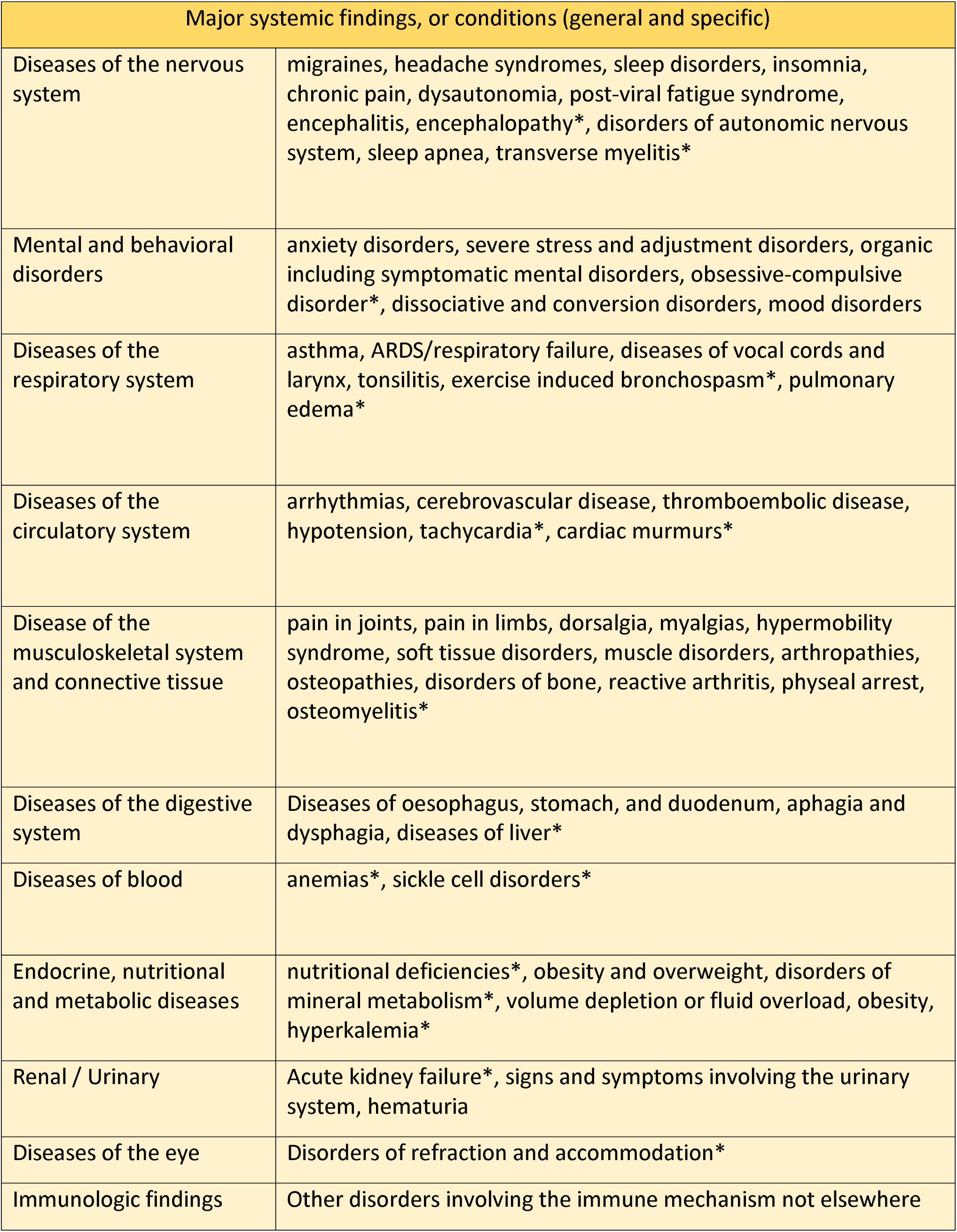
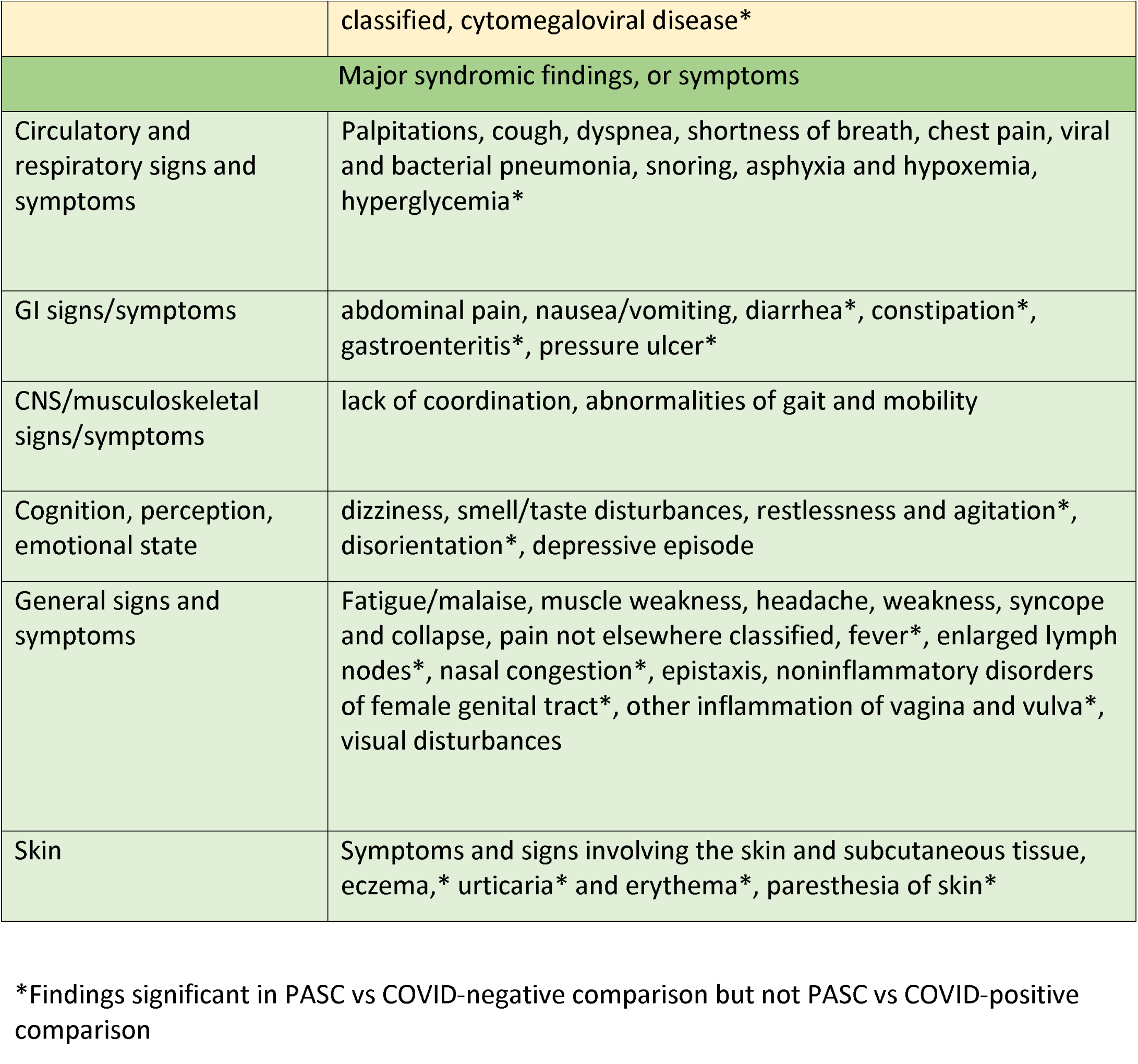
Summary of significant features in the two comparison cohorts

**Table 3a:**
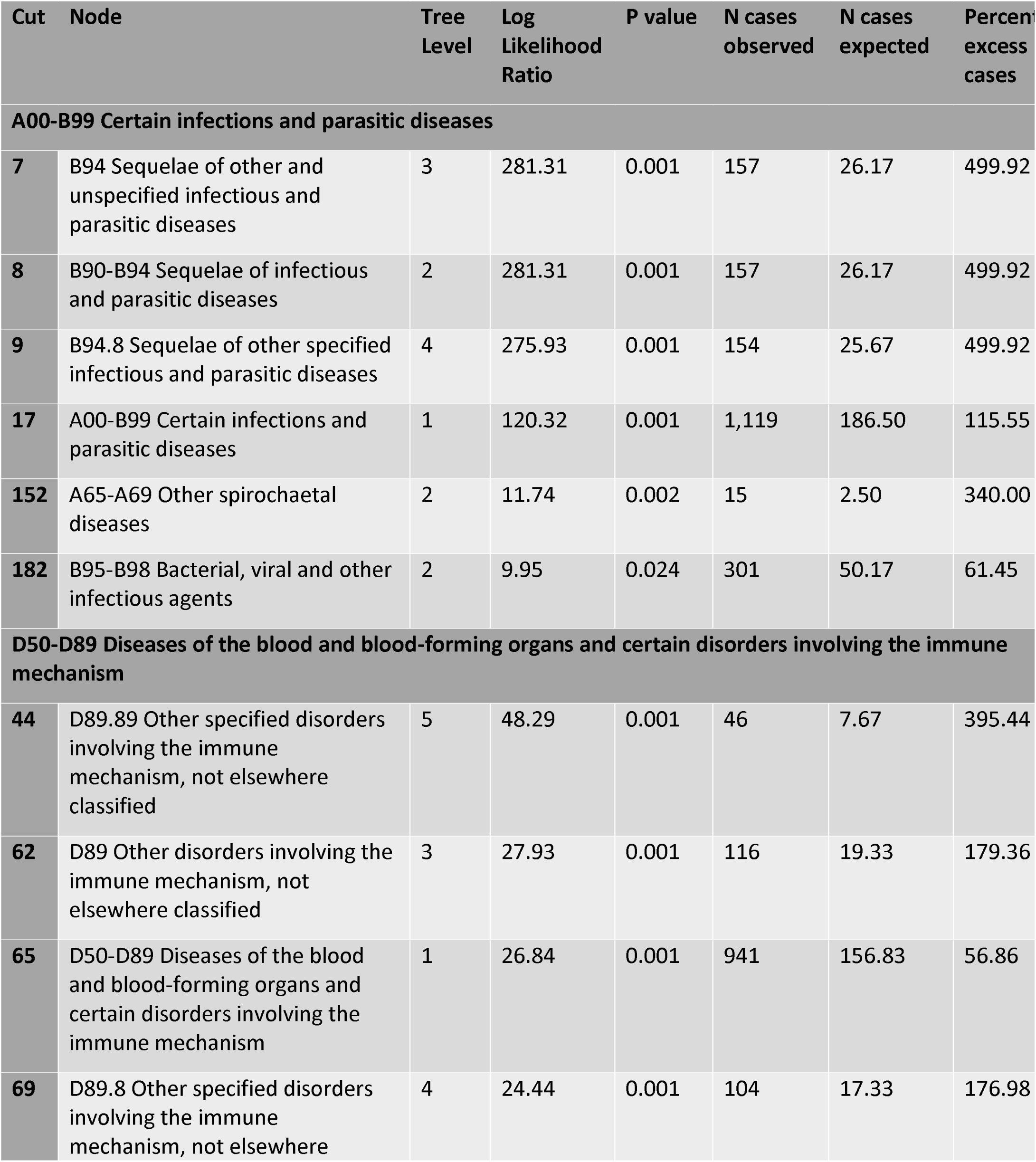
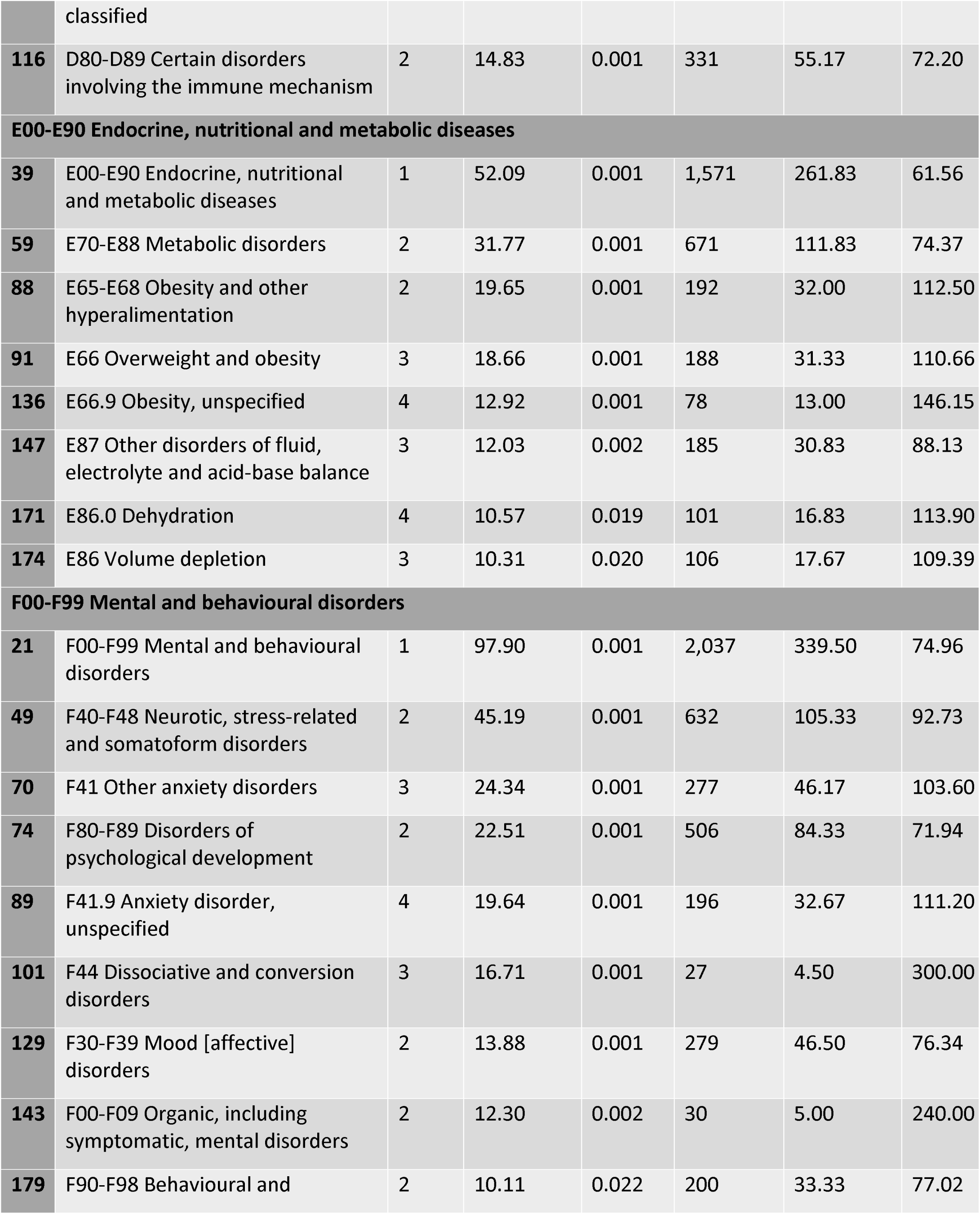
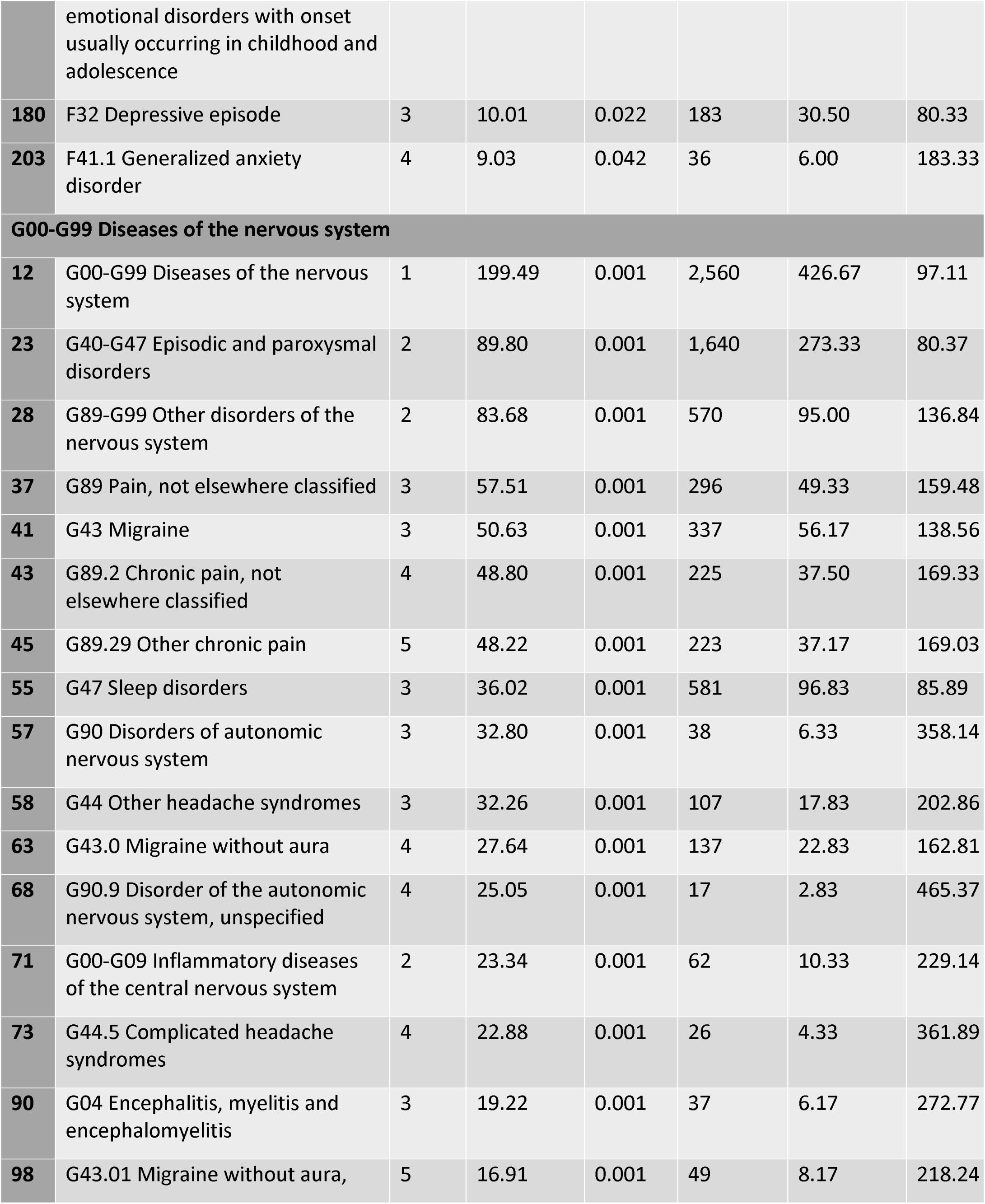
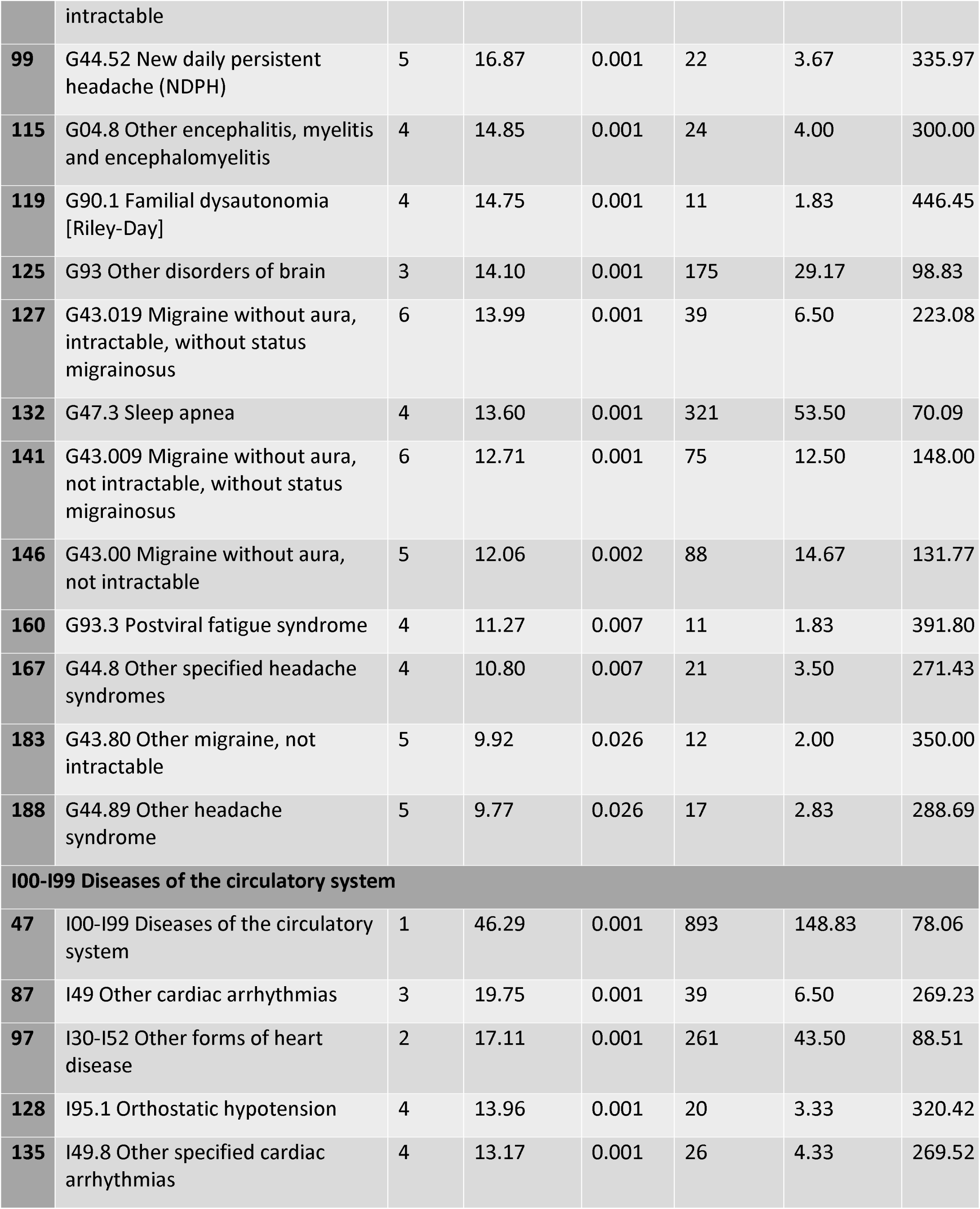
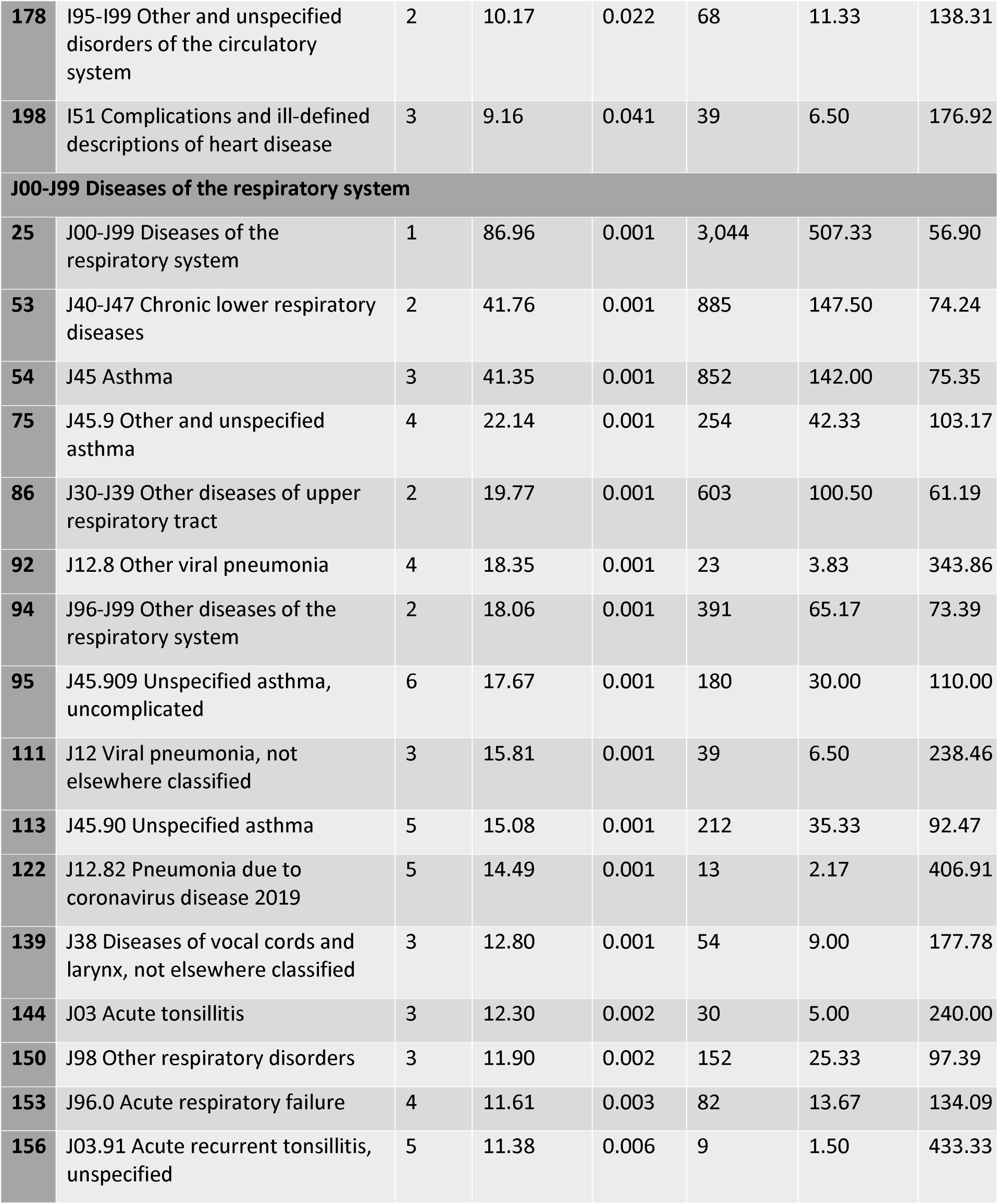
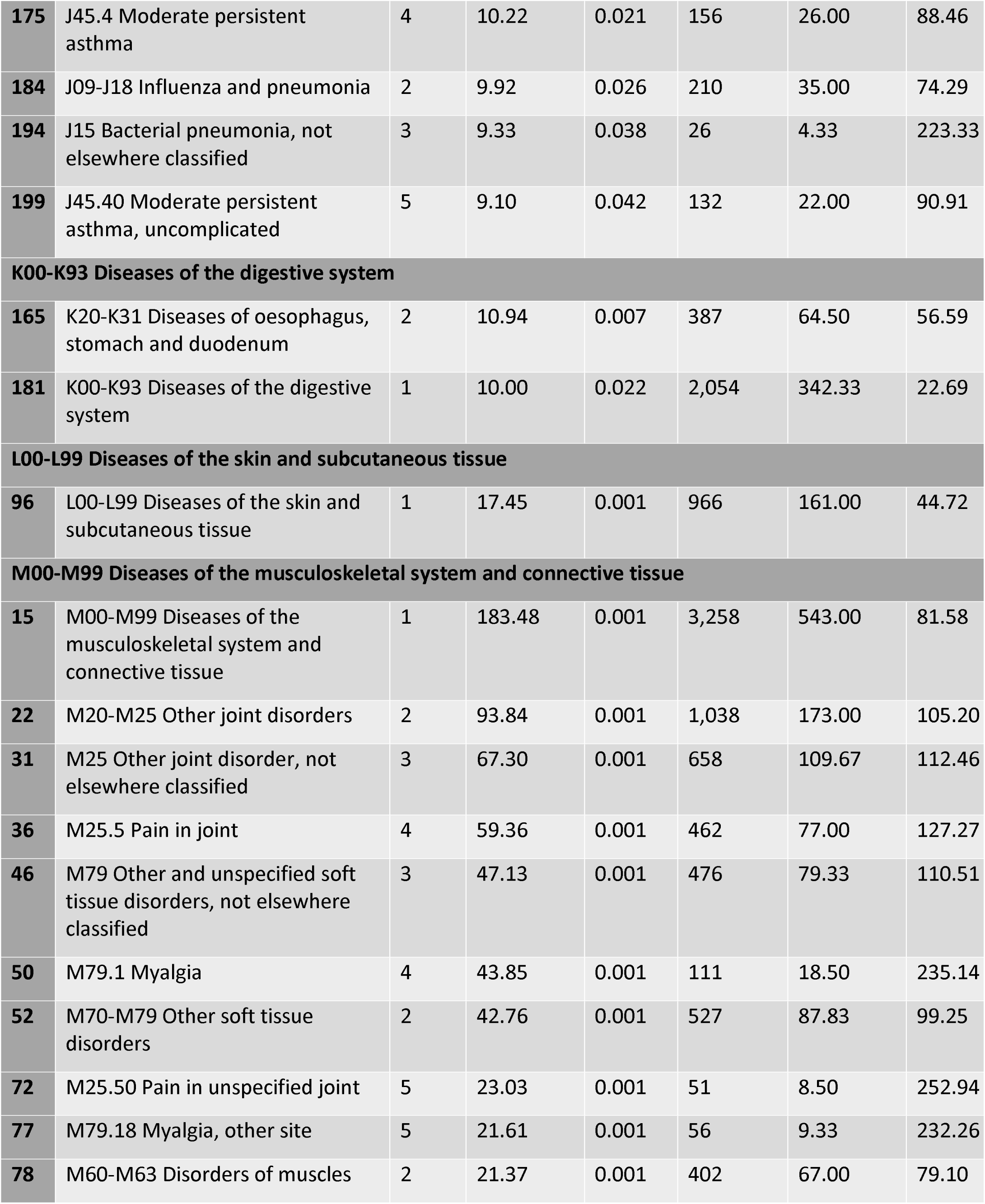
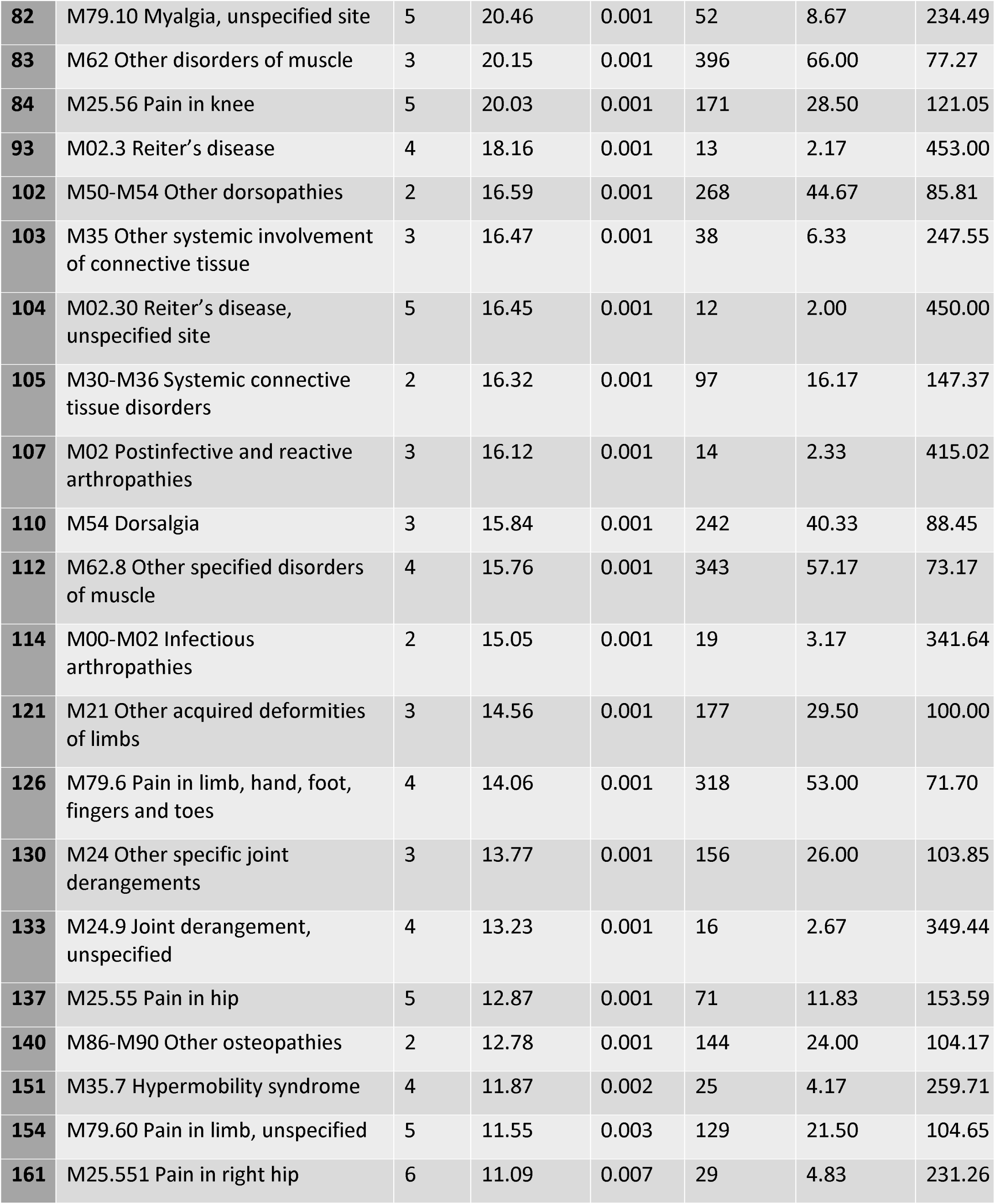
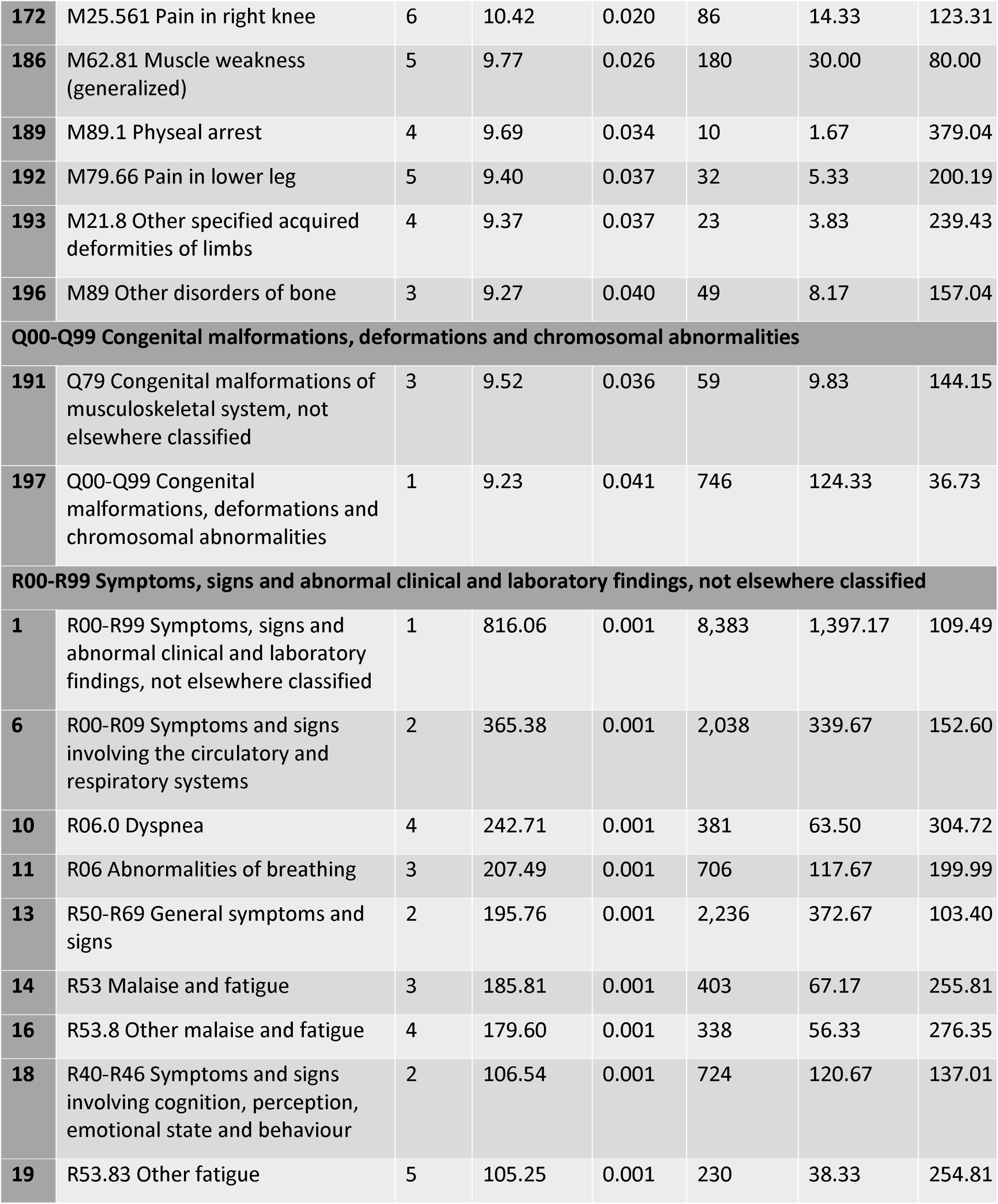
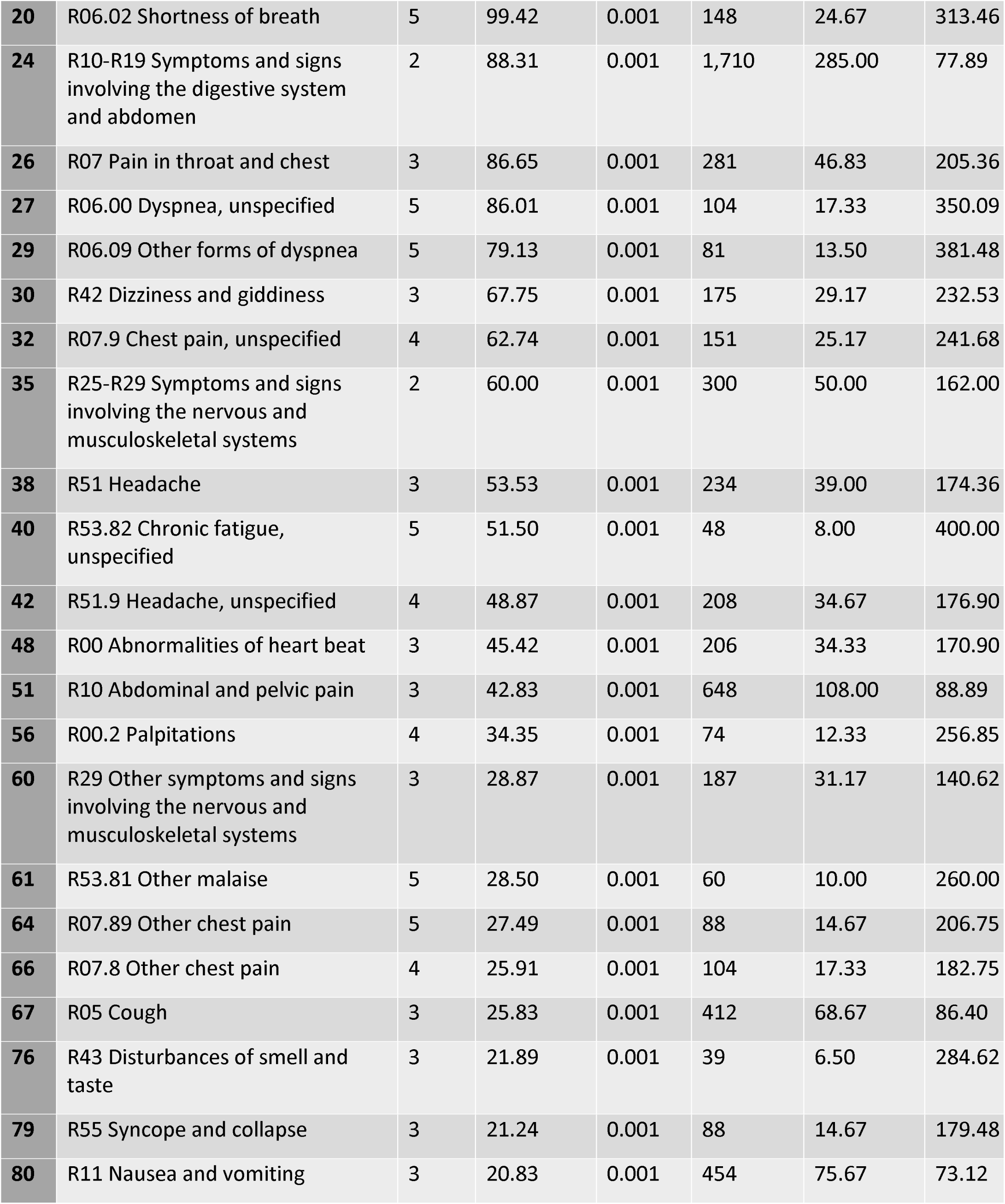
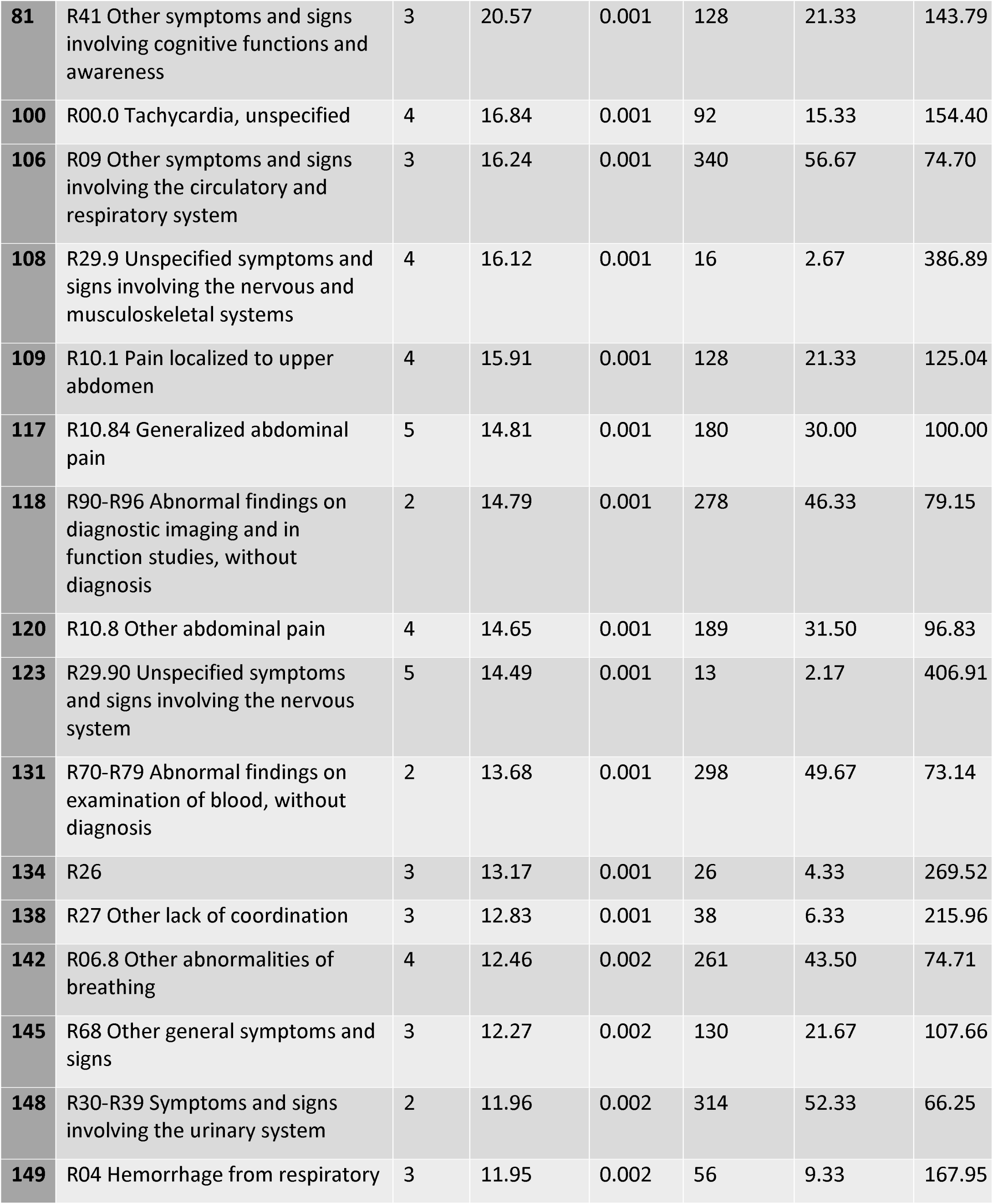
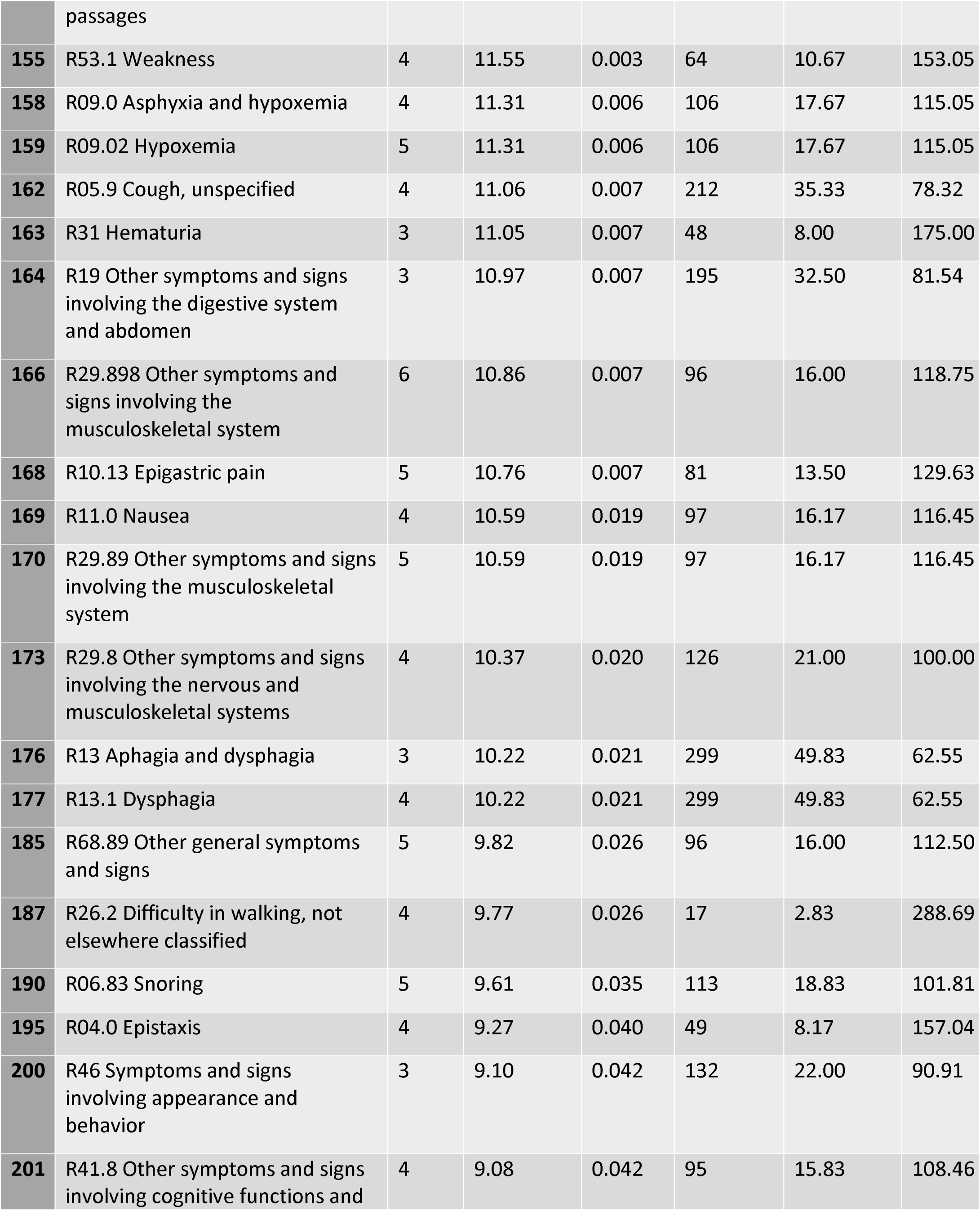
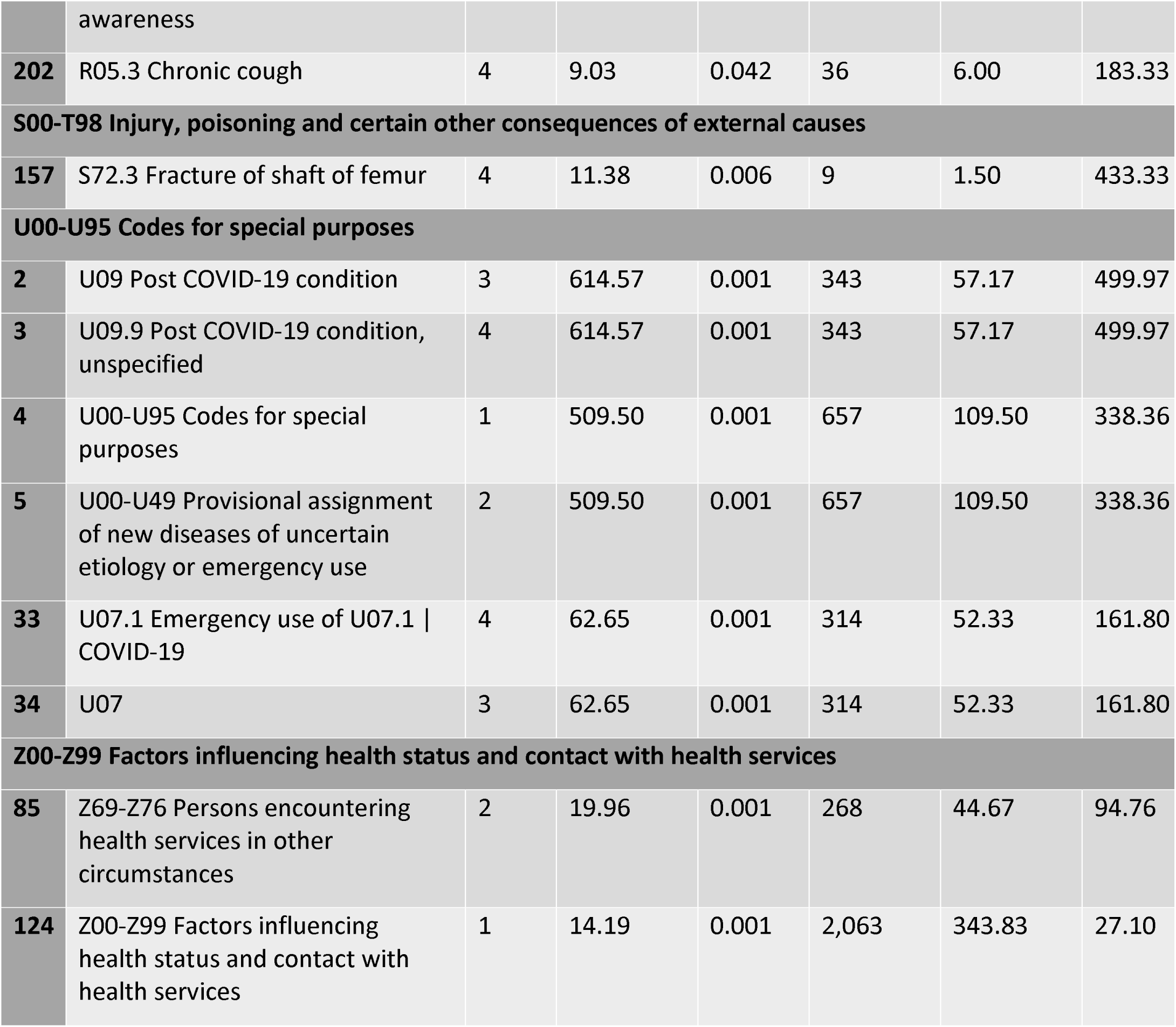
PASC vs COVID positive comparison TreeScan results

**Table 3b:**
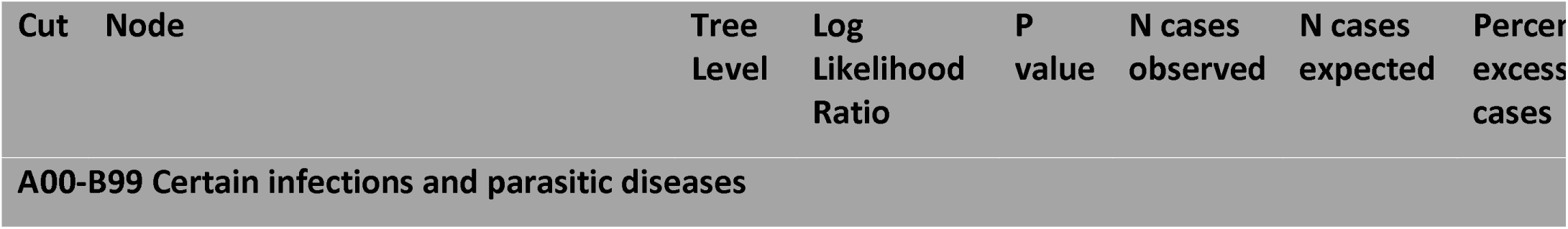
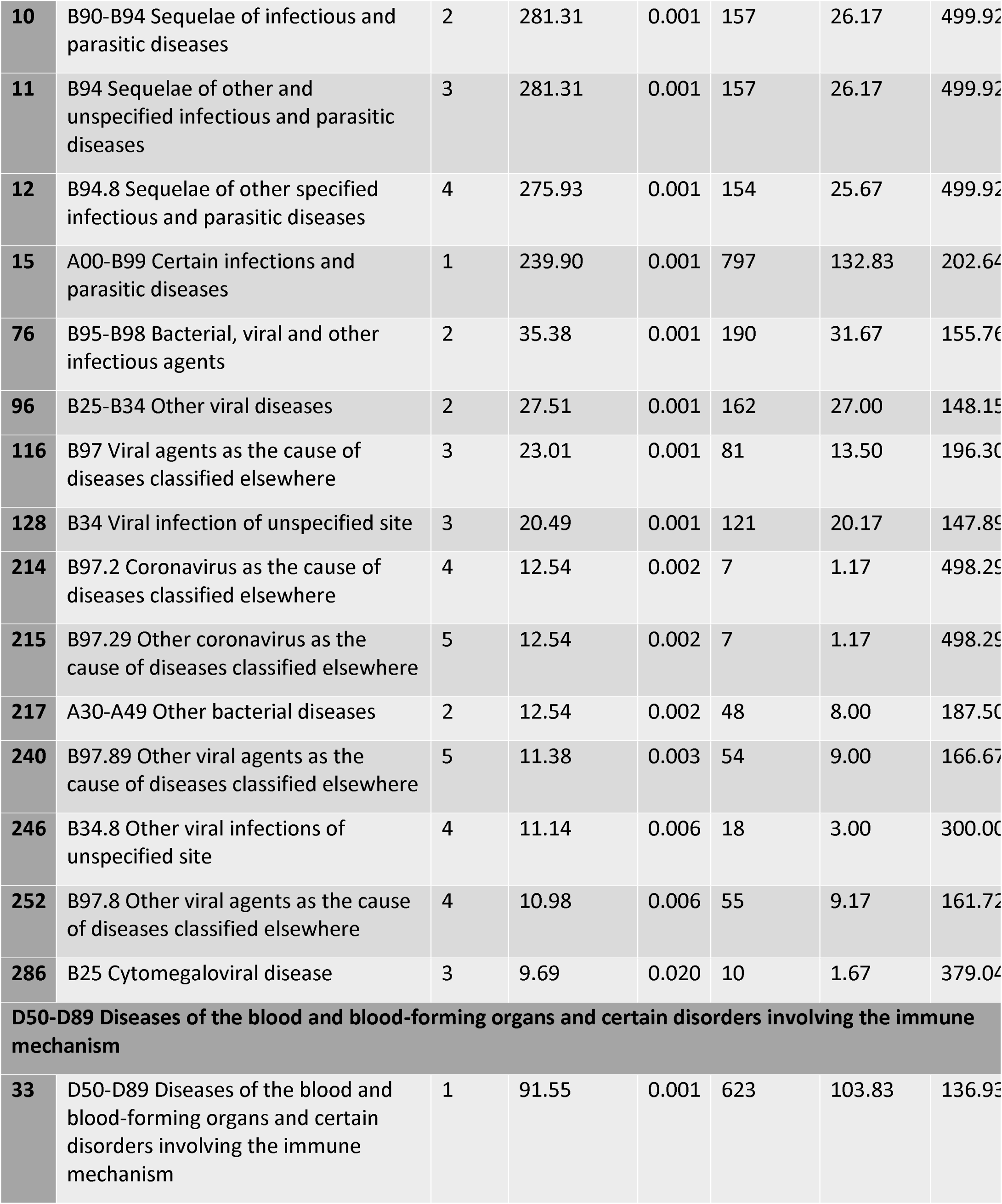
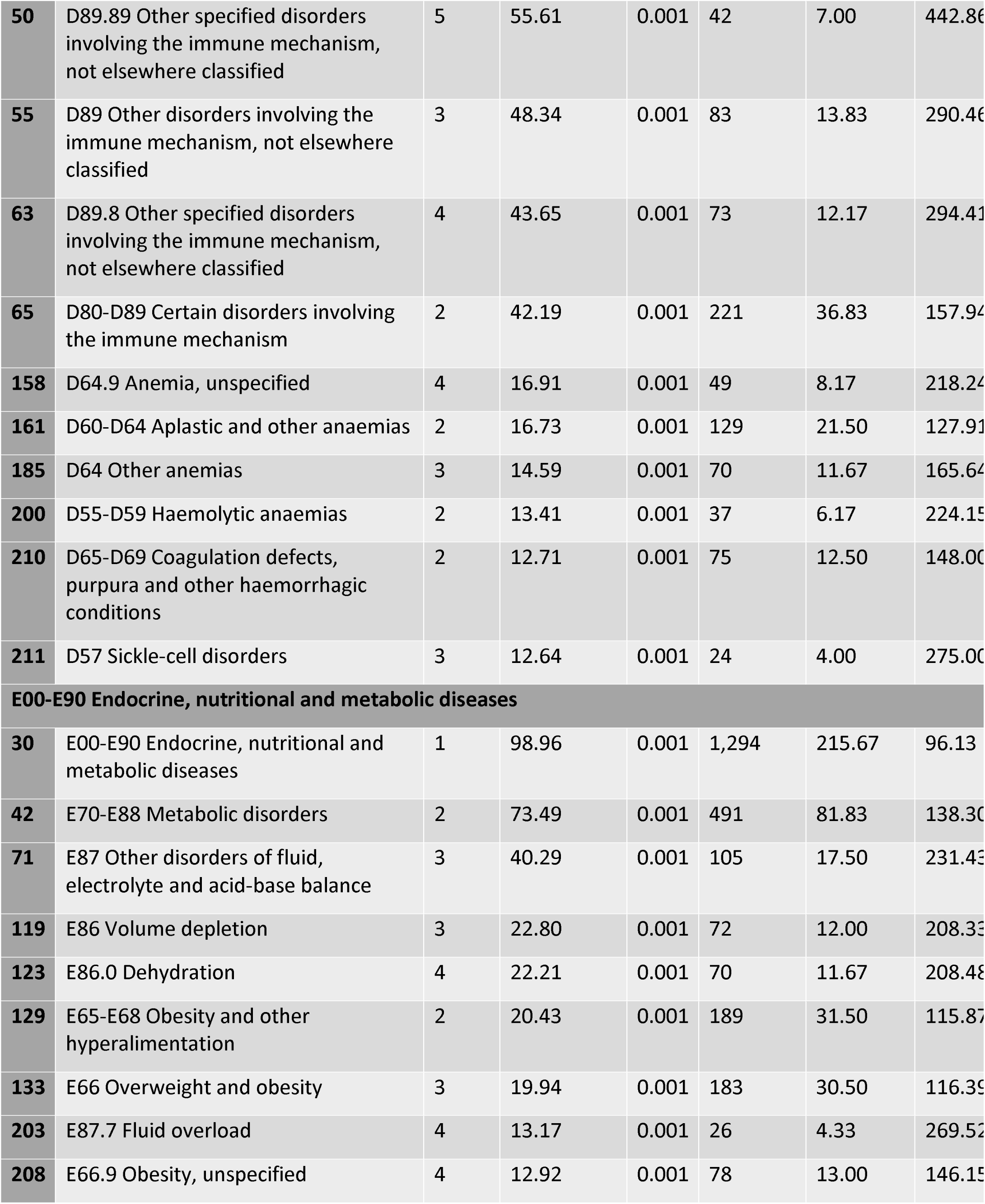
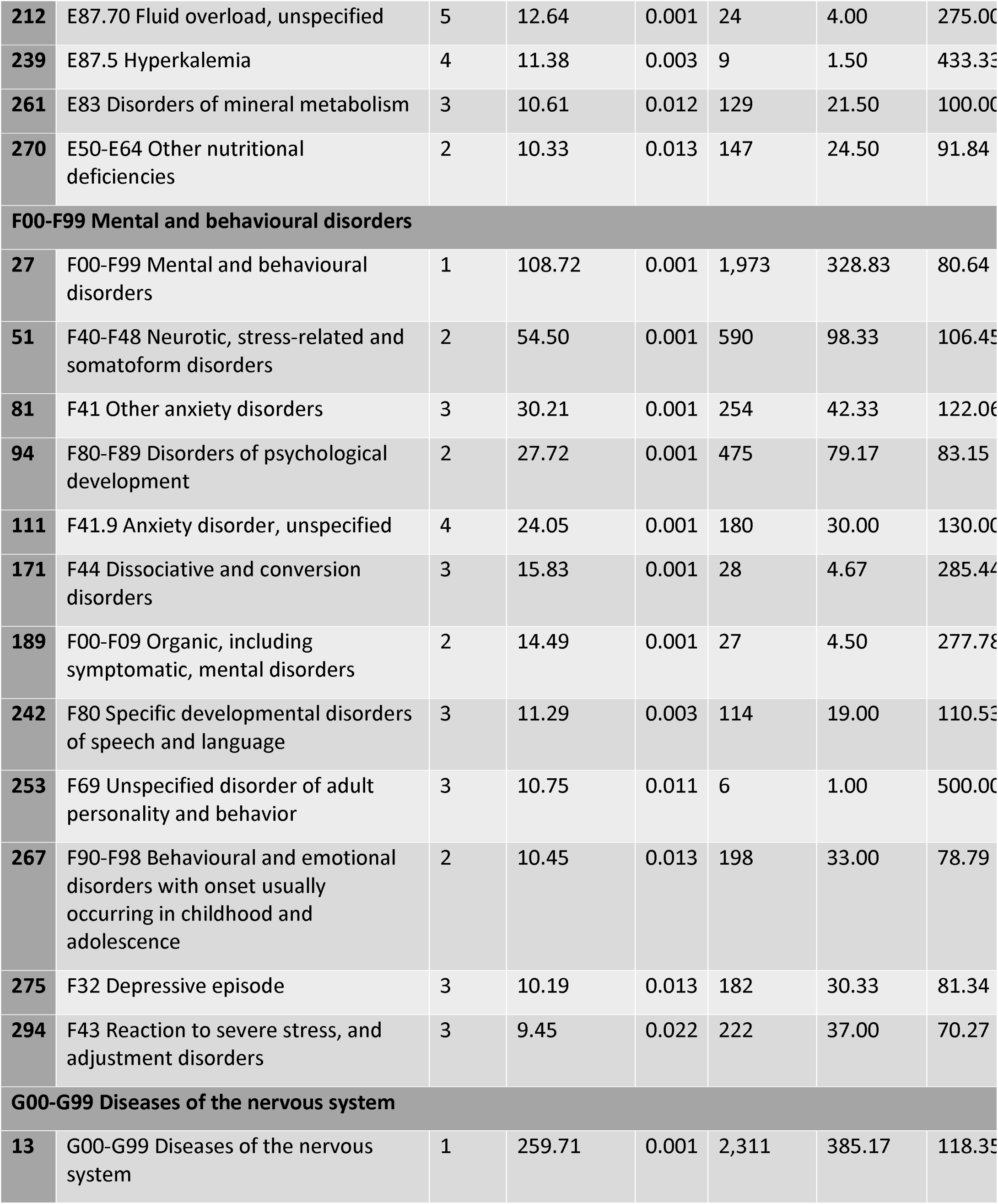
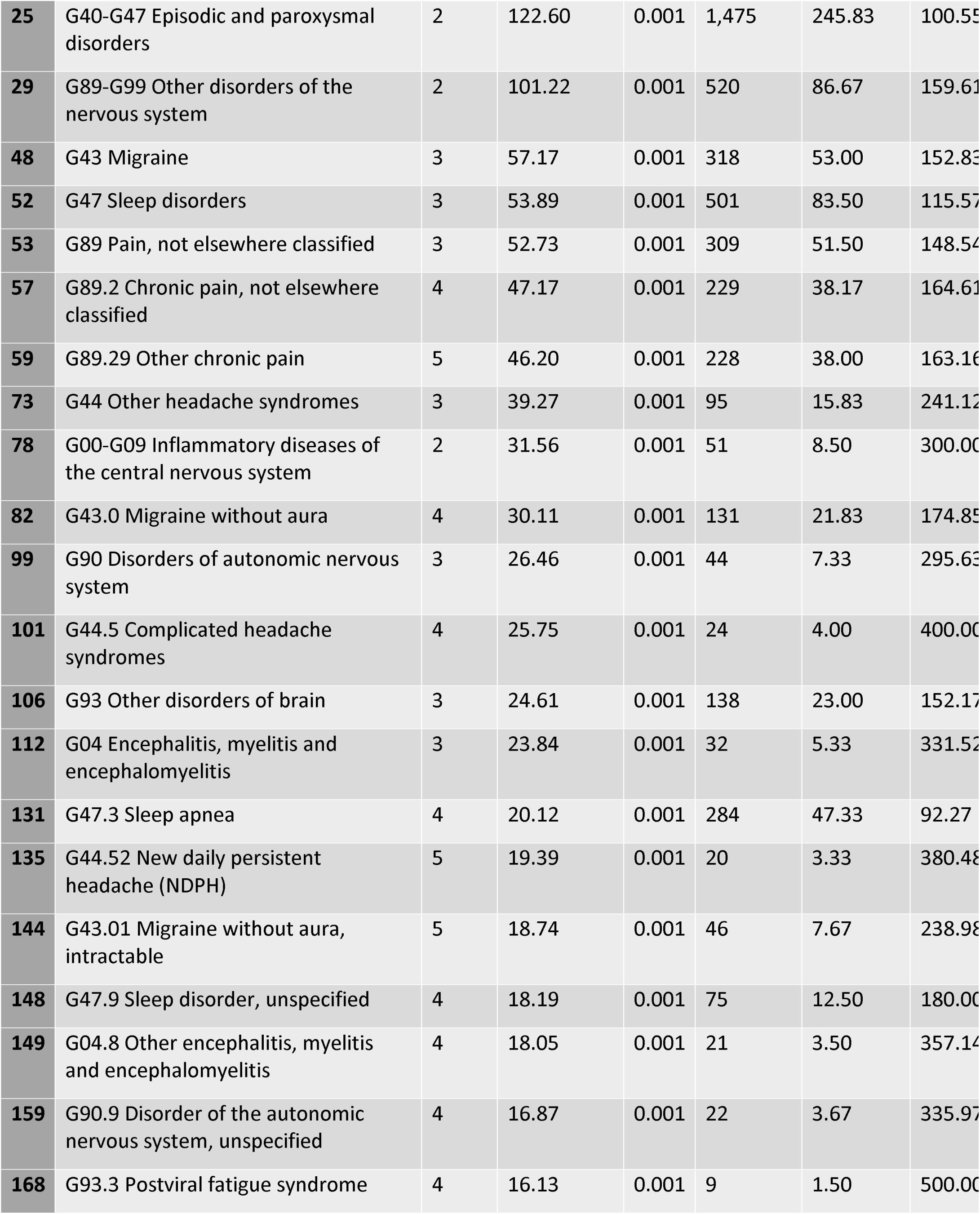
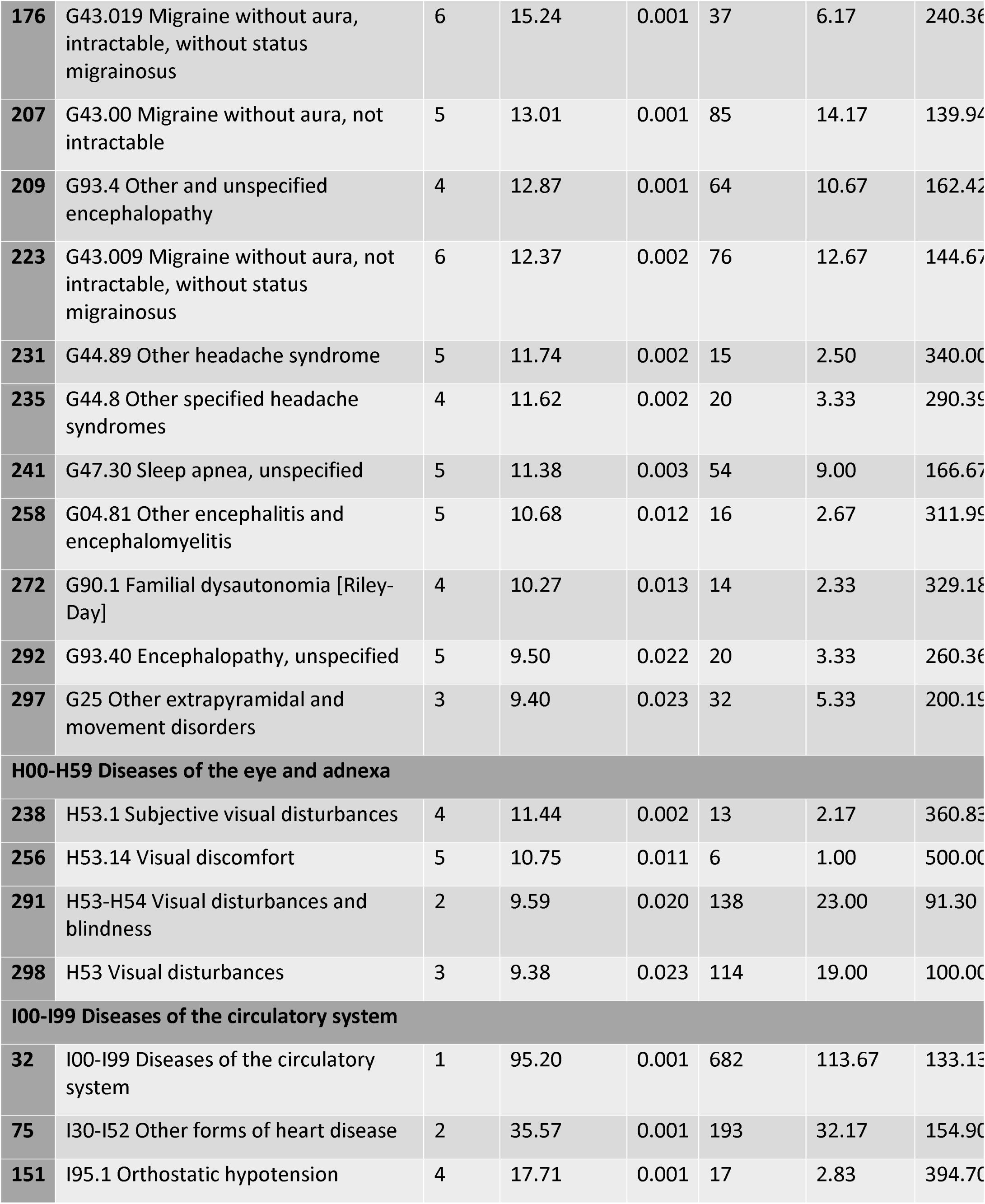
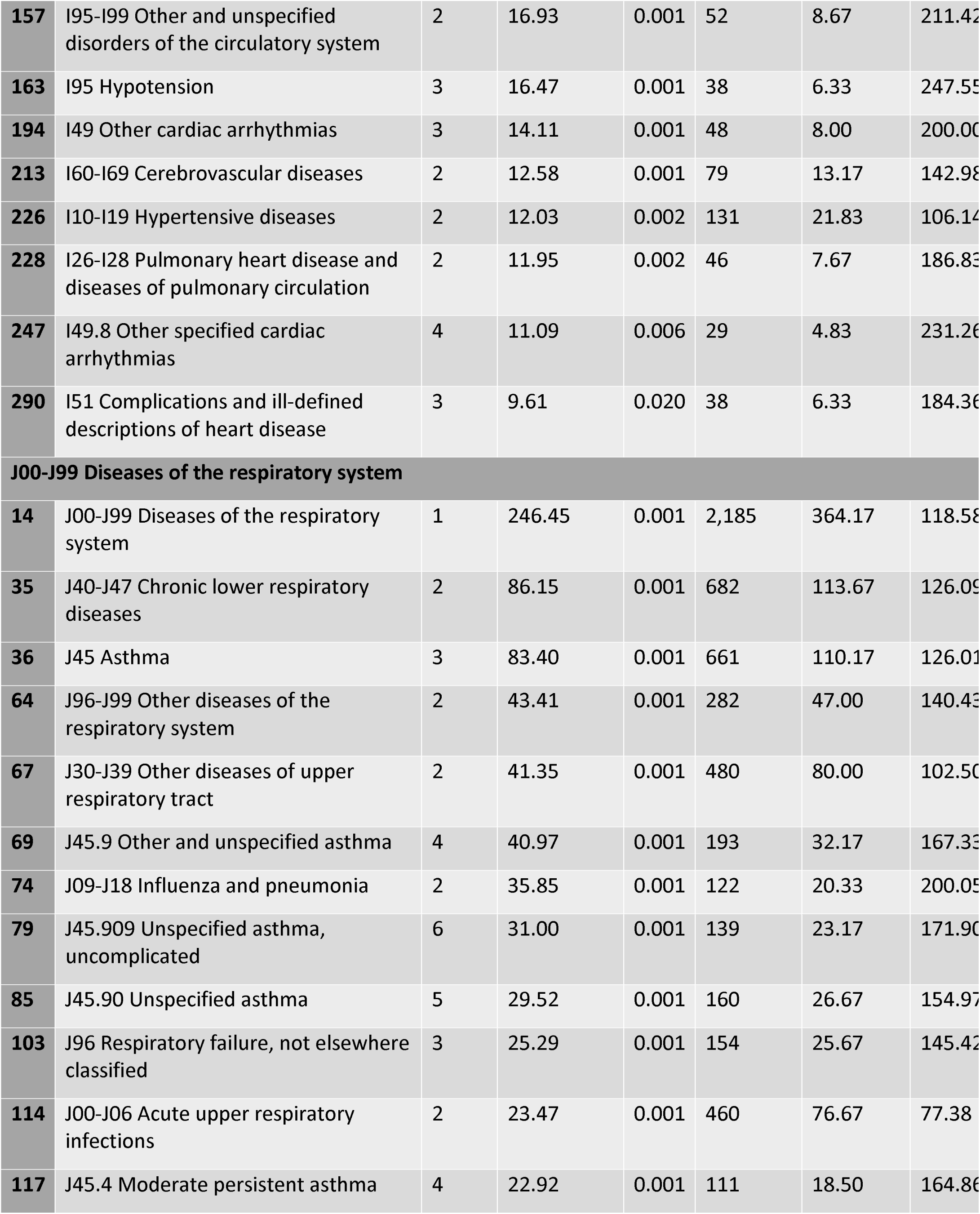
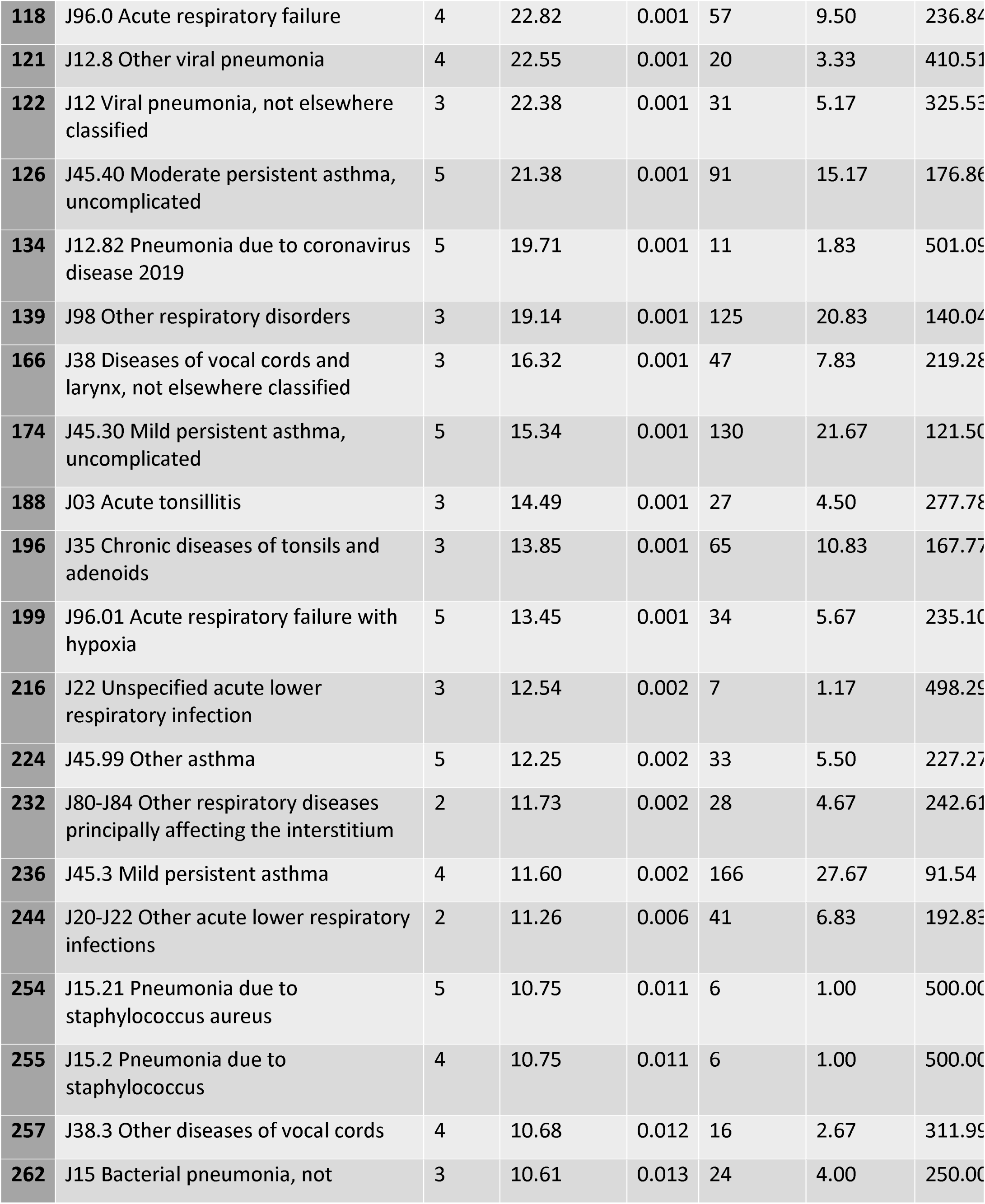
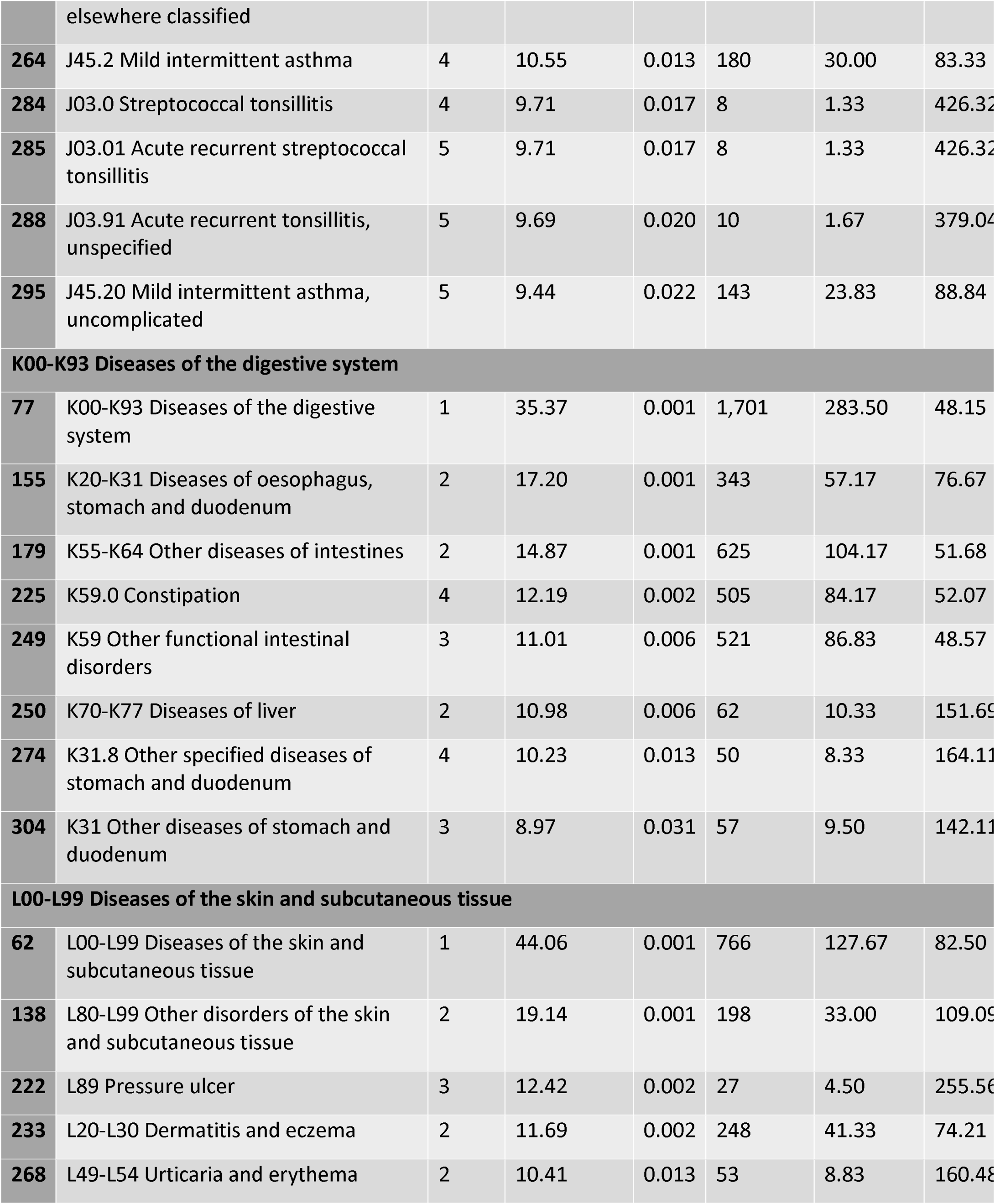
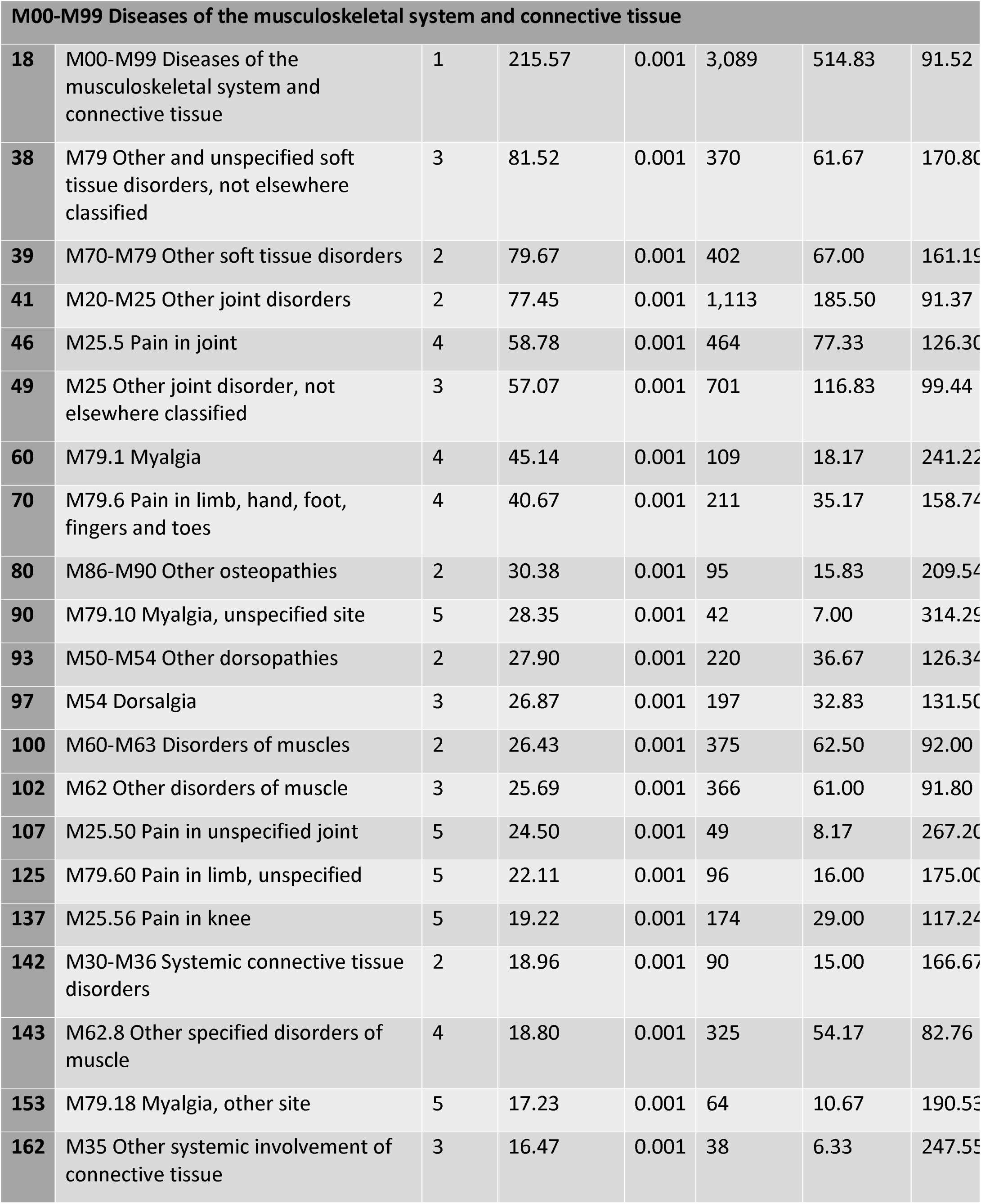
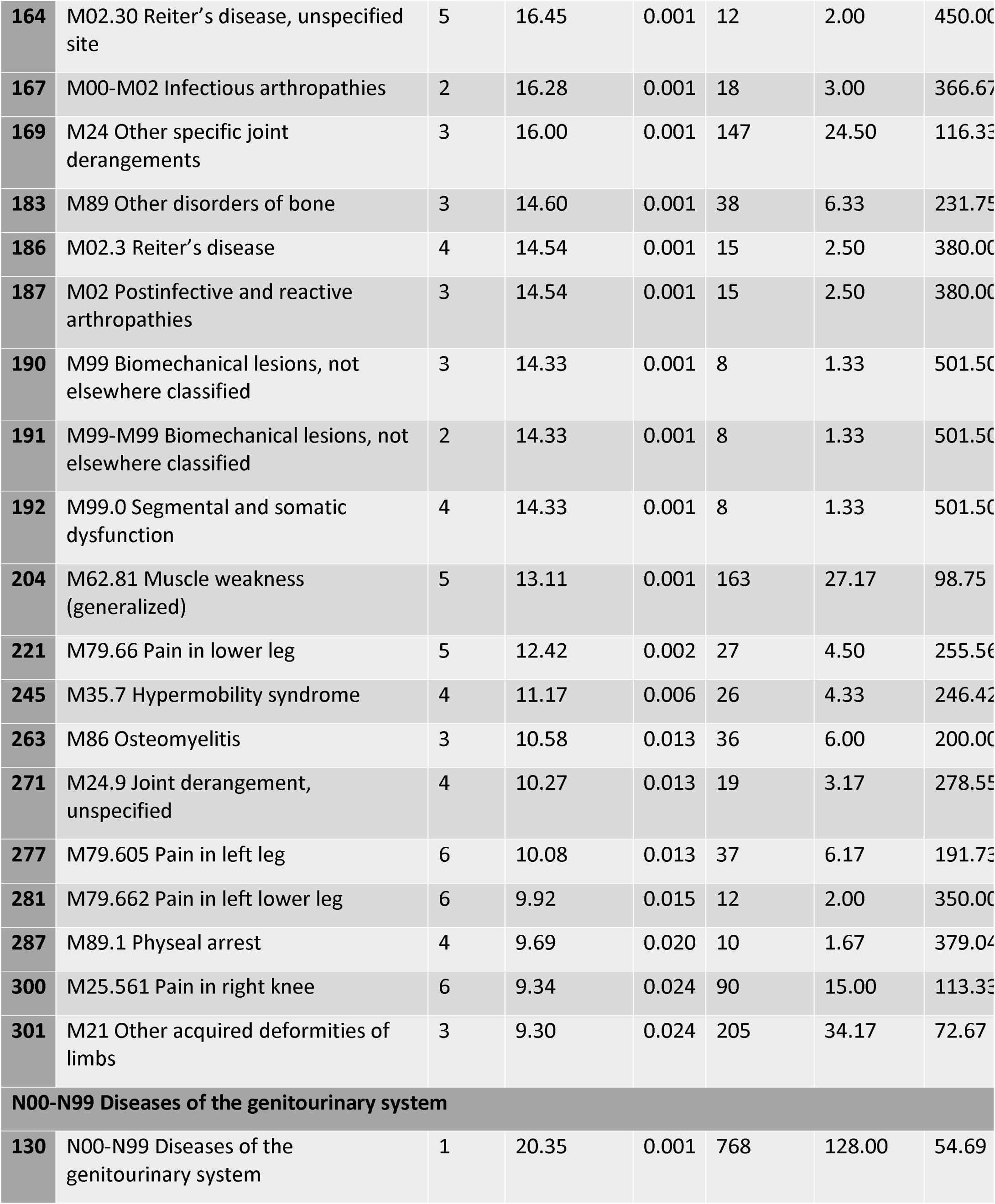
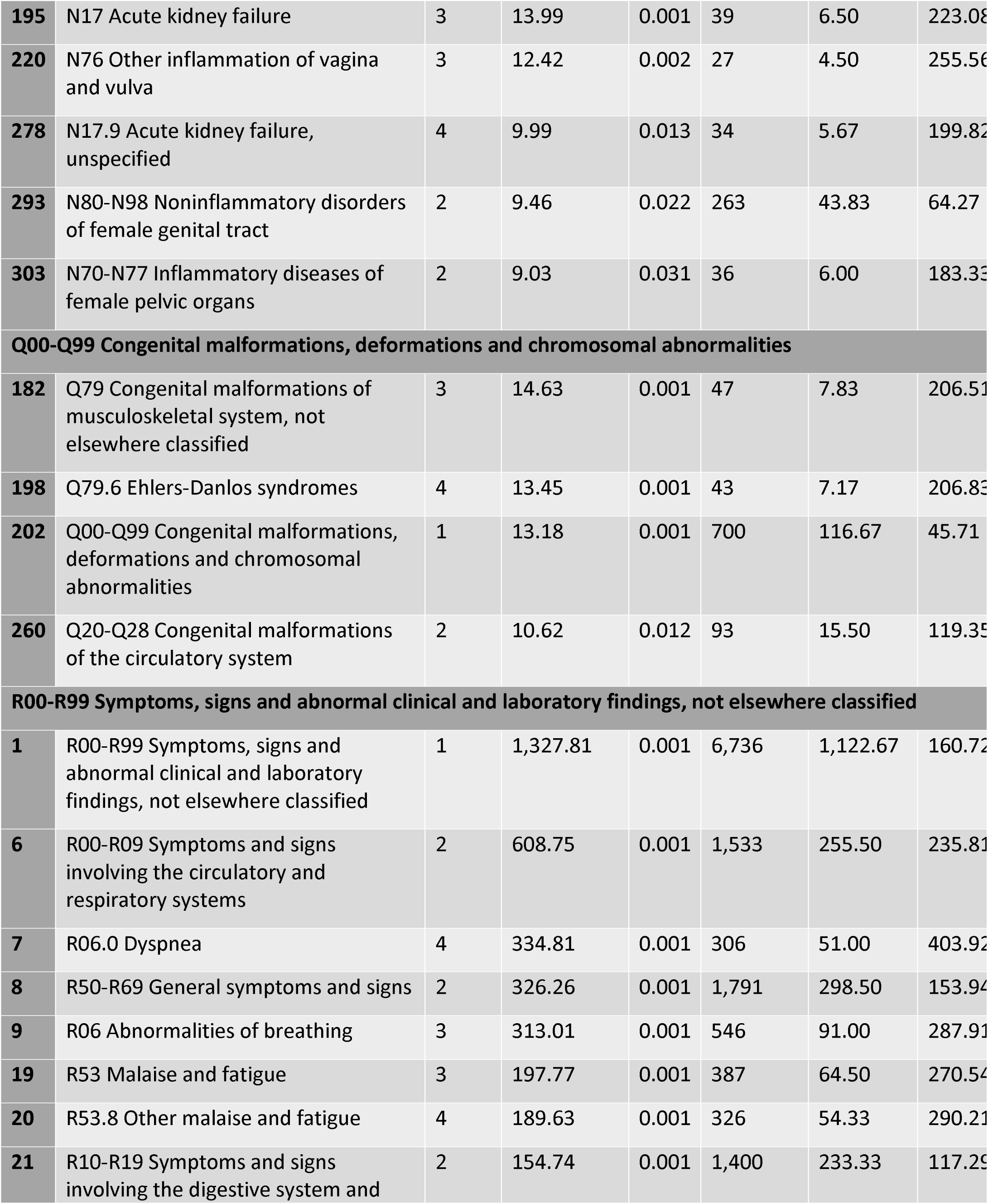
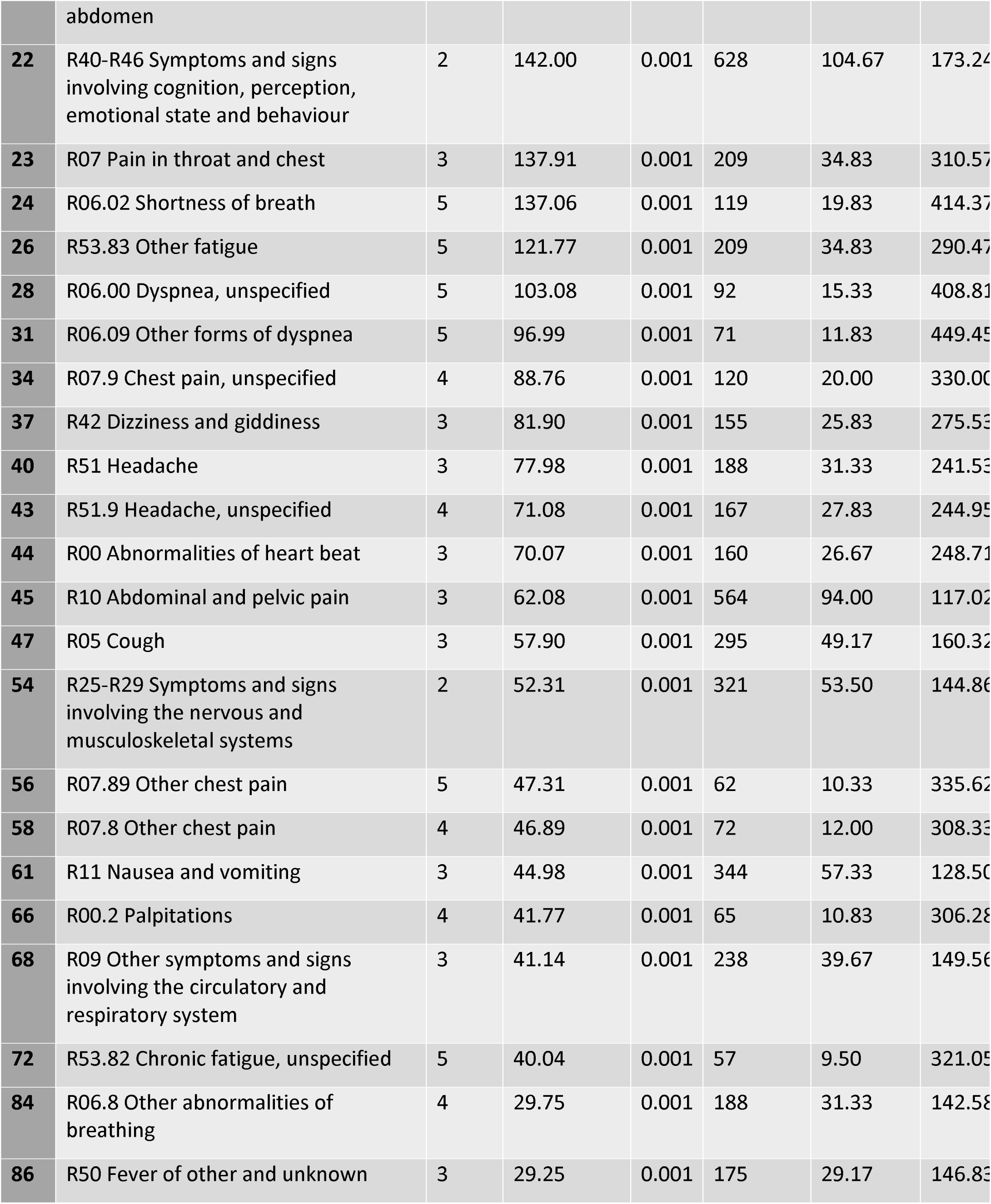
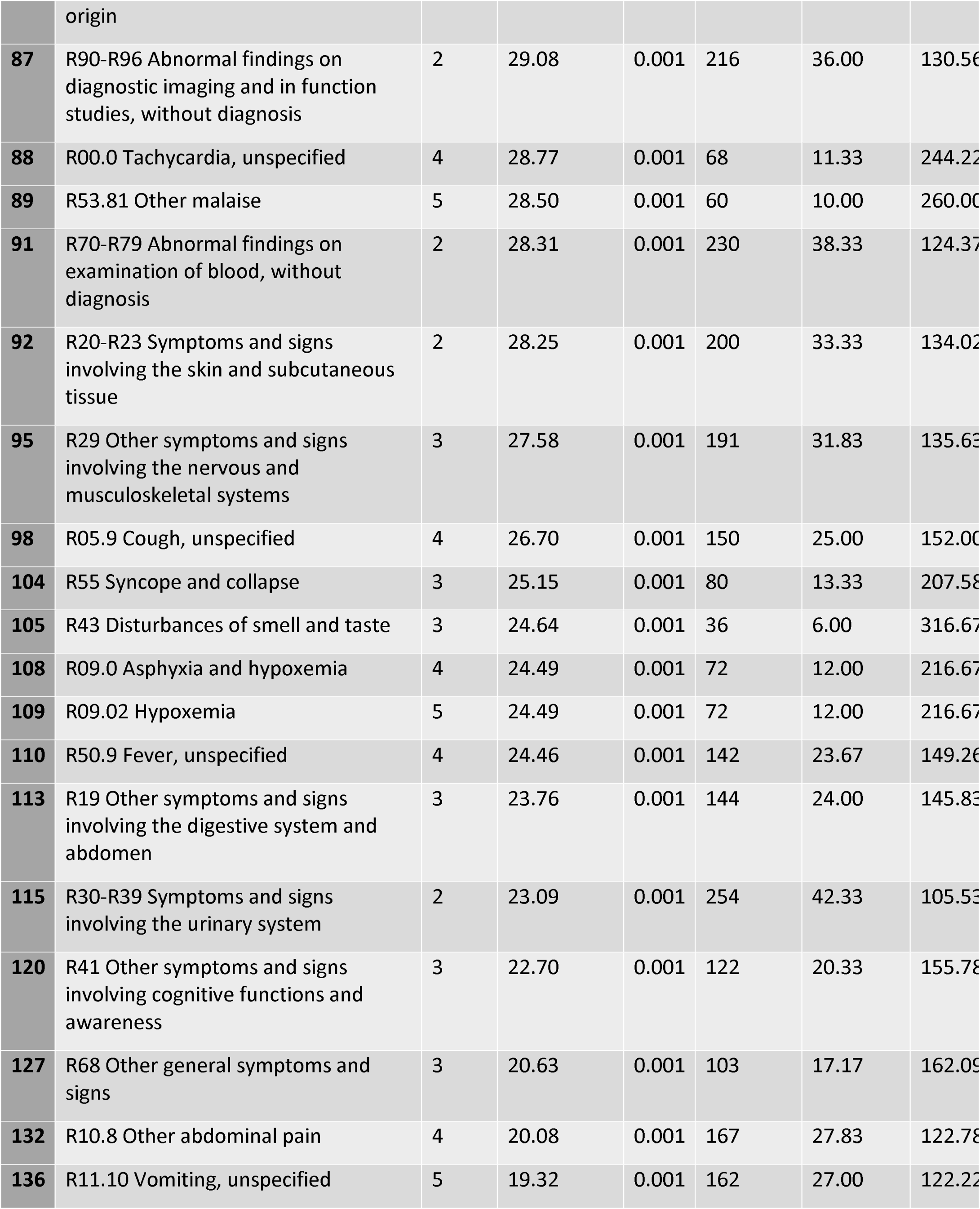
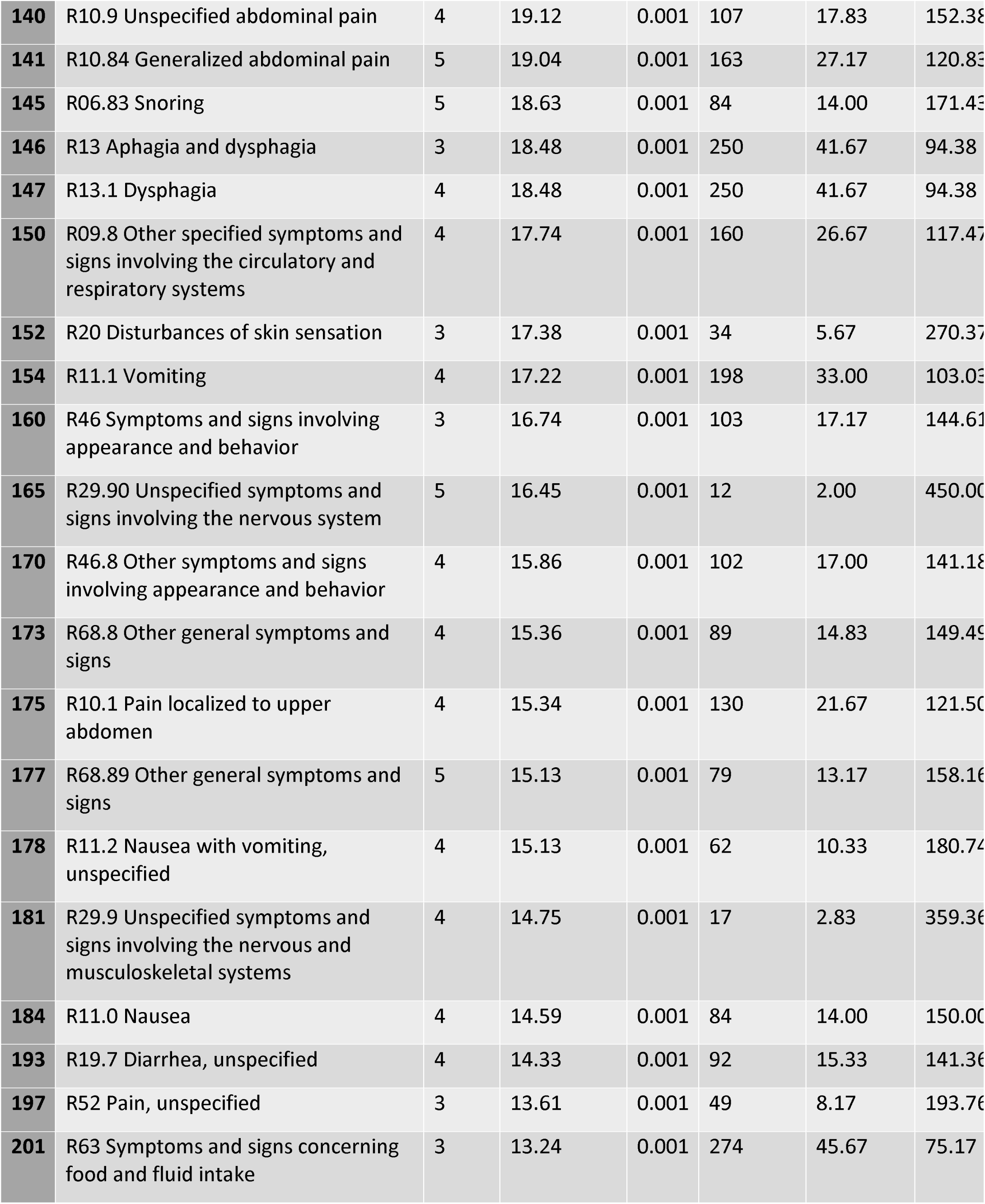
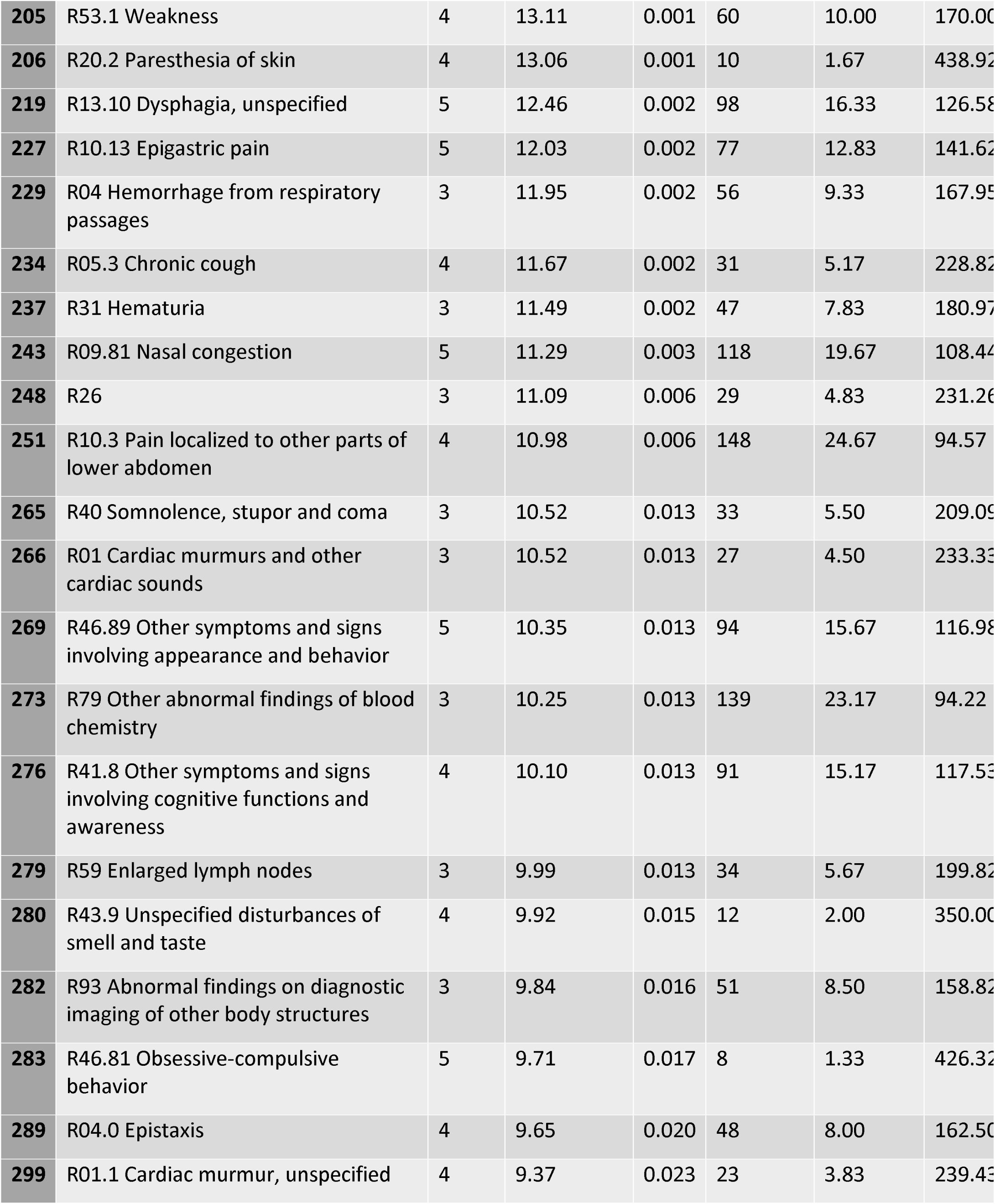
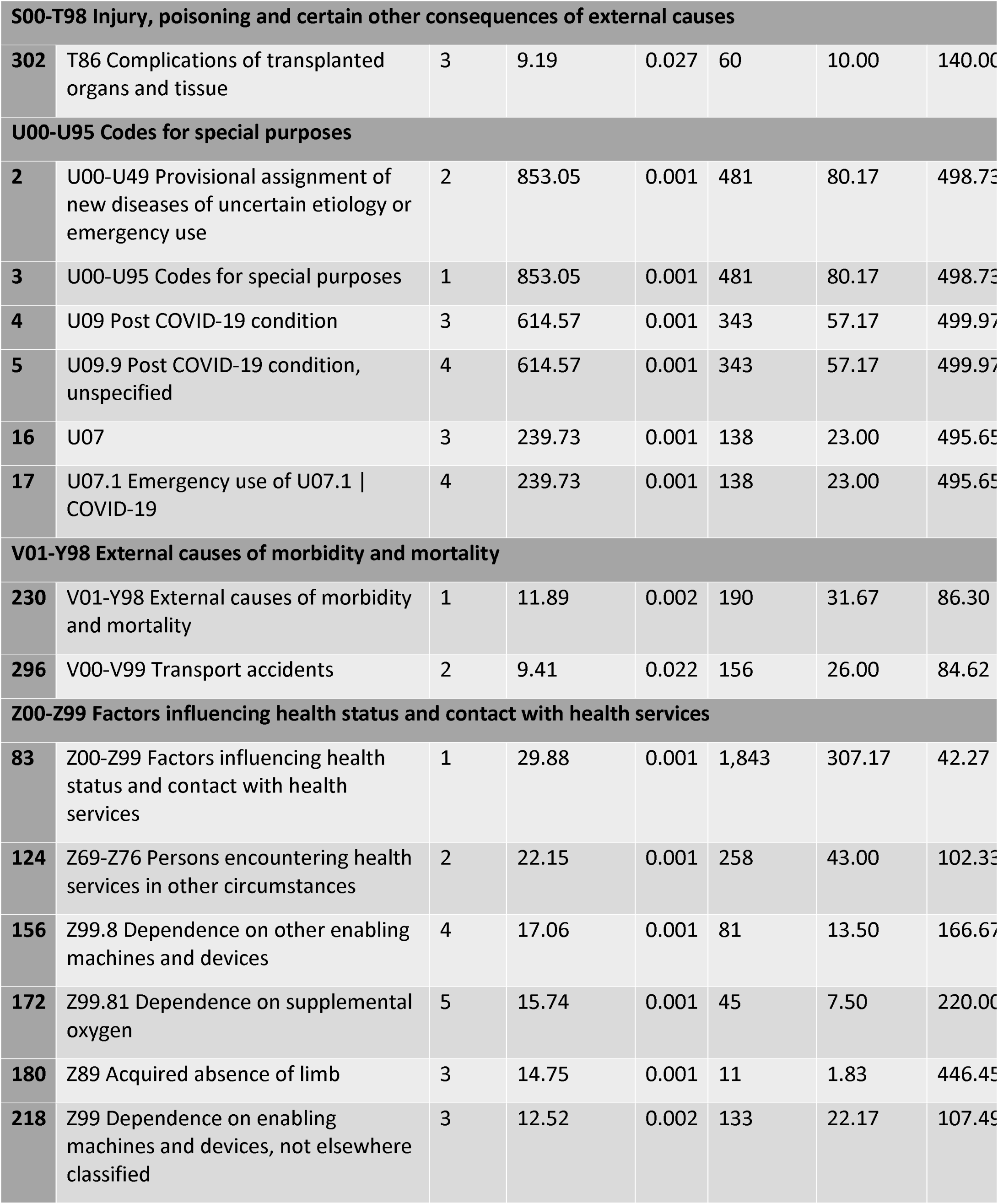
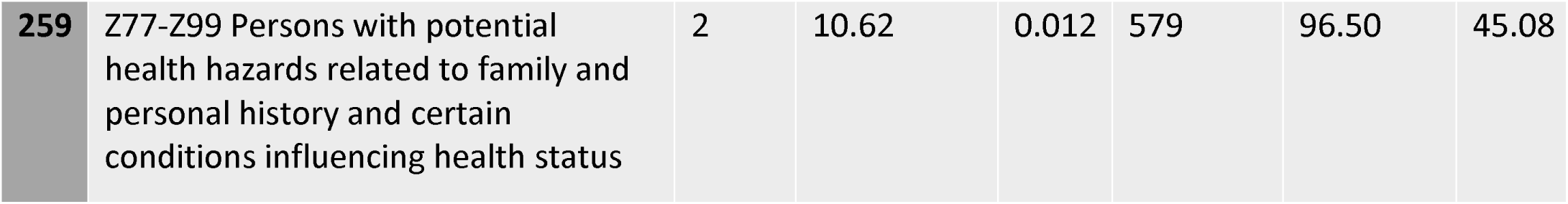
PASC vs COVID negative comparison TreeScan results

## DISCUSSION

We have employed a tree-based data mining approach to detect new-onset conditions presenting in children with a PASC (U09.9 or B94.8) diagnosis. Our data-driven approach identified and ranked features in a large corpus of EHR data. In the first of our two primary analyses, we compared PASC-diagnosed patients with non-PASC-diagnosed SARS-CoV-2 positive patients. The signals identified indicate long term sequelae of PASC that differentiate from having COVID-19.

Significant results from this comparison included both systemic and syndromic manifestations of PASC, with the former describing organ-level dysfunction and the latter focusing on signs and symptoms. Consistent with other studies and biologic mechanisms, we found significant enrichment among children with PASC in cardiac, respiratory, neurologic, psychological, endocrine, gastrointestinal, and musculoskeletal systems(27).

We found the most significant point of difference between PASC and COVID-19 positive children was at the R00-R99 cluster of diagnostic codes *(Symptoms, signs, and abnormal clinical and laboratory findings, not otherwise classified).* These codes designate abnormal clinical findings not elsewhere described that encompass a range of organ systems. This finding is not surprising given the heterogeneity of previously reported manifestations of PASC and the difficulty in gaining clinical consensus to define PASC(28, 29).

Within the R00-R99 category, the codes with the greatest signal are symptoms related to circulatory and respiratory systems. These results align again with prior observations in smaller groups of patients or previous studies, and include dyspnea(12, 16), shortness of breath(16, and other breathing abnormalities(31); chest pain(10, 32, 33); malaise and fatigue(14, 30, 33, 34); cough(14); and abnormalities of heartbeat including palpitations and tachycardia(35, 36). However, within this category we also detected novel signals such as hematuria as well as non-specific symptoms and signs involving the urinary system. Additionally, within this category we also detected novel signals such as hematuria as well as non-specific symptoms and signs involving the urinary system.

Neurologic diagnoses such as headache and migraine were recurring, with several specific codes enriched in the cases. Sleep disturbances and, specifically, sleep apnea, were also present (12, 31).(12, 31). Generalized pain was a key feature in our musculoskeletal system findings, which include non-localized myalgias, joint pain, knee and leg pain, hip pain, upper abdomen and epigastric pain, and generalized soft tissue pain. The recurring appearance of multiple and diverse pain symptoms in this study warrants further investigation to better characterize causal biologic pathways.

Respiratory conditions were also significantly featured in our analysis. Both vocal cord disorders and dysautonomia were present as PASC-related features in our study. Interestingly, new onset asthma was present in our results as a specific diagnosis rather than a constellation of non-specific symptoms. Asthma is one of the few specific health conditions with a significant and strong signal in our study. Respiratory findings associated with PASC have been characterized in detail elsewhere (9).

In contrast to previous work, our study did not identify new-onset diabetes as a persistent long-term complication of COVID-19 (37). Endocrine findings in this study included nutritional deficiencies, volume depletion or fluid overload, and obesity. To our knowledge, these symptoms have not been described in detail elsewhere. It is unclear whether clinicians are recording obesity in the post-infection period as an indication that obesity complicated recovery of SARS-CoV-2 infection or that it points to a patient’s exacerbation of health conditions unrelated to COVID-19.

Gastrointestinal (e.g., *Diseases of esophagus, stomach and duodenum)* and psychological findings (e.g., *Reaction to severe stress, and adjustment disorders)* followed a pattern of TreeScan cuts primarily at non-specific levels. Future work should focus on characterizing co-occurrences of these non-specific symptoms across body systems to detect clinically meaningful patterns. The branches of the ICD hierarchy that are a priori unlikely to be related to COVID-19, e.g. injuries (S00-T98), external causes of morbidity and mortality (V01-Y98), and congenital malformations, deformations, and chromosomal abnormalities (Q00-Q99) serve as a negative control outcome; the relative dearth of diagnoses from these branches serves as a check on residual bias from unmeasured confounding.

We also compared children with a PASC diagnosis code to SARS-CoV-2 test-negative patients. Most cuts from this comparator cohort overlap with the findings where SARS-CoV-2 test-positive patients served as the control. These demonstrate consistency and increase the likelihood that these findings are truly associated with PASC. Additionally, several diagnoses unique to the test-negative cohort were identified, many of which include symptoms relating to acute respiratory infection. Some specific infection codes, such as B25, *Cytomegaloviral Disease* (CMV) were found significant, which may be a result of reactivation in patients infected with SARS-CoV-2(38). Other codes, such as D89, *Other disorders involving the immune mechanism, not elsewhere classified,* or skin conditions such as dermatitis and eczema (L20-L30), were also enriched in the PASC patients when compared to SARS-CoV-2 negative patients. Autoimmunity following an infection has been documented and warrants further investigation in the pediatric population(39). Alternate findings enriched in this analysis include features plausibly associated with COVID-19 infection including acute myocarditis, hypotension, fever, nasal congestion, epistaxis, tachycardia, and gastrointestinal symptoms such as diarrhea, constipation, and gastroenteritis. More serious conditions such as pulmonary embolism and pulmonary edema were also enriched in this population but may be due to follow up care from an acute COVID-19 infection, and not a new finding in the post-acute period. However, these findings warrant further investigation.

Compared to previous work identifying pediatric PASC features by comparing SARS-CoV-2 test-positive to test-negative patients in the PEDSnet EHR, we found several differences. Abnormal liver enzymes, allergies, disorders of teeth/gingiva, and bronchiolitis were all reported as potential PASC-associated features in Rao et al(17), but were not identified as significant cuts in this study when PASC patients were compared to COVID+ or COVID-cohorts, either at the specific code or macro level. Conversely, significant findings in our study which were not present in the results of Rao et al (17), and included sleep disorders, anemias, eye disorders, constipation, sepsis, nutritional deficiencies, hypotension, and various musculoskeletal conditions and soft tissue and muscle disorders.

Finally, we conducted two sensitivity analyses: the first comparing COVID positive to COVID negative patients, and the second comparing PASC-positive to COVID positive patients pre- and post-omicron. By identifying events that occur in the PASC risk window more frequently in COVID-positive patients, we can increase the sensitivity of capturing potential PASC features. This may come at the expense of precision, as we recognize that statistical signals identified in this analysis may correspond to features not truly associated with PASC, either due to confounding resulting from a more heterogeneous population than PASC-diagnosed patients or due to surveillance bias due to COVID-positivity. While there was significant overlap of the features identified in this and the primary analyses, there were also many additional features identified in the COVID-positive to COVID-negative analysis. In our analysis stratified by pre-and post-omicron variant, we found that the pre-omicron era was enriched for metabolic (e.g., E70-E88: metabolic disorders), immunologic (e.g., D80-D89: Certain disorders involving the immune mechanism), and circulatory system (e.g., 195: hypotension) codes, as well as disturbances of smell and taste, while the post-omicron era was enriched for conditions affecting the urinary system (e.g., R31: hematuria) and multiple migraine diagnoses. However, further analyses and additional methodologic rigor to ensure adequate power as well as balance of visit utilization and other potential confounders between the cohorts is warranted and beyond the scope of the current analysis.In our analysis stratified by pre-omicron and omicron variants, we found that the pre-omicron era was enriched for metabolic (e.g., E70-E88: metabolic disorders), immunologic (e.g., D80-D89: Certain disorders involving the immune mechanism), and circulatory system (e.g., 195: hypotension) codes, as well as disturbances of smell and taste, while the omicron era was enriched for conditions affecting the urinary system (e.g., R31: hematuria) and multiple migraine diagnoses. However, further analyses and additional methodologic rigor to ensure adequate power as well as balance of potential confounders between the cohorts is warranted and beyond the scope of the current analysis. Strengths of our study include using a tree-based scan statistic approach in children to detect groups of clinical diagnoses specific to this population, as well as the use of the U09.9 and B94.8 diagnosis codes to do so. Our study is further strengthened by multiple comparison groups diagnoses of PASC rather than surveys or case series analyses. We have confirmed previous findings and provide new insights into how diagnosis clusters relate to each other in importance. Further, an advantage of EHR data is the ability to learn from previous clinical history of patients. In our analysis, we included an 18-month washout period to ensure that the signals we were observing were not due to baseline differences in disease prevalence.

Prior work identifying pediatric PASC-associated features(17) relied on a set of diagnosis clusters developed by the study team to group similar diagnoses. Our study addresses this methodological limitation by a broader approach where diagnosis clusters are tested along all levels of the ICD-10-CM hierarchy. Importantly, among features of PASC found in previous work, our study contributes precise, diagnosis code-level results showing how PASC features are identified in clinical practice. This makes our findings ideally suited for development of rules-based phenotypes for identifying pediatric PASC in EHRs.

At the same time, there were several significant limitations to our study. First, we are at risk of misclassifying patients in our PASC-negative comparator cohort by relying solely on the U09.9 and B94.8 codes to identify cases. There are likely children and adolescents in our comparison cohort with complications of COVID-19 who never receive a formal PASC diagnosis and may either present to a health system with symptoms that the clinician did not distinguish as a complication of COVID-19 or may not receive care for their symptoms at all. However, this would likely bias toward the null. Further, the U09.9 code has only been formally implemented since October 2021, and while the B94.8 code likely served as a proxy for PASC prior to this, the likely lack of uniform code usage, particularly early in the pandemic, introduces potential bias with regards to the timing of cohort entry dates in our cohort during the pandemic. Additionally, for PASC-diagnosed and serology test positive patients in our cohort who had no prior COVID diagnoses or positive viral testing, our imputation of cohort entry dates may not accurately reflect timing of COVID infection, and in the case of PASC-diagnosed patients, PASC onset. Second, our ascertainment of other diagnosis codes is similarly subject to the biases of health care delivery. For example, our findings may disproportionately include diagnoses that are required for utilization *(e.g.,* to justify diagnostic testing) or symptoms that are uncommon and previously associated in clinical practice with COVID-19 and may under-detect changes following COVID-19 in relatively common childhood illness that escape coding practices. Third, we have analyzed the pandemic as a single cross-section and thus our results concern PASC-associated features in aggregate over the time period of the study; the course of PASC may differ by variant and further information may be gained by studying evolution of diagnoses across different portions of the pandemic or over time since infection. Finally, the cohort comprises patients followed in large pediatric health systems, and results may reflect the largely urban demographics of these systems or the practice patterns of pediatric specialists. Only about half of these hospitals offer robust primary care networks, and therefore children seen in tertiary care centers exclusively may not be representative of all children in the US. This is complicated by the fact that children may only visit these health systems occasionally (e.g., ED visits) and receive the majority of their care elsewhere. Nonetheless, despite these limitations, in previous studies, we have successfully replicated similar rates of health outcomes in our database as have been reported in data representative of the general pediatric population.(40, 41) In the COVID-19context, we have reported that 76% of our patient population have no chronic conditions, 12% have chronic conditions, and 11% have complex-chronic conditions,(42) which are similar to patients found in medicated populations(43). Further, the risk of this bias altering the findings of the current study relies on differential impact on cases and controls, and we have mitigated this risk by matching on medical complexity and demographic factors. Finally, we required at least two encounters with the health system 7 days to 18 months prior to cohort entry to ensure history of clinical care within the network.

## CONCLUSION

This systematic evaluation of diagnoses in a large, multi-system national cohort of children with PASC provides a valuable addition to our understanding of this condition’s protean manifestations. Our results can inform the design of further prospective studies to more deeply investigate the patterns identified here to develop improved clinical practice and to direct the study of the biological basis of PASC.

## Data Availability

The results reported here are based on detailed individual-level patient data compiled as part of the RECOVER program. Due to the high risk of reidentification based on the number of unique patterns in the date, patient privacy regulations prohibit us from releasing the data publicly. The data are maintained in a secure enclave, with access managed by the program coordinating center to remain compliant with regulatory and program requirements. Please direct requests to access the data, either for reproduction of the work reported here or for other purposes, to.

## CONTRIBUTORSHIP STATEMENT

Dr. Bailey, Dr. Lorman and Ms. Razzaghi had full access to all of the data in the study and take responsibility for the integrity of the data and the accuracy of the data analysis.

Dr. Lee, Dr. Lorman, and Ms. Razzaghi were responsible for the study conceptualization and design.

Dr. Lee, Dr. Lorman, Ms. Razzaghi, and Dr. Chen were involved in statistical analyses.

Dr. Lee, Dr. Lorman, and Ms. Razzaghi contributed to the drafting of the manuscript.

All authors contributed to interpretation of the data and review of the manuscript.

Dr. Rao, Dr. Lee, Dr. Bailey, and Dr. Forrest were involved in obtaining funding for the study.

The content is solely the responsibility of the authors and does not necessarily represent the official views of the RECOVER Program, the NIH or other funders.

## ACKNOWLEDGEMENTS

The authors gratefully acknowledge the contributions of Miranda Higginbotham.

## Footnotes

1 More information on the PEDSnet RECOVER cohort is available at https://github.com/PEDSnet/Data_Models_Public/blob/master/PEDSnet/docs/RECOVER%20Cohort.md

