## Supplementary appendix for "Understanding pediatric long COVID using a tree-based scan statistic approach: An EHR-based cohort study from the RECOVER Program"

**Sensitivity analysis methods**

In a sensitivity analysis, we compared SARS-CoV-2 infected to uninfected patients. We defined the COVID-positive and COVID-negative cohorts using the same inclusion criteria as in the primary analyses and defined their cohort entry dates in the same way. Outcomes were evaluated identically as in the primary analysis during the 28-179 day period following the index date. Unlike the primary analysis, which used 5:1 nearest neighbor matching, in this analysis we used 1:1 nearest neighbor matching, requiring exact matches on age group at cohort entrance and cohort entry month.

**Sensitivity analysis results**

The sensitivity analysis comparing patients with COVID-19 infection to those without COVID-19 infection which employed 1:1 nearest neighbor matching was well-balanced with absolute SMD<0.1 for all covariates (Supplementary Figure 1c). Major findings which were not present in the primary analyses are summarized in Supplementary Table 2, and the full list of findings can be found in Supplementary Table 3.

**Supplementary Table 1: COVID positive vs negative sensitivity analysis, population characteristics after matching**

|  | Level | Overall N (%) | COVID-negative N (%) | COVID-19 N (%) |
| --- | --- | --- | --- | --- |
| n |  | 255,134 | 127,567 | 127,567 |
| Age at cohort entrance (%) | <1 | 27060 (10.6) | 13530 (10.6) | 13530 (10.6) |
|  | 1-4 | 67886 (26.6) | 33943 (26.6) | 33943 (26.6) |
|  | 5-11 | 79112 (31.0) | 39556 (31.0) | 39556 (31.0) |
|  | 12-15 | 43096 (16.9) | 21548 (16.9) | 21548 (16.9) |
|  | 16-20 | 37980 (14.9) | 18990 (14.9) | 18990 (14.9) |
| Sex (%) | Female | 124682 (48.9) | 62453 (49.0) | 62229 (48.8) |
|  | Male/Other/Unknown | 130452 (51.1) | 65114 (51.0) | 65338 (51.2) |
| Race/ethnicity (%) | Non-Hispanic Asian/PI | 10000 ( 3.9) | 5025 ( 3.9) | 4975 ( 3.9) |
|  | Non-Hispanic Black/AA | 52341 (20.5) | 25495 (20.0) | 26846 (21.0) |
|  | Hispanic | 41452 (16.2) | 20168 (15.8) | 21284 (16.7) |
|  | Multiple | 10983 ( 4.3) | 5384 ( 4.2) | 5599 ( 4.4) |
|  | Other/Unknown | 26543 (10.4) | 13459 (10.6) | 13084 (10.3) |
|  | Non-Hispanic White | 113815 (44.6) | 58036 (45.5) | 55779 (43.7) |
| Cohort entry month (%) | Mar 2020 | 462 (0.2) | 231 (0.2) | 231 (0.2) |
|  | Apr 2020 | 1006 (0.4) | 503 (0.4) | 503 (0.4) |
|  | May 2020 | 1656 (0.6) | 828 (0.6) | 828 (0.6) |
|  | Jun 2020 | 1862 (0.7) | 931 (0.7) | 931 (0.7) |
|  | Jul 2020 | 3662 (1.4) | 1831 (1.4) | 1831 (1.4) |
|  | Aug 2020 | 2742 (1.1) | 1371 (1.1) | 1371 (1.1) |
|  | Sep 2020 | 2216 (0.9) | 1108 (0.9) | 1108 (0.9) |
|  | Oct 2020 | 4824 (1.9) | 2412 (1.9) | 2412 (1.9) |
|  | Nov 2020 | 13996 (5.5) | 6998 (5.5) | 6998 (5.5) |
|  | Dec 2020 | 15610 (6.1) | 7805 (6.1) | 7805 (6.1) |
|  | Jan 2021 | 13406 (5.3) | 6703 (5.3) | 6703 (5.3) |
|  | Feb 2021 | 6260 (2.5) | 3130 (2.5) | 3130 (2.5) |
|  | Mar 2021 | 6054 (2.4) | 3027 (2.4) | 3027 (2.4) |
|  | Apr 2021 | 8190 (3.2) | 4095 (3.2) | 4095 (3.2) |
|  | May 2021 | 4306 (1.7) | 2153 (1.7) | 2153 (1.7) |
|  | Jun 2021 | 1682 (0.7) | 841 (0.7) | 841 (0.7) |
|  | Jul 2021 | 3432 (1.3) | 1716 (1.3) | 1716 (1.3) |
|  | Aug 2021 | 12248 (4.8) | 6124 (4.8) | 6124 (4.8) |
|  | Sep 2021 | 14886 (5.8) | 7443 (5.8) | 7443 (5.8) |
|  | Oct 2021 | 9218 (3.6) | 4609 (3.6) | 4609 (3.6) |
|  | Nov 2021 | 10294 (4.0) | 5147 (4.0) | 5147 (4.0) |
|  | Dec 2021 | 28786 (11.3) | 14393 (11.3) | 14393 (11.3) |
|  | Jan 2022 | 58826 (23.1) | 29413 (23.1) | 29413 (23.1) |
|  | Feb 2022 | 9044 (3.5) | 4522 (3.5) | 4522 (3.5) |
|  | Mar 2022 | 3430 (1.3) | 1715 (1.3) | 1715 (1.3) |
|  | Apr 2022 | 5296 (2.1) | 2648 (2.1) | 2648 (2.1) |
|  | May 2022 | 11740 (4.6) | 5870 (4.6) | 5870 (4.6) |
| Institution (%) | A | 14843 (5.8) | 7484 (5.9) | 7359 (5.8) |
|  | B | 34728 (13.6) | 17206 (13.5) | 17522 (13.7) |
|  | C | 58580 (23.0) | 29445 (23.1) | 29135 (22.8) |
|  | D | 11494 (4.5) | 5845 (4.6) | 5649 (4.4) |
|  | E | 15147 (5.9) | 7420 (5.8) | 7727 (6.1) |
|  | F | 13462 (5.3) | 6912 (5.4) | 6550 (5.1) |
|  | G | 7048 (2.8) | 3574 (2.8) | 3474 (2.7) |
|  | H | 50714 (19.9) | 25098 (19.7) | 25616 (20.1) |
|  | I | 49118 (19.3) | 24583 (19.3) | 24535 (19.2) |
| Test Location (%) | Emergency Department | 67886 (26.6) | 32236 (25.3) | 35650 (27.9) |
|  | Inpatient | 11383 (4.5) | 5688 (4.5) | 5695 (4.5) |
|  | Other/Unknown | 1706 (0.7) | 795 (0.6) | 911 (0.7) |
|  | Outpatient Office | 69073 (27.1) | 35338 (27.7) | 33735 (26.4) |
|  | Outpatient: Test Only | 105086 (41.2) | 53510 (41.9) | 51576 (40.4) |
| PMCA Index (%) | Non-chronic | 153149 (60.0) | 77755 (61.0) | 75394 (59.1) |
|  | Chronic non-complex | 83636 (32.8) | 41482 (32.5) | 42154 (33.0) |
|  | Complex chronic | 18349 (7.2) | 8330 (6.5) | 10019 (7.9) |

**Supplementary Figure 1a Standardized Mean Differences before and after matching, non-MIS-C PASC compared to COVID positives**

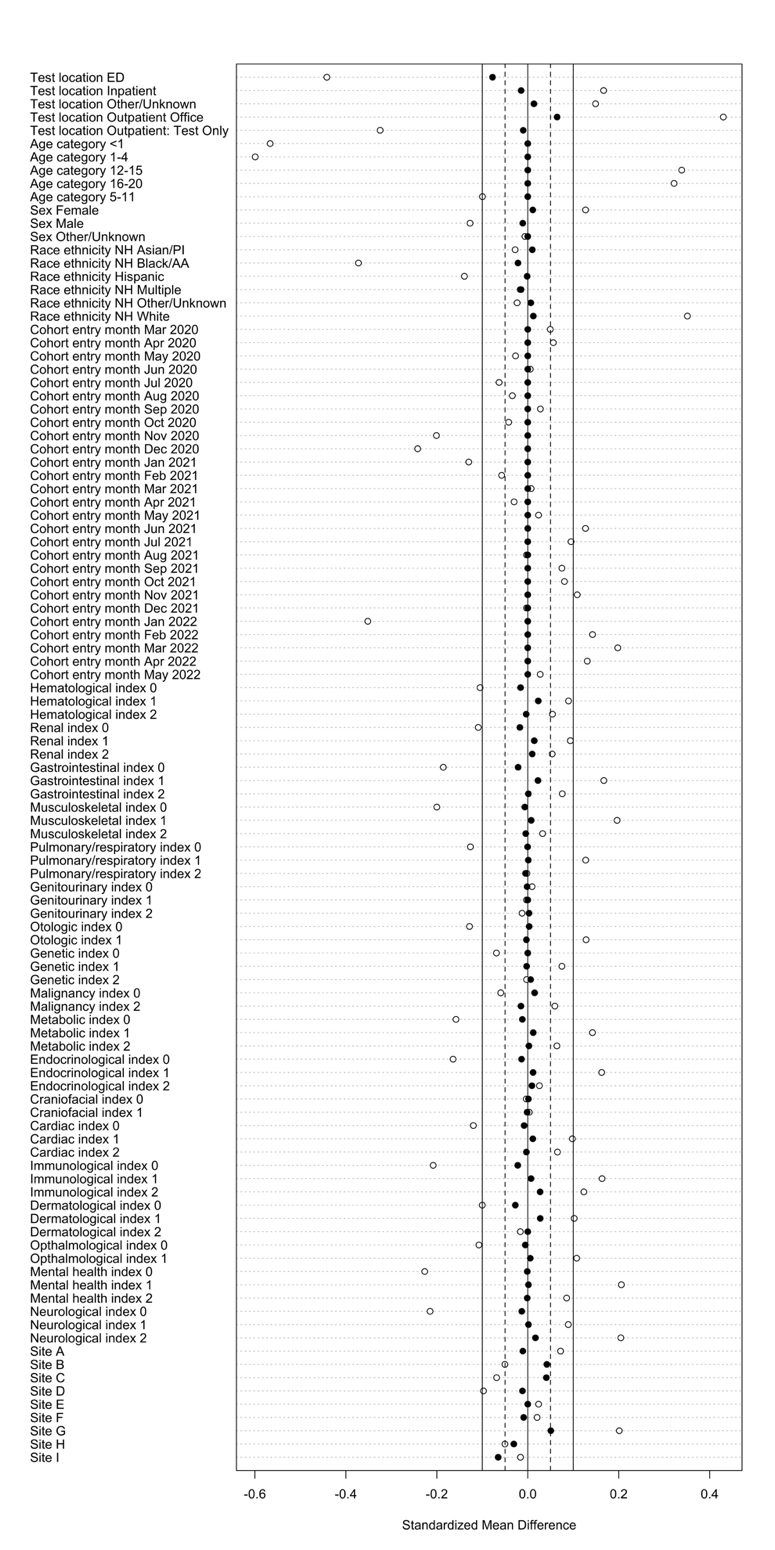

^1^ Standardized Mean Difference (SMD), computed as the difference in means between the positive and negative cohorts divided by the pooled standard deviation. An SMD less than 0.1 in absolute value is often taken to indicate a negligible difference in means between the two groups[25]

**Supplementary Figure 1b Standardized Mean Differences before and after matching, non-MIS-C PASC compared to COVID negatives**

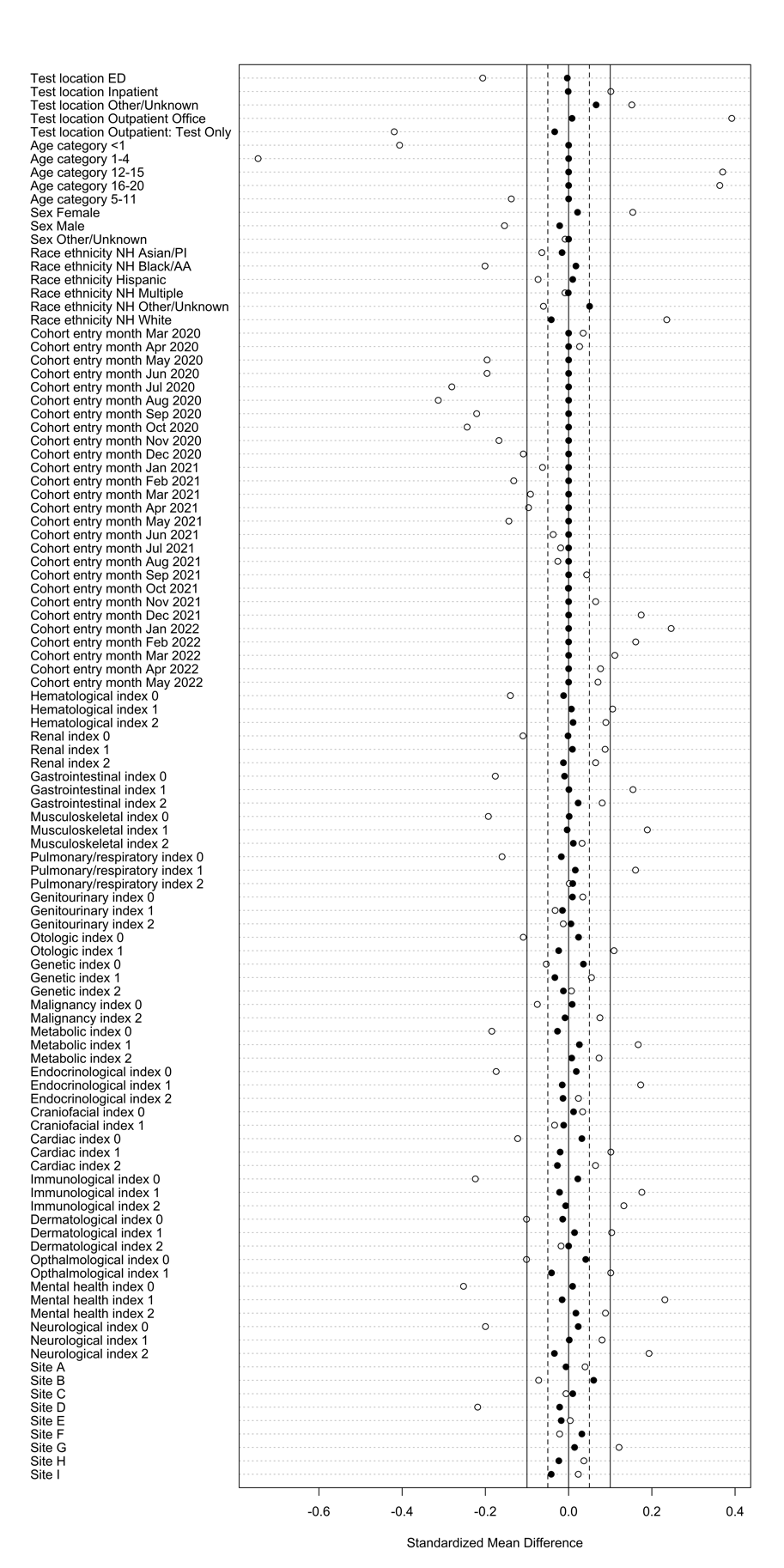

**Supplementary Figure 1c Standardized Mean Differences before and after matching, COVID positives compared to COVID negatives**

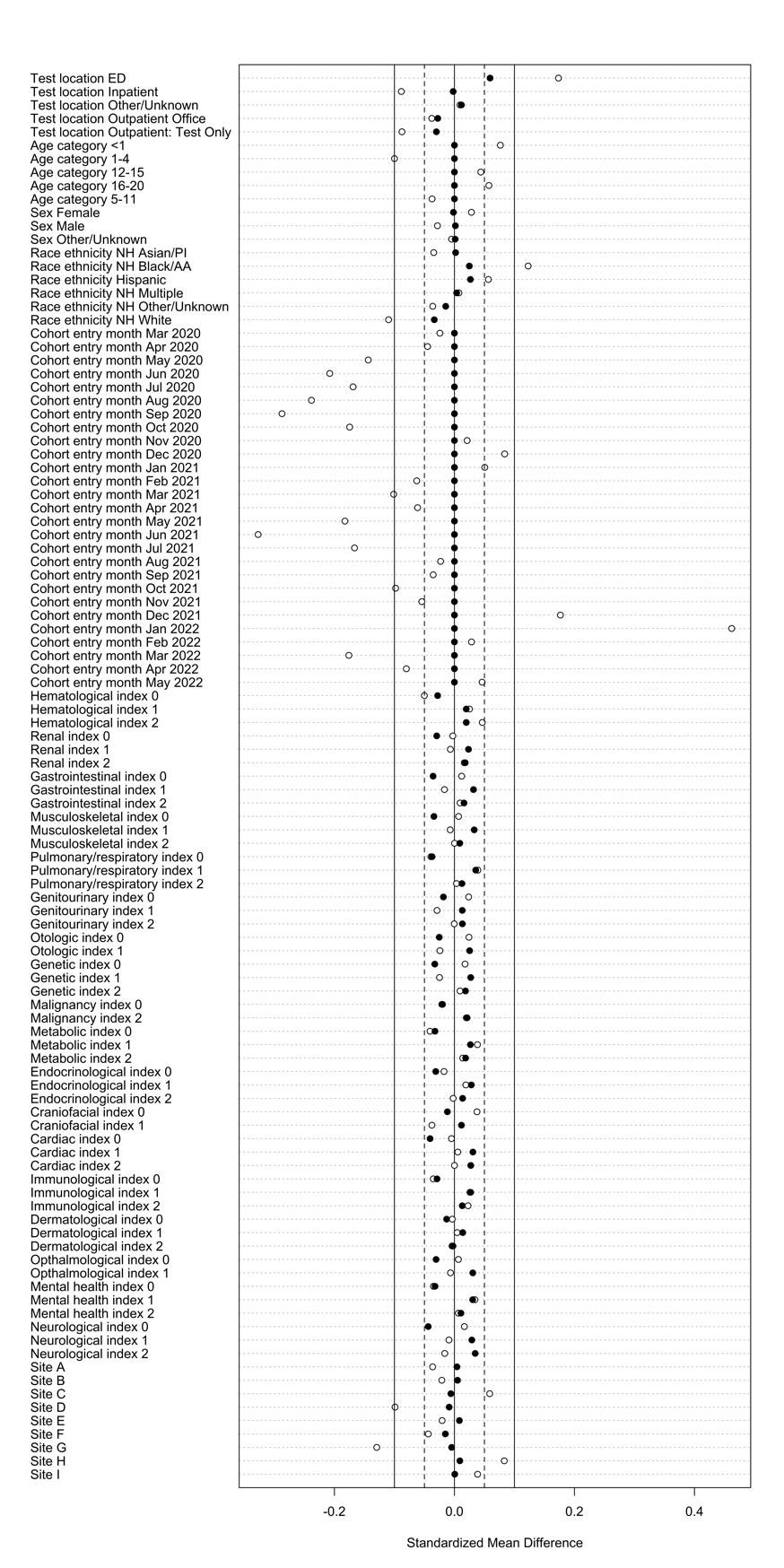

**Supplementary Table 2: Summary of significant features in sensitivity analysis that were not detected in primary analyses**

| Major systemic findings, or conditions (general and specific) | |
| --- | --- |
| Diseases of the nervous system | Epilepsy and recurrent seizures, other and unspecified polyneuropathies, extrapyramidal and movement disorders, cerebral palsy, disorders of autonomic nervous system |
| Mental and behavioral disorders | Developmental disorders of speech and language, severe intellectual disabilities, opioid related disorders |
| Diseases of the respiratory system | pharyngitis, laryngitis, other nasal disorders, allergic rhinitis, pulmonary collapse, atelectasis, hypertrophy of adenoids/tonsils, |
| Diseases of the circulatory system | Hypertension, heart failure, thrombosis, conduction disorder, tricuspid valve disorders, aortic ectasia, aschemic heart disease |
| Disease of the musculoskeletal system and connective tissue | Osteonecrosis |
| Diseases of the digestive system | Dental caries, Stomatitis, oral mucositis, gastrostomy complications, gingivitis, teething syndrome, diseases of pulp and periapical tissues, diseases of lip and oral mucosa, acute pancreatisis, cholangitis, unspecified ileu, chronic gastritis, other diseases of esophagus, other diseases of pancreas |
| Diseases of blood | Aplastic anemias, pancytopenia, neutropenia, disorders of white blood cells/decreased white blood cell counts, thrombocytopenia, Hb-SS disease, hypogammaglobulinemia, agranulocytosis, coagulation defects, other diseases of spleen |
| Endocrine, nutritional and metabolic diseases | Hypo-osmolality and hyponatremia, hypokalemia, disorders of magnesium metabolism, disorders of adrenal gland, disorders of phosphorus metabolism, hypomagnesemia, adrenocortical insufficiency, acidosis, type 1diabetes mellitus, malnutrition, hypocalcemia |
| Renal | Disorders resulting from impaired renal tubular function, cystitis |
| Disorders of the eye and ear | Conjunctivitis, disorders of optic nerve, disorders of eyelid, optic atrophy, otitis media, otalgia, impacted cerumen, other disorders of tympanic membrane, astigmatism |
| Neoplasms | Lymphoid leukemia, lymphoma, secondary malignant neoplasm of lung, secondary malignant neoplasm of bone and bone marrow, lymphocytopenia |
| Major syndromic findings, or symptoms | |
| Circulatory and respiratory signs and symptoms | Sequelae of cerebrovascular disease, snoring, bacteremia, tachypnea, aortic ectasia |
| GI signs/symptoms | Enterocolitis, gastroenteropathy, candida stomatitis, polyuria/urinary incontinence, ascites, intestinal obstruction |
| Cognition, perception, emotional state | Hearing loss, visual disturbances, failure to thrive, somnolence, reduced mobility |
| Musculoskeletal | Segmental and somatic dysfunction, biomechanical lesions |
| General signs and symptoms | Sepsis, lack of expected normal physiological development in childhood, feeding difficulties, febrile convulsions, acute cystitis, epileptic spasms, sinusitis |
| Skin | Mycoses, candidiasis of skin and nail, dermatitis, erhythematous condition, cellutitis, acanthosis nigricans, rash, anesthesia of skin, cutaneous abscess, dry skin, other follicular disorders |
| Viral and bacterial infections | Enterovirus infection, infective mononucleosis, viral infections as cause of diseases classified elsewhere, influenza |

**Supplementary Table 3: COVID positive vs negative comparison TreeScan results**

| Cut | Node | Tree Level | Log Likelihood Ratio | P value | N cases observed | N cases expected | Percent excess cases |
| --- | --- | --- | --- | --- | --- | --- | --- |
| A00-B99 Certain infections and parasitic diseases | | | | | | | |
| 8 | A00-B99 Certain infections and parasitic diseases | 1 | 924.67 | 0.001 | 22,210 | 11,105.0 | 28.65 |
| 18 | B95-B98 Bacterial, viral and other infectious agents | 2 | 362.94 | 0.001 | 5,844 | 2,922.0 | 34.87 |
| 24 | B25-B34 Other viral diseases | 2 | 307.69 | 0.001 | 6,939 | 3,469.5 | 29.56 |
| 28 | B34 Viral infection of unspecified site | 3 | 282.89 | 0.001 | 6,203 | 3,101.5 | 29.97 |
| 30 | B97 Viral agents as the cause of diseases classified elsewhere | 3 | 281.96 | 0.001 | 3,906 | 1,953.0 | 37.53 |
| 53 | B34.9 Viral infection, unspecified | 4 | 177.94 | 0.001 | 5,295 | 2,647.5 | 25.78 |
| 58 | B97.8 Other viral agents as the cause of diseases classified elsewhere | 4 | 163.12 | 0.001 | 3,053 | 1,526.5 | 32.39 |
| 67 | B97.89 Other viral agents as the cause of diseases classified elsewhere | 5 | 147.41 | 0.001 | 2,958 | 1,479.0 | 31.30 |
| 82 | A00-A09 Intestinal infectious diseases | 2 | 128.00 | 0.001 | 2,068 | 1,034.0 | 34.82 |
| 95 | B94 Sequelae of other and unspecified infectious and parasitic diseases | 3 | 108.13 | 0.001 | 156 | 78.0 | 100.00 |
| 96 | B90-B94 Sequelae of infectious and parasitic diseases | 2 | 108.13 | 0.001 | 156 | 78.0 | 100.00 |
| 100 | B94.8 Sequelae of other specified infectious and parasitic diseases | 4 | 105.36 | 0.001 | 152 | 76.0 | 100.00 |
| 124 | A08 Viral and other specified intestinal infections | 3 | 85.91 | 0.001 | 1,455 | 727.5 | 34.02 |
| 137 | B34.8 Other viral infections of unspecified site | 4 | 75.47 | 0.001 | 491 | 245.5 | 53.97 |
| 143 | B97.2 Coronavirus as the cause of diseases classified elsewhere | 4 | 73.83 | 0.001 | 127 | 63.5 | 95.28 |
| 158 | B35-B49 Mycoses | 2 | 65.95 | 0.001 | 2,391 | 1,195.5 | 23.38 |
| 163 | B97.29 Other coronavirus as the cause of diseases classified elsewhere | 5 | 62.48 | 0.001 | 110 | 55.0 | 94.55 |
| 190 | B96 Other bacterial agents as the cause of diseases classified elsewhere | 3 | 54.32 | 0.001 | 1,513 | 756.5 | 26.64 |
| 200 | B97.1 Enterovirus as the cause of diseases classified elsewhere | 4 | 52.28 | 0.001 | 408 | 204.0 | 49.51 |
| 202 | A30-A49 Other bacterial diseases | 2 | 51.80 | 0.001 | 665 | 332.5 | 38.95 |
| 207 | A41 Other sepsis | 3 | 50.57 | 0.001 | 327 | 163.5 | 54.13 |
| 210 | A08.4 Viral intestinal infection, unspecified | 4 | 49.54 | 0.001 | 1,156 | 578.0 | 29.07 |
| 211 | B37 Candidiasis | 3 | 49.49 | 0.001 | 1,577 | 788.5 | 24.92 |
| 220 | B34.2 Coronavirus infection, unspecified | 4 | 45.75 | 0.001 | 66 | 33.0 | 100.00 |
| 232 | B97.10 Unspecified enterovirus as the cause of diseases classified elsewhere | 5 | 43.73 | 0.001 | 357 | 178.5 | 48.46 |
| 273 | B95 Streptococcus, Staphylococcus, and Enterococcus as the cause of diseases classified elsewhere | 3 | 34.55 | 0.001 | 425 | 212.5 | 39.76 |
| 321 | B96.89 Other specified bacterial agents as the cause of diseases classified elsewhere | 5 | 29.68 | 0.001 | 1,046 | 523.0 | 23.71 |
| 329 | B96.8 Other specified bacterial agents as the cause of diseases classified elsewhere | 4 | 29.21 | 0.001 | 1,114 | 557.0 | 22.80 |
| 337 | A04 Other bacterial intestinal infections | 3 | 28.81 | 0.001 | 345 | 172.5 | 40.29 |
| 348 | A04.7 Enterocolitis due to Clostridium difficile | 4 | 27.54 | 0.001 | 233 | 116.5 | 47.64 |
| 356 | A41.9 Sepsis, unspecified organism | 4 | 26.80 | 0.001 | 174 | 87.0 | 54.02 |
| 383 | B00-B09 Viral infections characterized by skin and mucous membrane lesions | 2 | 24.21 | 0.001 | 2,975 | 1,487.5 | 12.74 |
| 399 | A04.72 Enterocolitis due to Clostridium difficile, not specified as recurrent | 5 | 22.97 | 0.001 | 192 | 96.0 | 47.92 |
| 400 | B97.81 Human metapneumovirus as the cause of diseases classified elsewhere | 5 | 22.78 | 0.001 | 95 | 47.5 | 66.32 |
| 407 | B34.1 Enterovirus infection, unspecified | 4 | 22.54 | 0.001 | 225 | 112.5 | 44.00 |
| 429 | B37.2 Candidiasis of skin and nail | 4 | 21.42 | 0.001 | 850 | 425.0 | 22.35 |
| 445 | B97.0 Adenovirus as the cause of diseases classified elsewhere | 4 | 20.77 | 0.001 | 112 | 56.0 | 58.93 |
| 455 | B97.4 Respiratory syncytial virus as the cause of diseases classified elsewhere | 4 | 20.19 | 0.001 | 203 | 101.5 | 43.84 |
| 479 | B34.0 Adenovirus infection, unspecified | 4 | 18.84 | 0.001 | 122 | 61.0 | 54.10 |
| 493 | B37.0 Candidal stomatitis | 4 | 18.05 | 0.001 | 366 | 183.0 | 31.15 |
| 501 | A08.3 Other viral enteritis | 4 | 17.56 | 0.001 | 69 | 34.5 | 68.12 |
| 537 | B96.2 Escherichia coli [E. coli ] as the cause of diseases classified elsewhere | 4 | 15.53 | 0.001 | 227 | 113.5 | 36.56 |
| 556 | B27 Infectious mononucleosis | 3 | 14.79 | 0.001 | 285 | 142.5 | 31.93 |
| 567 | B95.6 Staphylococcus aureus as the cause of diseases classified elsewhere | 4 | 14.47 | 0.001 | 184 | 92.0 | 39.13 |
| 573 | A08.11 Acute gastroenteropathy due to Norwalk agent | 5 | 14.25 | 0.002 | 127 | 63.5 | 46.46 |
| 581 | A08.1 Acute gastroenteropathy due to Norwalk agent and other small round viruses | 4 | 14.01 | 0.002 | 129 | 64.5 | 45.74 |
| 601 | B96.20 Unspecified Escherichia coli [E. coli] as the cause of diseases classified elsewhere | 5 | 13.36 | 0.003 | 215 | 107.5 | 34.88 |
| 606 | B99-B99 Other infectious diseases | 2 | 13.23 | 0.003 | 217 | 108.5 | 34.56 |
| 607 | B99 Other and unspecified infectious diseases | 3 | 13.23 | 0.003 | 217 | 108.5 | 34.56 |
| 611 | B35 Dermatophytosis | 3 | 13.20 | 0.003 | 559 | 279.5 | 21.65 |
| 625 | B95.61 Methicillin susceptible Staphylococcus aureus infection as the cause of diseases classified elsewhere | 5 | 12.69 | 0.004 | 115 | 57.5 | 46.09 |
| 629 | B99.9 Unspecified infectious disease | 4 | 12.60 | 0.004 | 210 | 105.0 | 34.29 |
| 656 | A08.39 Other viral enteritis | 5 | 11.89 | 0.007 | 42 | 21.0 | 71.43 |
| 666 | B97.21 SARS-associated coronavirus as the cause of diseases classified elsewhere | 5 | 11.78 | 0.008 | 17 | 8.5 | 100.00 |
| 748 | B09 Unspecified viral infection characterized by skin and mucous membrane lesions | 3 | 10.13 | 0.034 | 762 | 381.0 | 16.27 |
| 756 | B08.5 Enteroviral vesicular pharyngitis | 4 | 10.04 | 0.034 | 227 | 113.5 | 29.52 |
| 770 | B27.00 Gammaherpesviral mononucleosis without complication | 5 | 9.77 | 0.046 | 147 | 73.5 | 36.05 |
| C00-D49 Neoplasms | | | | | | | |
| 70 | C00-D49 Neoplasms | 1 | 141.51 | 0.001 | 6,544 | 3,272.0 | 20.72 |
| 75 | C81-C96 Malignant neoplasms of lymphoid, hematopoietic and related tissue | 2 | 135.65 | 0.001 | 2,346 | 1,173.0 | 33.67 |
| 93 | C91 Lymphoid leukemia | 3 | 110.22 | 0.001 | 1,360 | 680.0 | 39.71 |
| 106 | C91.0 Acute lymphoblastic leukemia [ALL] | 4 | 98.99 | 0.001 | 1,266 | 633.0 | 39.02 |
| 189 | C76-C80 Malignant neoplasms of ill-defined, other secondary and unspecified sites | 2 | 54.88 | 0.001 | 611 | 305.5 | 41.73 |
| 213 | C91.00 Acute lymphoblastic leukemia not having achieved remission | 5 | 49.05 | 0.001 | 528 | 264.0 | 42.42 |
| 224 | C91.01 Acute lymphoblastic leukemia, in remission | 5 | 44.85 | 0.001 | 636 | 318.0 | 37.11 |
| 288 | C79 Secondary malignant neoplasm of other and unspecified sites | 3 | 33.16 | 0.001 | 337 | 168.5 | 43.62 |
| 317 | C95 Leukemia of unspecified cell type | 3 | 30.24 | 0.001 | 179 | 89.5 | 56.42 |
| 418 | C78 | 3 | 21.84 | 0.001 | 197 | 98.5 | 46.19 |
| 482 | C78.0 Secondary malignant neoplasm of lung | 4 | 18.71 | 0.001 | 186 | 93.0 | 44.09 |
| 531 | C95.0 Acute leukemia of unspecified cell type | 4 | 15.82 | 0.001 | 101 | 50.5 | 54.46 |
| 540 | C83 Non-follicular lymphoma | 3 | 15.42 | 0.001 | 138 | 69.0 | 46.38 |
| 565 | C95.9 Leukemia, unspecified | 4 | 14.49 | 0.001 | 78 | 39.0 | 58.97 |
| 637 | C79.5 | 4 | 12.33 | 0.005 | 155 | 77.5 | 39.35 |
| 674 | C95.90 Leukemia, unspecified not having achieved remission | 5 | 11.55 | 0.008 | 57 | 28.5 | 61.40 |
| 737 | C40-C41 Malignant neoplasms of bone and articular cartilage | 2 | 10.37 | 0.030 | 124 | 62.0 | 40.32 |
| D50-D89 Diseases of the blood and blood-forming organs and certain disorders involving the immune mechanism | | | | | | | |
| 10 | D50-D89 Diseases of the blood and blood-forming organs and certain disorders involving the immune mechanism | 1 | 550.78 | 0.001 | 9,904 | 4,952.0 | 33.04 |
| 51 | D80-D89 Certain disorders involving the immune mechanism | 2 | 179.80 | 0.001 | 2,151 | 1,075.5 | 40.31 |
| 61 | D84 Other immunodeficiencies | 3 | 158.66 | 0.001 | 1,439 | 719.5 | 46.07 |
| 76 | D84.8 Other specified immunodeficiencies | 4 | 132.88 | 0.001 | 956 | 478.0 | 51.46 |
| 81 | D70-D77 Other diseases of blood and blood-forming organs | 2 | 128.26 | 0.001 | 1,630 | 815.0 | 39.14 |
| 89 | D60-D64 Aplastic and other anaemias | 2 | 119.34 | 0.001 | 2,441 | 1,220.5 | 31.01 |
| 108 | D84.82 Immunodeficiency due to drugs and external causes | 5 | 97.69 | 0.001 | 780 | 390.0 | 48.97 |
| 118 | D84.821 Immunodeficiency due to drugs | 6 | 93.49 | 0.001 | 721 | 360.5 | 49.79 |
| 145 | D65-D69 Coagulation defects, purpura and other haemorrhagic conditions | 2 | 70.91 | 0.001 | 1,169 | 584.5 | 34.47 |
| 147 | D55-D59 Haemolytic anaemias | 2 | 70.08 | 0.001 | 1,217 | 608.5 | 33.61 |
| 165 | D57 Sickle-cell disorders | 3 | 62.01 | 0.001 | 915 | 457.5 | 36.39 |
| 179 | D61 Other aplastic anemias and other bone marrow failure syndromes | 3 | 57.95 | 0.001 | 696 | 348.0 | 40.23 |
| 186 | D72 Other disorders of white blood cells | 3 | 55.53 | 0.001 | 577 | 288.5 | 43.15 |
| 187 | D70 Neutropenia | 3 | 55.13 | 0.001 | 563 | 281.5 | 43.52 |
| 188 | D61.8 Other specified aplastic anemias and other bone marrow failure syndromes | 4 | 54.94 | 0.001 | 629 | 314.5 | 41.18 |
| 194 | D64 Other anemias | 3 | 52.98 | 0.001 | 1,468 | 734.0 | 26.70 |
| 198 | D69 Purpura and other hemorrhagic conditions | 3 | 52.50 | 0.001 | 712 | 356.0 | 37.92 |
| 201 | D61.81 Pancytopenia | 5 | 52.24 | 0.001 | 611 | 305.5 | 40.75 |
| 203 | D72.8 Other specified disorders of white blood cells | 4 | 51.50 | 0.001 | 540 | 270.0 | 42.96 |
| 225 | D57.0 Hb-SS disease with crisis | 4 | 44.82 | 0.001 | 397 | 198.5 | 46.60 |
| 256 | D61.810 Antineoplastic chemotherapy induced pancytopenia | 6 | 37.88 | 0.001 | 302 | 151.0 | 49.01 |
| 287 | D69.6 Thrombocytopenia, unspecified | 4 | 33.16 | 0.001 | 337 | 168.5 | 43.62 |
| 304 | D84.9 Immunodeficiency, unspecified | 4 | 31.17 | 0.001 | 474 | 237.0 | 35.86 |
| 310 | D68 Other coagulation defects | 3 | 30.81 | 0.001 | 333 | 166.5 | 42.34 |
| 313 | D70.9 Neutropenia, unspecified | 4 | 30.62 | 0.001 | 335 | 167.5 | 42.09 |
| 323 | D72.81 Decreased white blood cell count | 5 | 29.62 | 0.001 | 278 | 139.0 | 45.32 |
| 327 | D64.8 Other specified anemias | 4 | 29.22 | 0.001 | 308 | 154.0 | 42.86 |
| 330 | D64.9 Anemia, unspecified | 4 | 29.18 | 0.001 | 1,159 | 579.5 | 22.35 |
| 359 | D84.89 Other immunodeficiencies | 5 | 26.50 | 0.001 | 121 | 60.5 | 63.64 |
| 388 | D89.89 Other specified disorders involving the immune mechanism, not elsewhere classified | 5 | 23.97 | 0.001 | 81 | 40.5 | 72.84 |
| 396 | D89.8 Other specified disorders involving the immune mechanism, not elsewhere classified | 4 | 23.02 | 0.001 | 300 | 150.0 | 38.67 |
| 398 | D64.81 Anemia due to antineoplastic chemotherapy | 5 | 22.99 | 0.001 | 234 | 117.0 | 43.59 |
| 417 | D57.00 Hb-SS disease with crisis, unspecified | 5 | 21.96 | 0.001 | 249 | 124.5 | 41.37 |
| 430 | D72.82 Elevated white blood cell count | 5 | 21.39 | 0.001 | 260 | 130.0 | 40.00 |
| 441 | D72.829 Elevated white blood cell count, unspecified | 6 | 21.00 | 0.001 | 200 | 100.0 | 45.00 |
| 446 | D89 Other disorders involving the immune mechanism, not elsewhere classified | 3 | 20.72 | 0.001 | 421 | 210.5 | 31.12 |
| 453 | D69.5 Secondary thrombocytopenia | 4 | 20.19 | 0.001 | 221 | 110.5 | 42.08 |
| 454 | D69.59 Other secondary thrombocytopenia | 5 | 20.19 | 0.001 | 221 | 110.5 | 42.08 |
| 528 | D72.819 Decreased white blood cell count, unspecified | 6 | 15.91 | 0.001 | 155 | 77.5 | 44.52 |
| 551 | D61.818 Other pancytopenia | 6 | 14.91 | 0.001 | 295 | 147.5 | 31.53 |
| 554 | D50-D53 Nutritional anaemias | 2 | 14.90 | 0.001 | 1,294 | 647.0 | 15.15 |
| 587 | D75 Other and unspecified diseases of blood and blood-forming organs | 3 | 13.83 | 0.002 | 359 | 179.5 | 27.58 |
| 636 | D80.1 Nonfamilial hypogammaglobulinemia | 4 | 12.36 | 0.005 | 145 | 72.5 | 40.69 |
| 642 | D50 Iron deficiency anemia | 3 | 12.09 | 0.005 | 1,228 | 614.0 | 14.01 |
| 658 | D57.01 Hb-SS disease with acute chest syndrome | 5 | 11.84 | 0.007 | 75 | 37.5 | 54.67 |
| 681 | D57.2 Sickle-cell/Hb-C disease | 4 | 11.33 | 0.009 | 220 | 110.0 | 31.82 |
| 695 | D84.81 Immunodeficiency due to conditions classified elsewhere | 5 | 11.07 | 0.014 | 53 | 26.5 | 62.26 |
| 733 | D72.810 Lymphocytopenia | 6 | 10.47 | 0.019 | 109 | 54.5 | 43.12 |
| 747 | D75.8 Other specified diseases of blood and blood-forming organs | 4 | 10.14 | 0.033 | 158 | 79.0 | 35.44 |
| 751 | D50.9 Iron deficiency anemia, unspecified | 4 | 10.09 | 0.034 | 705 | 352.5 | 16.88 |
| 761 | D68.9 Coagulation defect, unspecified | 4 | 9.92 | 0.036 | 84 | 42.0 | 47.62 |
| 762 | D73.89 Other diseases of spleen | 5 | 9.91 | 0.037 | 35 | 17.5 | 71.43 |
| 763 | D73.8 Other diseases of spleen | 4 | 9.91 | 0.037 | 35 | 17.5 | 71.43 |
| 764 | D75.9 Disease of blood and blood-forming organs, unspecified | 4 | 9.89 | 0.043 | 115 | 57.5 | 40.87 |
| 769 | D80 Immunodeficiency with predominantly antibody defects | 3 | 9.84 | 0.045 | 205 | 102.5 | 30.73 |
| E00-E90 Endocrine, nutritional and metabolic diseases | | | | | | | |
| 16 | E00-E90 Endocrine, nutritional and metabolic diseases | 1 | 442.88 | 0.001 | 19,742 | 9,871.0 | 21.10 |
| 17 | E70-E88 Metabolic disorders | 2 | 366.68 | 0.001 | 7,912 | 3,956.0 | 30.21 |
| 46 | E87 Other disorders of fluid, electrolyte and acid-base balance | 3 | 194.26 | 0.001 | 1,767 | 883.5 | 46.01 |
| 78 | E86 Volume depletion | 3 | 130.74 | 0.001 | 1,720 | 860.0 | 38.49 |
| 86 | E86.0 Dehydration | 4 | 122.07 | 0.001 | 1,636 | 818.0 | 38.14 |
| 110 | E83 Disorders of mineral metabolism | 3 | 95.99 | 0.001 | 1,319 | 659.5 | 37.68 |
| 234 | E87.1 Hypo-osmolality and hyponatremia | 4 | 43.50 | 0.001 | 383 | 191.5 | 46.74 |
| 236 | E87.7 Fluid overload | 4 | 42.89 | 0.001 | 270 | 135.0 | 54.81 |
| 258 | E87.70 Fluid overload, unspecified | 5 | 37.60 | 0.001 | 241 | 120.5 | 54.36 |
| 260 | E83.4 Disorders of magnesium metabolism | 4 | 37.02 | 0.001 | 337 | 168.5 | 45.99 |
| 266 | E27 Other disorders of adrenal gland | 3 | 36.45 | 0.001 | 856 | 428.0 | 28.97 |
| 272 | E66 Overweight and obesity | 3 | 34.61 | 0.001 | 3,189 | 1,594.5 | 14.71 |
| 277 | E65-E68 Obesity and other hyperalimentation | 2 | 34.30 | 0.001 | 3,245 | 1,622.5 | 14.51 |
| 283 | E83.3 Disorders of phosphorus metabolism and phosphatases | 4 | 33.54 | 0.001 | 432 | 216.0 | 38.89 |
| 292 | E83.42 Hypomagnesemia | 5 | 32.48 | 0.001 | 317 | 158.5 | 44.48 |
| 305 | E83.39 Other disorders of phosphorus metabolism | 5 | 31.08 | 0.001 | 392 | 196.0 | 39.29 |
| 308 | E87.6 Hypokalemia | 4 | 31.01 | 0.001 | 331 | 165.5 | 42.60 |
| 333 | E20-E35 Disorders of other endocrine glands | 2 | 29.06 | 0.001 | 1,717 | 858.5 | 18.35 |
| 335 | E27.4 Other and unspecified adrenocortical insufficiency | 4 | 28.93 | 0.001 | 667 | 333.5 | 29.24 |
| 341 | E87.2 Acidosis | 4 | 28.51 | 0.001 | 306 | 153.0 | 42.48 |
| 375 | E40-E46 Malnutrition | 2 | 24.94 | 0.001 | 1,683 | 841.5 | 17.17 |
| 377 | E66.9 Obesity, unspecified | 4 | 24.87 | 0.001 | 1,235 | 617.5 | 20.00 |
| 389 | E27.40 Unspecified adrenocortical insufficiency | 5 | 23.78 | 0.001 | 432 | 216.0 | 32.87 |
| 392 | E87.8 Other disorders of electrolyte and fluid balance, not elsewhere classified | 4 | 23.37 | 0.001 | 193 | 96.5 | 48.19 |
| 411 | E88.0 Disorders of plasma-protein metabolism, not elsewhere classified | 4 | 22.34 | 0.001 | 177 | 88.5 | 49.15 |
| 416 | E08-E14 Diabetes mellitus | 2 | 22.05 | 0.001 | 2,145 | 1,072.5 | 14.31 |
| 420 | E88.09 Other disorders of plasma-protein metabolism, not elsewhere classified | 5 | 21.74 | 0.001 | 162 | 81.0 | 50.62 |
| 495 | E50-E64 Other nutritional deficiencies | 2 | 17.88 | 0.001 | 2,044 | 1,022.0 | 13.21 |
| 517 | E88 Other and unspecified metabolic disorders | 3 | 16.94 | 0.001 | 661 | 330.5 | 22.54 |
| 539 | E43 Unspecified severe protein-calorie malnutrition | 3 | 15.44 | 0.001 | 658 | 329.0 | 21.58 |
| 602 | E83.5 Disorders of calcium metabolism | 4 | 13.33 | 0.003 | 343 | 171.5 | 27.70 |
| 621 | E66.0 Obesity due to excess calories | 4 | 12.91 | 0.004 | 1,931 | 965.5 | 11.55 |
| 650 | E87.0 Hyperosmolality and hypernatremia | 4 | 11.92 | 0.007 | 89 | 44.5 | 50.56 |
| 675 | E10 Type 1 diabetes mellitus | 3 | 11.47 | 0.009 | 1,512 | 756.0 | 12.30 |
| 678 | E83.51 Hypocalcemia | 5 | 11.37 | 0.009 | 184 | 92.0 | 34.78 |
| 722 | E66.09 Other obesity due to excess calories | 5 | 10.57 | 0.017 | 640 | 320.0 | 18.12 |
| F00-F99 Mental and behavioural disorders | | | | | | | |
| 64 | F00-F99 Mental and behavioural disorders | 1 | 151.34 | 0.001 | 33,334 | 16,667.0 | 9.52 |
| 73 | F80-F89 Disorders of psychological development | 2 | 137.66 | 0.001 | 10,942 | 5,471.0 | 15.83 |
| 169 | F88 Other disorders of psychological development | 3 | 61.25 | 0.001 | 3,240 | 1,620.0 | 19.38 |
| 221 | F80 Specific developmental disorders of speech and language | 3 | 45.56 | 0.001 | 3,953 | 1,976.5 | 15.15 |
| 257 | F41.8 Other specified anxiety disorders | 4 | 37.66 | 0.001 | 487 | 243.5 | 38.81 |
| 275 | F40-F48 Neurotic, stress-related and somatoform disorders | 2 | 34.46 | 0.001 | 8,750 | 4,375.0 | 8.87 |
| 291 | F82 Specific developmental disorder of motor function | 3 | 32.54 | 0.001 | 2,092 | 1,046.0 | 17.59 |
| 302 | F80.9 Developmental disorder of speech and language, unspecified | 4 | 31.42 | 0.001 | 1,775 | 887.5 | 18.76 |
| 338 | F41 Other anxiety disorders | 3 | 28.79 | 0.001 | 3,912 | 1,956.0 | 12.12 |
| 541 | F70-F79 Mental retardation | 2 | 15.40 | 0.001 | 1,278 | 639.0 | 15.49 |
| 699 | F72 Severe intellectual disabilities | 3 | 11.02 | 0.014 | 146 | 73.0 | 38.36 |
| 752 | F80.2 Mixed receptive-expressive language disorder | 4 | 10.08 | 0.034 | 842 | 421.0 | 15.44 |
| G00-G99 Diseases of the nervous system | | | | | | | |
| 21 | G00-G99 Diseases of the nervous system | 1 | 338.38 | 0.001 | 28,499 | 14,249.5 | 15.38 |
| 43 | G40-G47 Episodic and paroxysmal disorders | 2 | 203.77 | 0.001 | 20,125 | 10,062.5 | 14.21 |
| 104 | G89-G99 Other disorders of the nervous system | 2 | 103.45 | 0.001 | 5,455 | 2,727.5 | 19.41 |
| 117 | G40 Epilepsy and recurrent seizures | 3 | 94.93 | 0.001 | 6,889 | 3,444.5 | 16.56 |
| 139 | G47 Sleep disorders | 3 | 74.81 | 0.001 | 8,757 | 4,378.5 | 13.05 |
| 180 | G47.3 Sleep apnea | 4 | 57.60 | 0.001 | 5,086 | 2,543.0 | 15.02 |
| 182 | G93 Other disorders of brain | 3 | 56.48 | 0.001 | 1,529 | 764.5 | 27.01 |
| 185 | G40.9 Epilepsy, unspecified | 4 | 55.57 | 0.001 | 2,135 | 1,067.5 | 22.72 |
| 227 | G40.8 Other epilepsy and recurrent seizures | 4 | 44.61 | 0.001 | 1,243 | 621.5 | 26.63 |
| 238 | G40.90 Epilepsy, unspecified, not intractable | 5 | 42.59 | 0.001 | 1,674 | 837.0 | 22.46 |
| 251 | G89.1 Acute pain, not elsewhere classified | 4 | 38.51 | 0.001 | 630 | 315.0 | 34.60 |
| 253 | G89.18 Other acute postprocedural pain | 5 | 38.13 | 0.001 | 619 | 309.5 | 34.73 |
| 286 | G89 Pain, not elsewhere classified | 3 | 33.35 | 0.001 | 3,184 | 1,592.0 | 14.45 |
| 296 | G93.4 Other and unspecified encephalopathy | 4 | 31.72 | 0.001 | 527 | 263.5 | 34.35 |
| 318 | G40.909 Epilepsy, unspecified, not intractable, without status epilepticus | 6 | 30.23 | 0.001 | 1,469 | 734.5 | 20.22 |
| 342 | G47.33 Obstructive sleep apnea (adult) (pediatric) | 5 | 28.30 | 0.001 | 2,922 | 1,461.0 | 13.89 |
| 347 | G43 Migraine | 3 | 27.57 | 0.001 | 3,473 | 1,736.5 | 12.58 |
| 382 | G93.8 Other specified disorders of brain | 4 | 24.25 | 0.001 | 597 | 298.5 | 28.31 |
| 386 | G93.89 Other specified disorders of brain | 5 | 24.01 | 0.001 | 575 | 287.5 | 28.70 |
| 468 | G93.49 Other encephalopathy | 5 | 19.42 | 0.001 | 347 | 173.5 | 33.14 |
| 472 | G62 Other and unspecified polyneuropathies | 3 | 19.18 | 0.001 | 213 | 106.5 | 41.78 |
| 505 | G60-G65 Polyneuropathies and other disorders of the peripheral nervous system | 2 | 17.45 | 0.001 | 270 | 135.0 | 35.56 |
| 512 | G47.30 Sleep apnea, unspecified | 5 | 17.17 | 0.001 | 1,641 | 820.5 | 14.44 |
| 523 | G20-G26 Extrapyramidal and movement disorders | 2 | 16.14 | 0.001 | 521 | 260.5 | 24.76 |
| 526 | G40.82 Epileptic spasms | 5 | 15.99 | 0.001 | 253 | 126.5 | 35.18 |
| 534 | G40.901 Epilepsy, unspecified, not intractable, with status epilepticus | 6 | 15.62 | 0.001 | 205 | 102.5 | 38.54 |
| 615 | G43.0 Migraine without aura | 4 | 13.03 | 0.003 | 1,493 | 746.5 | 13.19 |
| 617 | G40.91 Epilepsy, unspecified, intractable | 5 | 13.01 | 0.003 | 461 | 230.5 | 23.64 |
| 623 | G40.80 Other epilepsy | 5 | 12.82 | 0.004 | 410 | 205.0 | 24.88 |
| 659 | G43.9 Migraine, unspecified | 4 | 11.84 | 0.007 | 592 | 296.0 | 19.93 |
| 672 | G97 Intraoperative and postprocedural complications and disorders of nervous system, not elsewhere classified | 3 | 11.56 | 0.008 | 54 | 27.0 | 62.96 |
| 688 | G43.90 Migraine, unspecified, not intractable | 5 | 11.20 | 0.011 | 544 | 272.0 | 20.22 |
| 704 | G47.9 Sleep disorder, unspecified | 4 | 10.91 | 0.014 | 1,505 | 752.5 | 12.03 |
| 705 | G80-G88 Cerebral palsy and other paralytic syndromes | 2 | 10.89 | 0.014 | 1,023 | 511.5 | 14.57 |
| 721 | G43.909 Migraine, unspecified, not intractable, without status migrainosus | 6 | 10.58 | 0.016 | 476 | 238.0 | 21.01 |
| 731 | G40.2 Localization-related (focal) (partial) symptomatic epilepsy and epileptic syndromes with complex partial seizures | 4 | 10.51 | 0.018 | 795 | 397.5 | 16.23 |
| 760 | G90 Disorders of autonomic nervous system | 3 | 9.94 | 0.036 | 203 | 101.5 | 31.03 |
| 766 | G93.40 Encephalopathy, unspecified | 5 | 9.88 | 0.044 | 162 | 81.0 | 34.57 |
| H00-H59 Diseases of the eye and adnexa | | | | | | | |
| 74 | H00-H59 Diseases of the eye and adnexa | 1 | 136.05 | 0.001 | 25,004 | 12,502.0 | 10.42 |
| 255 | H10-H13 Disorders of conjunctiva | 2 | 38.06 | 0.001 | 4,147 | 2,073.5 | 13.53 |
| 268 | H10 Conjunctivitis | 3 | 36.09 | 0.001 | 3,992 | 1,996.0 | 13.43 |
| 309 | H49-H52 Disorders of ocular muscles, binocular movement, accommodation and refraction | 2 | 30.90 | 0.001 | 10,678 | 5,339.0 | 7.60 |
| 380 | H52 Disorders of refraction and accommodation | 3 | 24.48 | 0.001 | 6,974 | 3,487.0 | 8.37 |
| 394 | H55-H59 Other disorders of eye and adnexa | 2 | 23.18 | 0.001 | 1,477 | 738.5 | 17.67 |
| 458 | H00-H06 Disorders of eyelid, lacrimal system and orbit | 2 | 20.00 | 0.001 | 2,348 | 1,174.0 | 13.03 |
| 497 | H10.3 Unspecified acute conjunctivitis | 4 | 17.80 | 0.001 | 1,531 | 765.5 | 15.22 |
| 506 | H53-H54 Visual disturbances and blindness | 2 | 17.36 | 0.001 | 3,473 | 1,736.5 | 9.99 |
| 530 | H57 Other disorders of eye and adnexa | 3 | 15.82 | 0.001 | 962 | 481.0 | 18.09 |
| 564 | H53 Visual disturbances | 3 | 14.50 | 0.001 | 3,068 | 1,534.0 | 9.71 |
| 596 | H01.00 Unspecified blepharitis | 5 | 13.45 | 0.003 | 117 | 58.5 | 47.01 |
| 597 | H57.89 Other specified disorders of eye and adnexa | 5 | 13.42 | 0.003 | 472 | 236.0 | 23.73 |
| 620 | H57.8 Other specified disorders of eye and adnexa | 4 | 12.92 | 0.004 | 481 | 240.5 | 23.08 |
| 702 | H10.1 Acute atopic conjunctivitis | 4 | 10.97 | 0.014 | 1,142 | 571.0 | 13.84 |
| 707 | H52.2 Astigmatism | 4 | 10.82 | 0.014 | 3,176 | 1,588.0 | 8.25 |
| 712 | H53.8 Other visual disturbances | 4 | 10.78 | 0.014 | 371 | 185.5 | 23.99 |
| 727 | H02 Other disorders of eyelid | 3 | 10.54 | 0.017 | 793 | 396.5 | 16.27 |
| 729 | H46-H48 Disorders of optic nerve and visual pathways | 2 | 10.52 | 0.018 | 962 | 481.0 | 14.76 |
| 739 | H47 Other disorders of optic [2nd] nerve and visual pathways | 3 | 10.35 | 0.030 | 950 | 475.0 | 14.74 |
| H60-H95 Diseases of the ear and mastoid process | | | | | | | |
| 27 | H60-H95 Diseases of the ear and mastoid process | 1 | 285.90 | 0.001 | 34,961 | 17,480.5 | 12.77 |
| 35 | H65-H75 Diseases of middle ear and mastoid | 2 | 232.82 | 0.001 | 24,309 | 12,154.5 | 13.82 |
| 47 | H66 Suppurative and unspecified otitis media | 3 | 193.90 | 0.001 | 15,166 | 7,583.0 | 15.96 |
| 71 | H66.0 Acute suppurative otitis media | 4 | 140.36 | 0.001 | 7,139 | 3,569.5 | 19.76 |
| 72 | H66.00 Acute suppurative otitis media without spontaneous rupture of ear drum | 5 | 138.16 | 0.001 | 6,918 | 3,459.0 | 19.92 |
| 116 | H65 Nonsuppurative otitis media | 3 | 95.02 | 0.001 | 4,993 | 2,496.5 | 19.45 |
| 171 | H66.9 Otitis media, unspecified | 4 | 60.89 | 0.001 | 7,810 | 3,905.0 | 12.47 |
| 196 | H66.001 Acute suppurative otitis media without spontaneous rupture of ear drum, right ear | 6 | 52.60 | 0.001 | 2,356 | 1,178.0 | 21.05 |
| 243 | H66.002 Acute suppurative otitis media without spontaneous rupture of ear drum, left ear | 6 | 41.01 | 0.001 | 1,897 | 948.5 | 20.72 |
| 246 | H66.92 Otitis media, unspecified, left ear | 5 | 40.14 | 0.001 | 1,127 | 563.5 | 26.53 |
| 250 | H66.91 Otitis media, unspecified, right ear | 5 | 38.62 | 0.001 | 1,372 | 686.0 | 23.62 |
| 294 | H65.0 Acute serous otitis media | 4 | 31.98 | 0.001 | 1,254 | 627.0 | 22.49 |
| 297 | H92.0 Otalgia | 4 | 31.71 | 0.001 | 1,964 | 982.0 | 17.92 |
| 325 | H60-H62 Diseases of external ear | 2 | 29.47 | 0.001 | 3,161 | 1,580.5 | 13.63 |
| 336 | H90-H94 Other disorders of ear | 2 | 28.82 | 0.001 | 7,407 | 3,703.5 | 8.82 |
| 360 | H66.90 Otitis media, unspecified, unspecified ear | 5 | 26.28 | 0.001 | 989 | 494.5 | 22.95 |
| 397 | H65.1 Other acute nonsuppurative otitis media | 4 | 23.02 | 0.001 | 696 | 348.0 | 25.57 |
| 408 | H61.2 Impacted cerumen | 4 | 22.50 | 0.001 | 2,185 | 1,092.5 | 14.32 |
| 409 | H65.9 Unspecified nonsuppurative otitis media | 4 | 22.49 | 0.001 | 1,628 | 814.0 | 16.58 |
| 412 | H91 Other and unspecified hearing loss | 3 | 22.29 | 0.001 | 1,855 | 927.5 | 15.47 |
| 434 | H61 Other disorders of external ear | 3 | 21.25 | 0.001 | 2,268 | 1,134.0 | 13.67 |
| 456 | H66.003 Acute suppurative otitis media without spontaneous rupture of ear drum, bilateral | 6 | 20.11 | 0.001 | 1,475 | 737.5 | 16.47 |
| 477 | H92 Otalgia and effusion of ear | 3 | 18.93 | 0.001 | 2,831 | 1,415.5 | 11.55 |
| 478 | H65.19 Other acute nonsuppurative otitis media | 5 | 18.85 | 0.001 | 611 | 305.5 | 24.71 |
| 496 | H65.3 Chronic mucoid otitis media | 4 | 17.81 | 0.001 | 254 | 127.0 | 37.01 |
| 503 | H91.9 Unspecified hearing loss | 4 | 17.51 | 0.001 | 1,734 | 867.0 | 14.19 |
| 608 | H65.33 Chronic mucoid otitis media, bilateral | 5 | 13.23 | 0.003 | 195 | 97.5 | 36.41 |
| 619 | H73.8 Other specified disorders of tympanic membrane | 4 | 12.93 | 0.004 | 139 | 69.5 | 42.45 |
| 643 | H91.90 Unspecified hearing loss, unspecified ear | 5 | 12.07 | 0.005 | 1,051 | 525.5 | 15.13 |
| 673 | H73 Other disorders of tympanic membrane | 3 | 11.55 | 0.008 | 204 | 102.0 | 33.33 |
| 689 | H73.89 Other specified disorders of tympanic membrane | 5 | 11.20 | 0.011 | 134 | 67.0 | 40.30 |
| 717 | H61.23 Impacted cerumen, bilateral | 5 | 10.72 | 0.014 | 1,243 | 621.5 | 13.11 |
| 730 | H66.004 Acute suppurative otitis media without spontaneous rupture of ear drum, recurrent, right ear | 6 | 10.51 | 0.018 | 271 | 135.5 | 27.68 |
| 742 | H92.01 Otalgia, right ear | 5 | 10.24 | 0.031 | 594 | 297.0 | 18.52 |
| I00-I99 Diseases of the circulatory system | | | | | | | |
| 20 | I00-I99 Diseases of the circulatory system | 1 | 340.06 | 0.001 | 9,019 | 4,509.5 | 27.29 |
| 103 | I30-I52 Other forms of heart disease | 2 | 104.20 | 0.001 | 3,060 | 1,530.0 | 25.95 |
| 132 | I10-I19 Hypertensive diseases | 2 | 77.06 | 0.001 | 1,648 | 824.0 | 30.34 |
| 191 | I10 Essential (primary) hypertension | 3 | 53.77 | 0.001 | 1,250 | 625.0 | 29.12 |
| 241 | I80-I89 Diseases of veins, lymphatic vessels and lymph nodes, not elsewhere classified | 2 | 41.38 | 0.001 | 1,190 | 595.0 | 26.22 |
| 271 | I82 Other venous embolism and thrombosis | 3 | 35.10 | 0.001 | 695 | 347.5 | 31.51 |
| 316 | I60-I69 Cerebrovascular diseases | 2 | 30.30 | 0.001 | 780 | 390.0 | 27.69 |
| 328 | I26-I28 Pulmonary heart disease and diseases of pulmonary circulation | 2 | 29.22 | 0.001 | 729 | 364.5 | 28.12 |
| 334 | I95.9 Hypotension, unspecified | 4 | 29.04 | 0.001 | 207 | 103.5 | 51.69 |
| 355 | I50 Heart failure | 3 | 26.83 | 0.001 | 291 | 145.5 | 42.27 |
| 361 | I77 Other disorders of arteries and arterioles | 3 | 26.21 | 0.001 | 479 | 239.5 | 32.78 |
| 367 | I27 Other pulmonary heart diseases | 3 | 25.26 | 0.001 | 554 | 277.0 | 29.96 |
| 379 | I27.2 Other secondary pulmonary hypertension | 4 | 24.56 | 0.001 | 523 | 261.5 | 30.40 |
| 402 | I77.8 Other specified disorders of arteries and arterioles | 4 | 22.64 | 0.001 | 416 | 208.0 | 32.69 |
| 415 | I51 Complications and ill-defined descriptions of heart disease | 3 | 22.11 | 0.001 | 579 | 289.5 | 27.46 |
| 427 | I70-I79 Diseases of arteries, arterioles and capillaries | 2 | 21.42 | 0.001 | 755 | 377.5 | 23.71 |
| 431 | I95-I99 Other and unspecified disorders of the circulatory system | 2 | 21.39 | 0.001 | 613 | 306.5 | 26.26 |
| 448 | I77.81 Aortic ectasia | 5 | 20.61 | 0.001 | 368 | 184.0 | 33.15 |
| 481 | I95 Hypotension | 3 | 18.71 | 0.001 | 444 | 222.0 | 28.83 |
| 515 | I77.810 Thoracic aortic ectasia | 6 | 17.05 | 0.001 | 329 | 164.5 | 31.91 |
| 525 | I27.20 Pulmonary hypertension, unspecified | 5 | 15.99 | 0.001 | 426 | 213.0 | 27.23 |
| 560 | I69 Sequelae of cerebrovascular disease | 3 | 14.66 | 0.001 | 263 | 131.5 | 33.08 |
| 562 | I25 Chronic ischemic heart disease | 3 | 14.53 | 0.001 | 109 | 54.5 | 50.46 |
| 570 | I20-I25 Ischaemic heart diseases | 2 | 14.39 | 0.001 | 130 | 65.0 | 46.15 |
| 580 | I50.8 Other heart failure | 4 | 14.02 | 0.002 | 77 | 38.5 | 58.44 |
| 583 | I15 Secondary hypertension | 3 | 13.93 | 0.002 | 258 | 129.0 | 32.56 |
| 588 | I63.5 Cerebral infarction due to unspecified occlusion or stenosis of cerebral arteries | 4 | 13.78 | 0.003 | 42 | 21.0 | 76.19 |
| 614 | I36 Nonrheumatic tricuspid valve disorders | 3 | 13.05 | 0.003 | 147 | 73.5 | 41.50 |
| 655 | I15.8 Other secondary hypertension | 4 | 11.89 | 0.007 | 105 | 52.5 | 46.67 |
| 690 | I36.1 Nonrheumatic tricuspid (valve) insufficiency | 4 | 11.18 | 0.011 | 139 | 69.5 | 39.57 |
| J00-J99 Diseases of the respiratory system | | | | | | | |
| 6 | J00-J99 Diseases of the respiratory system | 1 | 1,883.66 | 0.001 | 64,279 | 32,139.5 | 24.09 |
| 11 | J00-J06 Acute upper respiratory infections | 2 | 545.34 | 0.001 | 19,956 | 9,978.0 | 23.27 |
| 14 | J40-J47 Chronic lower respiratory diseases | 2 | 459.57 | 0.001 | 18,532 | 9,266.0 | 22.18 |
| 15 | J45 Asthma | 3 | 452.15 | 0.001 | 18,209 | 9,104.5 | 22.19 |
| 25 | J09-J18 Influenza and pneumonia | 2 | 294.50 | 0.001 | 3,758 | 1,879.0 | 39.06 |
| 26 | J96-J99 Other diseases of the respiratory system | 2 | 288.99 | 0.001 | 4,762 | 2,381.0 | 34.48 |
| 41 | J45.9 Other and unspecified asthma | 4 | 211.84 | 0.001 | 4,360 | 2,180.0 | 30.92 |
| 42 | J45.90 Unspecified asthma | 5 | 208.14 | 0.001 | 3,908 | 1,954.0 | 32.34 |
| 44 | J30-J39 Other diseases of upper respiratory tract | 2 | 202.85 | 0.001 | 13,553 | 6,776.5 | 17.26 |
| 48 | J06.9 Acute upper respiratory infection, unspecified | 4 | 189.95 | 0.001 | 9,092 | 4,546.0 | 20.37 |
| 49 | J06 Acute upper respiratory infections of multiple and unspecified sites | 3 | 189.64 | 0.001 | 9,097 | 4,548.5 | 20.35 |
| 59 | J96 Respiratory failure, not elsewhere classified | 3 | 162.87 | 0.001 | 2,299 | 1,149.5 | 37.19 |
| 65 | J02 Acute pharyngitis | 3 | 149.80 | 0.001 | 5,141 | 2,570.5 | 24.02 |
| 68 | J45.909 Unspecified asthma, uncomplicated | 6 | 147.12 | 0.001 | 2,790 | 1,395.0 | 32.19 |
| 84 | J98 Other respiratory disorders | 3 | 126.26 | 0.001 | 2,421 | 1,210.5 | 32.01 |
| 88 | J02.9 Acute pharyngitis, unspecified | 4 | 120.37 | 0.001 | 4,303 | 2,151.5 | 23.54 |
| 90 | J20-J22 Other acute lower respiratory infections | 2 | 118.98 | 0.001 | 2,304 | 1,152.0 | 31.86 |
| 102 | J96.0 Acute respiratory failure | 4 | 104.57 | 0.001 | 891 | 445.5 | 47.47 |
| 113 | J45.2 Mild intermittent asthma | 4 | 95.41 | 0.001 | 5,678 | 2,839.0 | 18.28 |
| 114 | J21 Acute bronchiolitis | 3 | 95.25 | 0.001 | 1,998 | 999.0 | 30.63 |
| 120 | J10 Influenza due to other identified influenza virus | 3 | 89.53 | 0.001 | 1,087 | 543.5 | 40.02 |
| 123 | J45.3 Mild persistent asthma | 4 | 85.98 | 0.001 | 4,461 | 2,230.5 | 19.57 |
| 126 | J10.1 Influenza due to other identified influenza virus with other respiratory manifestations | 4 | 83.38 | 0.001 | 1,046 | 523.0 | 39.39 |
| 131 | J00 Acute nasopharyngitis [common cold] | 3 | 78.34 | 0.001 | 2,042 | 1,021.0 | 27.52 |
| 133 | J12 Viral pneumonia, not elsewhere classified | 3 | 76.23 | 0.001 | 436 | 218.0 | 57.34 |
| 146 | J05 Acute obstructive laryngitis [croup] and epiglottitis | 3 | 70.66 | 0.001 | 1,915 | 957.5 | 27.00 |
| 148 | J05.0 Acute obstructive laryngitis [croup] | 4 | 69.95 | 0.001 | 1,912 | 956.0 | 26.88 |
| 156 | J96.01 Acute respiratory failure with hypoxia | 5 | 67.17 | 0.001 | 526 | 263.0 | 49.43 |
| 160 | J34 Other and unspecified disorders of nose and nasal sinuses | 3 | 64.19 | 0.001 | 2,276 | 1,138.0 | 23.64 |
| 167 | J45.4 Moderate persistent asthma | 4 | 61.43 | 0.001 | 3,139 | 1,569.5 | 19.72 |
| 170 | J11.1 Influenza due to unidentified influenza virus with other respiratory manifestations | 4 | 61.17 | 0.001 | 791 | 395.5 | 38.81 |
| 172 | J12.8 Other viral pneumonia | 4 | 60.53 | 0.001 | 159 | 79.5 | 81.13 |
| 173 | J11 Influenza due to unidentified influenza virus | 3 | 59.88 | 0.001 | 802 | 401.0 | 38.15 |
| 175 | J21.9 Acute bronchiolitis, unspecified | 4 | 59.45 | 0.001 | 1,286 | 643.0 | 30.17 |
| 176 | J12.82 Pneumonia due to coronavirus disease 2019 | 5 | 58.92 | 0.001 | 85 | 42.5 | 100.00 |
| 177 | J45.30 Mild persistent asthma, uncomplicated | 5 | 58.51 | 0.001 | 3,390 | 1,695.0 | 18.53 |
| 183 | J45.20 Mild intermittent asthma, uncomplicated | 5 | 56.43 | 0.001 | 4,020 | 2,010.0 | 16.72 |
| 212 | J34.8 Other specified disorders of nose and nasal sinuses | 4 | 49.21 | 0.001 | 1,826 | 913.0 | 23.11 |
| 214 | J34.89 Other specified disorders of nose and nasal sinuses | 5 | 49.00 | 0.001 | 1,825 | 912.5 | 23.07 |
| 218 | J18 Pneumonia, unspecified organism | 3 | 46.66 | 0.001 | 923 | 461.5 | 31.53 |
| 222 | J45.901 Unspecified asthma with (acute) exacerbation | 6 | 45.41 | 0.001 | 954 | 477.0 | 30.61 |
| 223 | J98.11 Atelectasis | 5 | 44.87 | 0.001 | 488 | 244.0 | 42.21 |
| 226 | J35 Chronic diseases of tonsils and adenoids | 3 | 44.68 | 0.001 | 2,251 | 1,125.5 | 19.86 |
| 231 | J98.1 Pulmonary collapse | 4 | 44.04 | 0.001 | 506 | 253.0 | 41.11 |
| 237 | J98.8 Other specified respiratory disorders | 4 | 42.77 | 0.001 | 854 | 427.0 | 31.38 |
| 240 | J01 Acute sinusitis | 3 | 41.44 | 0.001 | 1,211 | 605.5 | 26.01 |
| 245 | J30 Vasomotor and allergic rhinitis | 3 | 40.59 | 0.001 | 6,084 | 3,042.0 | 11.54 |
| 259 | J18.9 Pneumonia, unspecified organism | 4 | 37.53 | 0.001 | 780 | 390.0 | 30.77 |
| 261 | J45.21 Mild intermittent asthma with (acute) exacerbation | 5 | 36.96 | 0.001 | 1,567 | 783.5 | 21.63 |
| 263 | J45.40 Moderate persistent asthma, uncomplicated | 5 | 36.60 | 0.001 | 2,231 | 1,115.5 | 18.06 |
| 265 | J80-J84 Other respiratory diseases principally affecting the interstitium | 2 | 36.51 | 0.001 | 277 | 138.5 | 50.18 |
| 270 | J90-J94 Other diseases of pleura | 2 | 35.89 | 0.001 | 531 | 265.5 | 36.35 |
| 274 | J96.2 Acute and chronic respiratory failure | 4 | 34.46 | 0.001 | 487 | 243.5 | 37.17 |
| 345 | J90 Pleural effusion, not elsewhere classified | 3 | 28.00 | 0.001 | 302 | 151.0 | 42.38 |
| 357 | J15 Bacterial pneumonia, not elsewhere classified | 3 | 26.57 | 0.001 | 410 | 205.0 | 35.61 |
| 362 | J45.31 Mild persistent asthma with (acute) exacerbation | 5 | 26.14 | 0.001 | 977 | 488.5 | 23.03 |
| 365 | J98.0 Diseases of bronchus, not elsewhere classified | 4 | 26.00 | 0.001 | 413 | 206.5 | 35.11 |
| 371 | J95-J95 Intraoperative and postprocedural complications and disorders of respiratory system, not elsewhere classified | 2 | 25.02 | 0.001 | 417 | 208.5 | 34.29 |
| 372 | J95 Intraoperative and postprocedural complications and disorders of respiratory system, not elsewhere classified | 3 | 25.02 | 0.001 | 417 | 208.5 | 34.29 |
| 373 | J38 Diseases of vocal cords and larynx, not elsewhere classified | 3 | 24.99 | 0.001 | 746 | 373.0 | 25.74 |
| 381 | J02.0 Streptococcal pharyngitis | 4 | 24.42 | 0.001 | 607 | 303.5 | 28.17 |
| 391 | J45.5 Severe persistent asthma | 4 | 23.59 | 0.001 | 571 | 285.5 | 28.55 |
| 395 | J96.9 Respiratory failure, unspecified | 4 | 23.11 | 0.001 | 195 | 97.5 | 47.69 |
| 413 | J35.2 Hypertrophy of adenoids | 4 | 22.14 | 0.001 | 600 | 300.0 | 27.00 |
| 414 | J01.9 Acute sinusitis, unspecified | 4 | 22.14 | 0.001 | 660 | 330.0 | 25.76 |
| 421 | J81 Pulmonary edema | 3 | 21.70 | 0.001 | 114 | 57.0 | 59.65 |
| 424 | J01.90 Acute sinusitis, unspecified | 5 | 21.62 | 0.001 | 637 | 318.5 | 25.90 |
| 426 | J32 Chronic sinusitis | 3 | 21.48 | 0.001 | 830 | 415.0 | 22.65 |
| 432 | J96.00 Acute respiratory failure, unspecified whether with hypoxia or hypercapnia | 5 | 21.29 | 0.001 | 242 | 121.0 | 41.32 |
| 440 | J30.2 Other seasonal allergic rhinitis | 4 | 21.02 | 0.001 | 1,566 | 783.0 | 16.35 |
| 447 | J45.41 Moderate persistent asthma with (acute) exacerbation | 5 | 20.70 | 0.001 | 781 | 390.5 | 22.92 |
| 459 | J96.21 Acute and chronic respiratory failure with hypoxia | 5 | 19.90 | 0.001 | 253 | 126.5 | 39.13 |
| 461 | J95.8 Other intraoperative and postprocedural complications and disorders of respiratory system, not elsewhere classified | 4 | 19.78 | 0.001 | 230 | 115.0 | 40.87 |
| 475 | J39 Other diseases of upper respiratory tract | 3 | 19.07 | 0.001 | 290 | 145.0 | 35.86 |
| 476 | J22 Unspecified acute lower respiratory infection | 3 | 19.03 | 0.001 | 128 | 64.0 | 53.12 |
| 484 | J21.0 Acute bronchiolitis due to respiratory syncytial virus | 4 | 18.58 | 0.001 | 356 | 178.0 | 32.02 |
| 489 | J39.8 Other specified diseases of upper respiratory tract | 4 | 18.28 | 0.001 | 218 | 109.0 | 40.37 |
| 492 | J98.09 Other diseases of bronchus, not elsewhere classified | 5 | 18.10 | 0.001 | 245 | 122.5 | 37.96 |
| 494 | J21.8 Acute bronchiolitis due to other specified organisms | 4 | 18.00 | 0.001 | 330 | 165.0 | 32.73 |
| 502 | J32.9 Chronic sinusitis, unspecified | 4 | 17.53 | 0.001 | 727 | 363.5 | 21.87 |
| 507 | J96.1 Chronic respiratory failure | 4 | 17.33 | 0.001 | 726 | 363.0 | 21.76 |
| 508 | J45.902 Unspecified asthma with status asthmaticus | 6 | 17.31 | 0.001 | 164 | 82.0 | 45.12 |
| 510 | J98.4 Other disorders of lung | 4 | 17.21 | 0.001 | 320 | 160.0 | 32.50 |
| 511 | J15.9 Unspecified bacterial pneumonia | 4 | 17.18 | 0.001 | 291 | 145.5 | 34.02 |
| 518 | J96.02 Acute respiratory failure with hypercapnia | 5 | 16.93 | 0.001 | 123 | 61.5 | 51.22 |
| 529 | J30.9 Allergic rhinitis, unspecified | 4 | 15.88 | 0.001 | 1,685 | 842.5 | 13.71 |
| 589 | J03 Acute tonsillitis | 3 | 13.68 | 0.003 | 370 | 185.0 | 27.03 |
| 618 | J12.3 Human metapneumovirus pneumonia | 4 | 13.00 | 0.003 | 60 | 30.0 | 63.33 |
| 628 | J12.89 Other viral pneumonia | 5 | 12.60 | 0.004 | 71 | 35.5 | 57.75 |
| 631 | J31 Chronic rhinitis, nasopharyngitis and pharyngitis | 3 | 12.41 | 0.005 | 1,010 | 505.0 | 15.64 |
| 640 | J35.3 Hypertrophy of tonsils with hypertrophy of adenoids | 4 | 12.17 | 0.005 | 775 | 387.5 | 17.68 |
| 662 | J31.0 Chronic rhinitis | 4 | 11.81 | 0.007 | 982 | 491.0 | 15.48 |
| 668 | J04 Acute laryngitis and tracheitis | 3 | 11.63 | 0.008 | 180 | 90.0 | 35.56 |
| 676 | J96.90 Respiratory failure, unspecified, unspecified whether with hypoxia or hypercapnia | 5 | 11.44 | 0.009 | 122 | 61.0 | 42.62 |
| 679 | J38.0 Paralysis of vocal cords and larynx | 4 | 11.36 | 0.009 | 258 | 129.0 | 29.46 |
| 683 | J35.0 Chronic tonsillitis and adenoiditis | 4 | 11.27 | 0.009 | 203 | 101.5 | 33.00 |
| 684 | J45.50 Severe persistent asthma, uncomplicated | 5 | 11.24 | 0.011 | 364 | 182.0 | 24.73 |
| 686 | J04.10 Acute tracheitis without obstruction | 5 | 11.22 | 0.011 | 129 | 64.5 | 41.09 |
| 692 | J81.0 Acute pulmonary edema | 4 | 11.14 | 0.011 | 47 | 23.5 | 65.96 |
| 696 | J01.0 Acute maxillary sinusitis | 4 | 11.06 | 0.014 | 238 | 119.0 | 30.25 |
| 710 | J81.1 Chronic pulmonary edema | 4 | 10.81 | 0.014 | 67 | 33.5 | 55.22 |
| 718 | J04.1 Acute tracheitis | 4 | 10.70 | 0.015 | 130 | 65.0 | 40.00 |
| 740 | J60-J70 Lung diseases due to external agents | 2 | 10.35 | 0.030 | 166 | 83.0 | 34.94 |
| 746 | J91 Pleural effusion in conditions classified elsewhere | 3 | 10.19 | 0.033 | 42 | 21.0 | 66.67 |
| 771 | J45.51 Severe persistent asthma with (acute) exacerbation | 5 | 9.77 | 0.046 | 147 | 73.5 | 36.05 |
| K00-K93 Diseases of the digestive system | | | | | | | |
| 12 | K00-K93 Diseases of the digestive system | 1 | 522.30 | 0.001 | 32,690 | 16,345.0 | 17.83 |
| 45 | K00-K14 Diseases of oral cavity, salivary glands and jaws | 2 | 197.57 | 0.001 | 4,837 | 2,418.5 | 28.39 |
| 54 | K55-K64 Other diseases of intestines | 2 | 176.01 | 0.001 | 12,610 | 6,305.0 | 16.67 |
| 63 | K59 Other functional intestinal disorders | 3 | 152.00 | 0.001 | 10,839 | 5,419.5 | 16.71 |
| 66 | K59.0 Constipation | 4 | 148.08 | 0.001 | 10,604 | 5,302.0 | 16.67 |
| 107 | K59.00 Constipation, unspecified | 5 | 98.53 | 0.001 | 6,387 | 3,193.5 | 17.52 |
| 157 | K90-K95 Other diseases of the digestive system | 2 | 66.79 | 0.001 | 2,295 | 1,147.5 | 24.01 |
| 159 | K02 Dental caries | 3 | 65.44 | 0.001 | 1,196 | 598.0 | 32.78 |
| 164 | K94 Complications of artificial openings of the digestive system | 3 | 62.32 | 0.001 | 787 | 393.5 | 39.26 |
| 168 | K02.9 Dental caries, unspecified | 4 | 61.35 | 0.001 | 1,179 | 589.5 | 31.98 |
| 192 | K12 Stomatitis and related lesions | 3 | 53.07 | 0.001 | 621 | 310.5 | 40.74 |
| 195 | K94.2 Gastrostomy complications | 4 | 52.95 | 0.001 | 681 | 340.5 | 38.91 |
| 215 | K52.9 Noninfective gastroenteritis and colitis, unspecified | 4 | 47.91 | 0.001 | 2,542 | 1,271.0 | 19.35 |
| 216 | K12.3 Oral mucositis (ulcerative) | 4 | 47.03 | 0.001 | 302 | 151.0 | 54.30 |
| 230 | K52 Other and unspecified noninfective gastroenteritis and colitis | 3 | 44.15 | 0.001 | 2,995 | 1,497.5 | 17.13 |
| 262 | K05 Gingivitis and periodontal diseases | 3 | 36.79 | 0.001 | 339 | 169.5 | 45.72 |
| 267 | K20-K31 Diseases of oesophagus, stomach and duodenum | 2 | 36.43 | 0.001 | 4,871 | 2,435.5 | 12.22 |
| 280 | K05.1 Chronic gingivitis | 4 | 33.82 | 0.001 | 318 | 159.0 | 45.28 |
| 281 | K05.10 Chronic gingivitis, plaque induced | 5 | 33.82 | 0.001 | 318 | 159.0 | 45.28 |
| 284 | K80-K87 Disorders of gallbladder, biliary tract and pancreas | 2 | 33.46 | 0.001 | 923 | 461.5 | 26.76 |
| 298 | K12.30 Oral mucositis (ulcerative), unspecified | 5 | 31.57 | 0.001 | 213 | 106.5 | 53.05 |
| 303 | K50-K52 Noninfective enteritis and colitis | 2 | 31.33 | 0.001 | 4,961 | 2,480.5 | 11.23 |
| 312 | K59.09 Other constipation | 5 | 30.65 | 0.001 | 2,043 | 1,021.5 | 17.28 |
| 319 | K70-K77 Diseases of liver | 2 | 30.12 | 0.001 | 858 | 429.0 | 26.34 |
| 344 | K29 Gastritis and duodenitis | 3 | 28.07 | 0.001 | 968 | 484.0 | 23.97 |
| 350 | K92 Other diseases of digestive system | 3 | 27.40 | 0.001 | 696 | 348.0 | 27.87 |
| 405 | K59.03 Drug induced constipation | 5 | 22.58 | 0.001 | 191 | 95.5 | 47.64 |
| 433 | K00 Disorders of tooth development and eruption | 3 | 21.27 | 0.001 | 901 | 450.5 | 21.64 |
| 469 | K04 Diseases of pulp and periapical tissues | 3 | 19.41 | 0.001 | 259 | 129.5 | 38.22 |
| 491 | K94.23 Gastrostomy malfunction | 5 | 18.20 | 0.001 | 270 | 135.0 | 36.30 |
| 513 | K04.7 Periapical abscess without sinus | 4 | 17.13 | 0.001 | 237 | 118.5 | 37.55 |
| 514 | K94.29 Other complications of gastrostomy | 5 | 17.11 | 0.001 | 212 | 106.0 | 39.62 |
| 544 | K56 Paralytic ileus and intestinal obstruction without hernia | 3 | 15.33 | 0.001 | 407 | 203.5 | 27.27 |
| 546 | K65-K69 Diseases of peritoneum | 2 | 15.27 | 0.001 | 127 | 63.5 | 48.03 |
| 571 | K00.7 Teething syndrome | 4 | 14.35 | 0.001 | 799 | 399.5 | 18.90 |
| 582 | K12.31 Oral mucositis (ulcerative) due to antineoplastic therapy | 5 | 13.99 | 0.002 | 74 | 37.0 | 59.46 |
| 612 | K22 Other diseases of esophagus | 3 | 13.17 | 0.003 | 160 | 80.0 | 40.00 |
| 670 | K94.22 Gastrostomy infection | 5 | 11.60 | 0.008 | 63 | 31.5 | 58.73 |
| 694 | K76 Other diseases of liver | 3 | 11.09 | 0.011 | 402 | 201.0 | 23.38 |
| 701 | K29.5 Unspecified chronic gastritis | 4 | 10.99 | 0.014 | 214 | 107.0 | 31.78 |
| 709 | K92.2 Gastrointestinal hemorrhage, unspecified | 4 | 10.82 | 0.014 | 40 | 20.0 | 70.00 |
| 724 | K29.50 Unspecified chronic gastritis without bleeding | 5 | 10.55 | 0.017 | 210 | 105.0 | 31.43 |
| 726 | K13 Other diseases of lip and oral mucosa | 3 | 10.54 | 0.017 | 567 | 283.5 | 19.22 |
| 750 | K86 Other diseases of pancreas | 3 | 10.11 | 0.034 | 320 | 160.0 | 25.00 |
| 753 | K31 Other diseases of stomach and duodenum | 3 | 10.06 | 0.034 | 562 | 281.0 | 18.86 |
| 768 | K65 Peritonitis | 3 | 9.86 | 0.045 | 62 | 31.0 | 54.84 |
| L00-L99 Diseases of the skin and subcutaneous tissue | | | | | | | |
| 31 | L00-L99 Diseases of the skin and subcutaneous tissue | 1 | 278.48 | 0.001 | 23,130 | 11,565.0 | 15.49 |
| 79 | L20-L30 Dermatitis and eczema | 2 | 130.67 | 0.001 | 11,377 | 5,688.5 | 15.13 |
| 154 | L22 Diaper dermatitis | 3 | 67.86 | 0.001 | 1,887 | 943.5 | 26.66 |
| 205 | L00-L08 Infections of the skin and subcutaneous tissue | 2 | 51.06 | 0.001 | 3,277 | 1,638.5 | 17.61 |
| 208 | L60-L75 Disorders of skin appendages | 2 | 50.50 | 0.001 | 2,552 | 1,276.0 | 19.83 |
| 242 | L03 Cellulitis and acute lymphangitis | 3 | 41.18 | 0.001 | 1,320 | 660.0 | 24.85 |
| 278 | L80-L99 Other disorders of the skin and subcutaneous tissue | 2 | 34.01 | 0.001 | 4,033 | 2,016.5 | 12.97 |
| 315 | L20 Atopic dermatitis | 3 | 30.33 | 0.001 | 4,383 | 2,191.5 | 11.75 |
| 346 | L20.8 Other atopic dermatitis | 4 | 27.93 | 0.001 | 3,602 | 1,801.0 | 12.44 |
| 352 | L30 Other and unspecified dermatitis | 3 | 27.25 | 0.001 | 2,440 | 1,220.0 | 14.92 |
| 410 | L30.9 Dermatitis, unspecified | 4 | 22.39 | 0.001 | 1,951 | 975.5 | 15.12 |
| 471 | L20.83 Infantile (acute) (chronic) eczema | 5 | 19.19 | 0.001 | 1,421 | 710.5 | 16.40 |
| 480 | L65 Other nonscarring hair loss | 3 | 18.79 | 0.001 | 311 | 155.5 | 34.41 |
| 535 | L85 Other epidermal thickening | 3 | 15.59 | 0.001 | 932 | 466.0 | 18.24 |
| 548 | L02 Cutaneous abscess, furuncle and carbuncle | 3 | 15.14 | 0.001 | 748 | 374.0 | 20.05 |
| 584 | L49-L54 Urticaria and erythema | 2 | 13.90 | 0.002 | 1,472 | 736.0 | 13.72 |
| 609 | L03.31 Cellulitis of trunk | 5 | 13.23 | 0.003 | 195 | 97.5 | 36.41 |
| 610 | L03.3 Cellulitis and acute lymphangitis of trunk | 4 | 13.23 | 0.003 | 195 | 97.5 | 36.41 |
| 634 | L85.3 Xerosis cutis | 4 | 12.39 | 0.005 | 595 | 297.5 | 20.34 |
| 638 | L65.9 Nonscarring hair loss, unspecified | 4 | 12.27 | 0.005 | 252 | 126.0 | 30.95 |
| 645 | L73.9 Follicular disorder, unspecified | 4 | 12.04 | 0.005 | 196 | 98.0 | 34.69 |
| 738 | L73 Other follicular disorders | 3 | 10.36 | 0.030 | 328 | 164.0 | 25.00 |
| 745 | L03.1 Cellulitis and acute lymphangitis of other parts of limb | 4 | 10.20 | 0.033 | 325 | 162.5 | 24.92 |
| 749 | L53 Other erythematous conditions | 3 | 10.11 | 0.034 | 153 | 76.5 | 35.95 |
| 757 | L03.11 Cellulitis of other parts of limb | 5 | 9.98 | 0.034 | 324 | 162.0 | 24.69 |
| M00-M99 Diseases of the musculoskeletal system and connective tissue | | | | | | | |
| 50 | M00-M99 Diseases of the musculoskeletal system and connective tissue | 1 | 187.33 | 0.001 | 37,918 | 18,959.0 | 9.93 |
| 136 | M70-M79 Other soft tissue disorders | 2 | 75.70 | 0.001 | 5,507 | 2,753.5 | 16.54 |
| 141 | M79 Other and unspecified soft tissue disorders, not elsewhere classified | 3 | 74.35 | 0.001 | 5,022 | 2,511.0 | 17.16 |
| 162 | M79.6 Pain in limb, hand, foot, fingers and toes | 4 | 63.07 | 0.001 | 3,719 | 1,859.5 | 18.37 |
| 193 | M54 Dorsalgia | 3 | 53.05 | 0.001 | 2,586 | 1,293.0 | 20.19 |
| 209 | M50-M54 Other dorsopathies | 2 | 50.38 | 0.001 | 2,826 | 1,413.0 | 18.83 |
| 279 | M60-M63 Disorders of muscles | 2 | 34.00 | 0.001 | 5,306 | 2,653.0 | 11.31 |
| 289 | M62 Other disorders of muscle | 3 | 33.08 | 0.001 | 5,219 | 2,609.5 | 11.25 |
| 299 | M79.67 Pain in foot and toes | 5 | 31.45 | 0.001 | 1,161 | 580.5 | 23.17 |
| 339 | M62.8 Other specified disorders of muscle | 4 | 28.59 | 0.001 | 4,867 | 2,433.5 | 10.83 |
| 369 | M25.5 Pain in joint | 4 | 25.06 | 0.001 | 5,935 | 2,967.5 | 9.18 |
| 376 | M20-M25 Other joint disorders | 2 | 24.88 | 0.001 | 13,786 | 6,893.0 | 6.01 |
| 378 | M62.89 Other specified disorders of muscle | 5 | 24.71 | 0.001 | 1,928 | 964.0 | 15.98 |
| 428 | M99.0 Segmental and somatic dysfunction | 4 | 21.42 | 0.001 | 172 | 86.0 | 48.84 |
| 442 | M99-M99 Biomechanical lesions, not elsewhere classified | 2 | 20.89 | 0.001 | 176 | 88.0 | 47.73 |
| 443 | M99 Biomechanical lesions, not elsewhere classified | 3 | 20.89 | 0.001 | 176 | 88.0 | 47.73 |
| 444 | M79.60 Pain in limb, unspecified | 5 | 20.84 | 0.001 | 1,641 | 820.5 | 15.90 |
| 487 | M54.5 Low back pain | 4 | 18.36 | 0.001 | 1,162 | 581.0 | 17.73 |
| 488 | M54.9 Dorsalgia, unspecified | 4 | 18.35 | 0.001 | 510 | 255.0 | 26.67 |
| 498 | M54.50 Low back pain, unspecified | 5 | 17.74 | 0.001 | 811 | 405.5 | 20.84 |
| 557 | M25.57 Pain in ankle and joints of foot | 5 | 14.73 | 0.001 | 940 | 470.0 | 17.66 |
| 558 | M79.10 Myalgia, unspecified site | 5 | 14.73 | 0.001 | 358 | 179.0 | 28.49 |
| 603 | M24 Other specific joint derangements | 3 | 13.31 | 0.003 | 1,276 | 638.0 | 14.42 |
| 639 | M87.1 Osteonecrosis due to drugs | 4 | 12.27 | 0.005 | 46 | 23.0 | 69.57 |
| 641 | M79.1 Myalgia | 4 | 12.13 | 0.005 | 744 | 372.0 | 18.01 |
| 647 | M21.85 Other specified acquired deformities of thigh | 5 | 12.01 | 0.006 | 191 | 95.5 | 35.08 |
| 648 | M21.4 Flat foot [pes planus] (acquired) | 4 | 12.00 | 0.006 | 1,057 | 528.5 | 15.04 |
| 651 | M79.671 Pain in right foot | 6 | 11.91 | 0.007 | 512 | 256.0 | 21.48 |
| 677 | M79.672 Pain in left foot | 6 | 11.40 | 0.009 | 478 | 239.0 | 21.76 |
| 725 | M21 Other acquired deformities of limbs | 3 | 10.55 | 0.017 | 3,110 | 1,555.0 | 8.23 |
| 743 | M25.571 Pain in right ankle and joints of right foot | 6 | 10.24 | 0.031 | 463 | 231.5 | 20.95 |
| 765 | M62.83 Muscle spasm | 5 | 9.89 | 0.043 | 335 | 167.5 | 24.18 |
| N00-N99 Diseases of the genitourinary system | | | | | | | |
| 87 | N00-N99 Diseases of the genitourinary system | 1 | 120.68 | 0.001 | 14,389 | 7,194.5 | 12.93 |
| 247 | N30-N39 Other diseases of urinary system | 2 | 39.23 | 0.001 | 2,950 | 1,475.0 | 16.27 |
| 249 | N17-N19 Renal failure | 2 | 38.76 | 0.001 | 865 | 432.5 | 29.71 |
| 276 | N17 Acute kidney failure | 3 | 34.37 | 0.001 | 432 | 216.0 | 39.35 |
| 306 | N17.9 Acute kidney failure, unspecified | 4 | 31.08 | 0.001 | 397 | 198.5 | 39.04 |
| 462 | N40-N53 Diseases of male genital organs | 2 | 19.68 | 0.001 | 2,386 | 1,193.0 | 12.82 |
| 490 | N30 Cystitis | 3 | 18.22 | 0.001 | 470 | 235.0 | 27.66 |
| 499 | N39 Other disorders of urinary system | 3 | 17.65 | 0.001 | 1,965 | 982.5 | 13.38 |
| 522 | N25-N29 Other disorders of kidney and ureter | 2 | 16.14 | 0.001 | 1,257 | 628.5 | 15.99 |
| 524 | N30.0 Acute cystitis | 4 | 16.06 | 0.001 | 382 | 191.0 | 28.80 |
| 555 | N80-N98 Noninflammatory disorders of female genital tract | 2 | 14.88 | 0.001 | 3,425 | 1,712.5 | 9.31 |
| 578 | N30.00 Acute cystitis without hematuria | 5 | 14.10 | 0.002 | 261 | 130.5 | 32.57 |
| 605 | N47 Disorders of prepuce | 3 | 13.26 | 0.003 | 922 | 461.0 | 16.92 |
| 616 | N28.8 Other specified disorders of kidney and ureter | 4 | 13.01 | 0.003 | 674 | 337.0 | 19.58 |
| 630 | N10-N16 Renal tubulo-interstitial diseases | 2 | 12.49 | 0.004 | 1,889 | 944.5 | 11.49 |
| 654 | N25 Disorders resulting from impaired renal tubular function | 3 | 11.89 | 0.007 | 210 | 105.0 | 33.33 |
| 661 | N28.89 Other specified disorders of kidney and ureter | 5 | 11.82 | 0.007 | 583 | 291.5 | 20.07 |
| 698 | N39.0 Urinary tract infection, site not specified | 4 | 11.02 | 0.014 | 855 | 427.5 | 16.02 |
| P00-P96 Certain conditions originating in the perinatal period | | | | | | | |
| 285 | P07 Disorders of newborn related to short gestation and low birth weight, not elsewhere classified | 3 | 33.42 | 0.001 | 2,908 | 1,454.0 | 15.13 |
| 290 | P05-P08 Disorders related to length of gestation and fetal growth | 2 | 33.03 | 0.001 | 3,117 | 1,558.5 | 14.53 |
| 314 | P00-P96 Certain conditions originating in the perinatal period | 1 | 30.46 | 0.001 | 5,048 | 2,524.0 | 10.97 |
| 519 | P07.3 Preterm [premature] newborn [other] | 4 | 16.29 | 0.001 | 1,759 | 879.5 | 13.59 |
| 635 | P07.2 Extreme immaturity of newborn | 4 | 12.37 | 0.005 | 433 | 216.5 | 23.79 |
| 716 | P91.6 Hypoxic ischemic encephalopathy [HIE] | 4 | 10.74 | 0.014 | 332 | 166.0 | 25.30 |
| 735 | P91 Other disturbances of cerebral status of newborn | 3 | 10.41 | 0.021 | 419 | 209.5 | 22.20 |
| Q00-Q99 Congenital malformations, deformations and chromosomal abnormalities | | | | | | | |
| 153 | Q00-Q99 Congenital malformations, deformations and chromosomal abnormalities | 1 | 68.05 | 0.001 | 14,672 | 7,336.0 | 9.62 |
| 331 | Q02 | 3 | 29.12 | 0.001 | 433 | 216.5 | 36.26 |
| 351 | Q20-Q28 Congenital malformations of the circulatory system | 2 | 27.38 | 0.001 | 2,083 | 1,041.5 | 16.18 |
| 419 | Q00-Q07 Congenital malformations of the nervous system | 2 | 21.83 | 0.001 | 1,027 | 513.5 | 20.55 |
| 538 | Q80-Q89 Other congenital malformations | 2 | 15.45 | 0.001 | 1,964 | 982.0 | 12.53 |
| 549 | Q87.89 Other specified congenital malformation syndromes, not elsewhere classified | 5 | 15.01 | 0.001 | 438 | 219.0 | 26.03 |
| 575 | Q90-Q99 Chromosomal abnormalities, not elsewhere classified | 2 | 14.18 | 0.002 | 1,000 | 500.0 | 16.80 |
| 586 | Q87.8 Other specified congenital malformation syndromes, not elsewhere classified | 4 | 13.90 | 0.002 | 456 | 228.0 | 24.56 |
| 598 | Q25 Congenital malformations of great arteries | 3 | 13.40 | 0.003 | 778 | 389.0 | 18.51 |
| 622 | Q65-Q79 Congenital malformations and deformations of the musculoskeletal system | 2 | 12.85 | 0.004 | 4,240 | 2,120.0 | 7.78 |
| 736 | Q93 Monosomies and deletions from the autosomes, not elsewhere classified | 3 | 10.41 | 0.021 | 617 | 308.5 | 18.31 |
| 741 | Q87 Other specified congenital malformation syndromes affecting multiple systems | 3 | 10.28 | 0.030 | 800 | 400.0 | 16.00 |
| R00-R99 Symptoms, signs and abnormal clinical and laboratory findings, not elsewhere classified | | | | | | | |
| 1 | R00-R99 Symptoms, signs and abnormal clinical and laboratory findings, not elsewhere classified | 1 | 2,822.93 | 0.001 | 130,582 | 65,291.0 | 20.72 |
| 7 | R00-R09 Symptoms and signs involving the circulatory and respiratory systems | 2 | 1,126.95 | 0.001 | 33,100 | 16,550.0 | 25.95 |
| 9 | R50-R69 General symptoms and signs | 2 | 835.14 | 0.001 | 36,081 | 18,040.5 | 21.43 |
| 13 | R10-R19 Symptoms and signs involving the digestive system and abdomen | 2 | 476.24 | 0.001 | 27,832 | 13,916.0 | 18.45 |
| 19 | R50 Fever of other and unknown origin | 3 | 350.58 | 0.001 | 7,987 | 3,993.5 | 29.41 |
| 22 | R06 Abnormalities of breathing | 3 | 332.80 | 0.001 | 9,751 | 4,875.5 | 25.98 |
| 29 | R06.0 Dyspnea | 4 | 282.38 | 0.001 | 2,305 | 1,152.5 | 48.46 |
| 32 | R50.9 Fever, unspecified | 4 | 269.36 | 0.001 | 7,127 | 3,563.5 | 27.32 |
| 33 | R05 Cough | 3 | 266.84 | 0.001 | 9,907 | 4,953.5 | 23.10 |
| 34 | R09 Other symptoms and signs involving the circulatory and respiratory system | 3 | 245.58 | 0.001 | 7,179 | 3,589.5 | 26.01 |
| 38 | R11 Nausea and vomiting | 3 | 227.84 | 0.001 | 8,567 | 4,283.5 | 22.96 |
| 52 | R07 Pain in throat and chest | 3 | 178.94 | 0.001 | 2,582 | 1,291.0 | 36.79 |
| 55 | R05.9 Cough, unspecified | 4 | 175.41 | 0.001 | 5,758 | 2,879.0 | 24.56 |
| 57 | R11.1 Vomiting | 4 | 165.85 | 0.001 | 5,973 | 2,986.5 | 23.46 |
| 60 | R09.8 Other specified symptoms and signs involving the circulatory and respiratory systems | 4 | 159.41 | 0.001 | 5,992 | 2,996.0 | 22.96 |
| 62 | R11.10 Vomiting, unspecified | 5 | 157.50 | 0.001 | 5,254 | 2,627.0 | 24.36 |
| 77 | R62 Lack of expected normal physiological development in childhood and adults | 3 | 132.16 | 0.001 | 8,716 | 4,358.0 | 17.37 |
| 80 | R09.81 Nasal congestion | 5 | 129.61 | 0.001 | 4,952 | 2,476.0 | 22.78 |
| 83 | R62.5 Other and unspecified lack of expected normal physiological development in childhood | 4 | 127.90 | 0.001 | 8,104 | 4,052.0 | 17.72 |
| 85 | R00 Abnormalities of heart beat | 3 | 125.04 | 0.001 | 1,742 | 871.0 | 37.43 |
| 91 | R13 Aphagia and dysphagia | 3 | 113.98 | 0.001 | 4,674 | 2,337.0 | 21.99 |
| 92 | R13.1 Dysphagia | 4 | 113.98 | 0.001 | 4,674 | 2,337.0 | 21.99 |
| 94 | R62.50 Unspecified lack of expected normal physiological development in childhood | 5 | 108.77 | 0.001 | 4,326 | 2,163.0 | 22.33 |
| 97 | R20-R23 Symptoms and signs involving the skin and subcutaneous tissue | 2 | 106.69 | 0.001 | 5,017 | 2,508.5 | 20.55 |
| 98 | R09.0 Asphyxia and hypoxemia | 4 | 106.03 | 0.001 | 1,168 | 584.0 | 41.95 |
| 99 | R09.02 Hypoxemia | 5 | 106.03 | 0.001 | 1,168 | 584.0 | 41.95 |
| 101 | R40-R46 Symptoms and signs involving cognition, perception, emotional state and behaviour | 2 | 105.04 | 0.001 | 9,087 | 4,543.5 | 15.18 |
| 105 | R06.02 Shortness of breath | 5 | 102.27 | 0.001 | 786 | 393.0 | 49.87 |
| 109 | R06.8 Other abnormalities of breathing | 4 | 97.59 | 0.001 | 5,187 | 2,593.5 | 19.34 |
| 112 | R50.81 Fever presenting with conditions classified elsewhere | 5 | 95.83 | 0.001 | 778 | 389.0 | 48.59 |
| 115 | R50.8 Other specified fever | 4 | 95.15 | 0.001 | 848 | 424.0 | 46.46 |
| 119 | R63 | 3 | 89.77 | 0.001 | 6,371 | 3,185.5 | 16.75 |
| 121 | R07.9 Chest pain, unspecified | 4 | 88.32 | 0.001 | 1,307 | 653.5 | 36.34 |
| 122 | R90-R96 Abnormal findings on diagnostic imaging and in function studies, without diagnosis | 2 | 86.27 | 0.001 | 3,888 | 1,944.0 | 20.99 |
| 125 | R53.8 Other malaise and fatigue | 4 | 84.14 | 0.001 | 2,432 | 1,216.0 | 26.15 |
| 127 | R53 Malaise and fatigue | 3 | 82.67 | 0.001 | 3,085 | 1,542.5 | 23.05 |
| 128 | R19 Other symptoms and signs involving the digestive system and abdomen | 3 | 82.20 | 0.001 | 3,940 | 1,970.0 | 20.36 |
| 129 | R53.83 Other fatigue | 5 | 79.80 | 0.001 | 1,767 | 883.5 | 29.82 |
| 130 | R07.8 Other chest pain | 4 | 79.60 | 0.001 | 992 | 496.0 | 39.52 |
| 134 | R63.3 | 4 | 76.03 | 0.001 | 5,762 | 2,881.0 | 16.21 |
| 135 | R19.7 Diarrhea, unspecified | 4 | 75.84 | 0.001 | 2,880 | 1,440.0 | 22.85 |
| 138 | R06.00 Dyspnea, unspecified | 5 | 74.92 | 0.001 | 545 | 272.5 | 51.19 |
| 140 | R06.89 Other abnormalities of breathing | 5 | 74.42 | 0.001 | 1,984 | 992.0 | 27.22 |
| 142 | R70-R79 Abnormal findings on examination of blood, without diagnosis | 2 | 73.91 | 0.001 | 3,830 | 1,915.0 | 19.58 |
| 144 | R00.0 Tachycardia, unspecified | 4 | 70.93 | 0.001 | 863 | 431.5 | 39.98 |
| 149 | R06.03 Acute respiratory distress | 5 | 69.44 | 0.001 | 770 | 385.0 | 41.82 |
| 150 | R07.89 Other chest pain | 5 | 69.24 | 0.001 | 863 | 431.5 | 39.51 |
| 151 | R21 Rash and other nonspecific skin eruption | 3 | 69.11 | 0.001 | 2,840 | 1,420.0 | 21.97 |
| 166 | R10 Abdominal and pelvic pain | 3 | 61.57 | 0.001 | 9,110 | 4,555.0 | 11.61 |
| 174 | R51.9 Headache, unspecified | 4 | 59.70 | 0.001 | 2,453 | 1,226.5 | 21.97 |
| 181 | R51 Headache | 3 | 57.52 | 0.001 | 2,697 | 1,348.5 | 20.58 |
| 197 | R94.3 Abnormal results of cardiovascular function studies | 4 | 52.50 | 0.001 | 927 | 463.5 | 33.33 |
| 199 | R94.31 Abnormal electrocardiogram [ECG] [EKG] | 5 | 52.45 | 0.001 | 922 | 461.0 | 33.41 |
| 206 | R68 Other general symptoms and signs | 3 | 50.68 | 0.001 | 2,788 | 1,394.0 | 19.01 |
| 217 | R63.30 Feeding difficulties, unspecified | 5 | 46.77 | 0.001 | 2,996 | 1,498.0 | 17.62 |
| 219 | R30-R39 Symptoms and signs involving the urinary system | 2 | 46.01 | 0.001 | 4,522 | 2,261.0 | 14.24 |
| 228 | R11.2 Nausea with vomiting, unspecified | 4 | 44.48 | 0.001 | 1,529 | 764.5 | 24.00 |
| 229 | R13.12 Dysphagia, oropharyngeal phase | 5 | 44.20 | 0.001 | 1,657 | 828.5 | 22.99 |
| 233 | R13.10 Dysphagia, unspecified | 5 | 43.57 | 0.001 | 1,778 | 889.0 | 22.05 |
| 235 | R94 Abnormal results of function studies | 3 | 43.23 | 0.001 | 2,223 | 1,111.5 | 19.66 |
| 244 | R63.39 Other feeding difficulties | 5 | 40.82 | 0.001 | 1,820 | 910.0 | 21.10 |
| 252 | R79 Other abnormal findings of blood chemistry | 3 | 38.50 | 0.001 | 2,101 | 1,050.5 | 19.09 |
| 254 | R06.09 Other forms of dyspnea | 5 | 38.08 | 0.001 | 199 | 99.5 | 59.80 |
| 264 | R06.2 Wheezing | 4 | 36.55 | 0.001 | 1,493 | 746.5 | 22.04 |
| 282 | R00.2 Palpitations | 4 | 33.62 | 0.001 | 504 | 252.0 | 36.11 |
| 295 | R41 Other symptoms and signs involving cognitive functions and awareness | 3 | 31.85 | 0.001 | 1,433 | 716.5 | 21.00 |
| 311 | R91 Abnormal findings on diagnostic imaging of lung | 3 | 30.81 | 0.001 | 463 | 231.5 | 36.07 |
| 322 | R91.8 Other nonspecific abnormal finding of lung field | 4 | 29.64 | 0.001 | 410 | 205.0 | 37.56 |
| 324 | R25-R29 Symptoms and signs involving the nervous and musculoskeletal systems | 2 | 29.54 | 0.001 | 4,091 | 2,045.5 | 12.00 |
| 332 | R68.8 Other general symptoms and signs | 4 | 29.09 | 0.001 | 2,164 | 1,082.0 | 16.36 |
| 340 | R79.8 Other specified abnormal findings of blood chemistry | 4 | 28.55 | 0.001 | 1,725 | 862.5 | 18.14 |
| 349 | R56 Convulsions, not elsewhere classified | 3 | 27.49 | 0.001 | 1,405 | 702.5 | 19.72 |
| 358 | R45 Symptoms and signs involving emotional state | 3 | 26.53 | 0.001 | 2,702 | 1,351.0 | 13.99 |
| 364 | R41.8 Other symptoms and signs involving cognitive functions and awareness | 4 | 26.03 | 0.001 | 1,219 | 609.5 | 20.59 |
| 366 | R65.2 Severe sepsis | 4 | 25.93 | 0.001 | 194 | 97.0 | 50.52 |
| 384 | R68.89 Other general symptoms and signs | 5 | 24.17 | 0.001 | 2,022 | 1,011.0 | 15.43 |
| 385 | R65 Symptoms and signs specifically associated with systemic inflammation and infection | 3 | 24.06 | 0.001 | 216 | 108.0 | 46.30 |
| 387 | R10.9 Unspecified abdominal pain | 4 | 24.00 | 0.001 | 2,332 | 1,166.0 | 14.32 |
| 390 | R65.21 Severe sepsis with septic shock | 5 | 23.72 | 0.001 | 175 | 87.5 | 50.86 |
| 393 | R62.51 Failure to thrive (child) | 5 | 23.35 | 0.001 | 2,469 | 1,234.5 | 13.73 |
| 401 | R80-R82 Abnormal findings on examination of urine, without diagnosis | 2 | 22.76 | 0.001 | 922 | 461.0 | 22.13 |
| 403 | R78.8 | 4 | 22.63 | 0.001 | 233 | 116.5 | 43.35 |
| 404 | R78.81 Bacteremia | 5 | 22.63 | 0.001 | 233 | 116.5 | 43.35 |
| 425 | R43 Disturbances of smell and taste | 3 | 21.54 | 0.001 | 183 | 91.5 | 47.54 |
| 435 | R20 Disturbances of skin sensation | 3 | 21.20 | 0.001 | 552 | 276.0 | 27.54 |
| 437 | R68.1 Nonspecific symptoms peculiar to infancy | 4 | 21.14 | 0.001 | 568 | 284.0 | 27.11 |
| 439 | R06.83 Snoring | 5 | 21.03 | 0.001 | 2,820 | 1,410.0 | 12.20 |
| 450 | R68.12 Fussy infant (baby) | 5 | 20.45 | 0.001 | 486 | 243.0 | 28.81 |
| 457 | R59 Enlarged lymph nodes | 3 | 20.03 | 0.001 | 728 | 364.0 | 23.35 |
| 463 | R10.3 Pain localized to other parts of lower abdomen | 4 | 19.63 | 0.001 | 2,486 | 1,243.0 | 12.55 |
| 464 | R09.89 Other specified symptoms and signs involving the circulatory and respiratory systems | 5 | 19.63 | 0.001 | 832 | 416.0 | 21.63 |
| 466 | R45.8 Other symptoms and signs involving emotional state | 4 | 19.47 | 0.001 | 2,256 | 1,128.0 | 13.12 |
| 467 | R41.82 Altered mental status, unspecified | 5 | 19.44 | 0.001 | 229 | 114.5 | 40.61 |
| 470 | R79.89 Other specified abnormal findings of blood chemistry | 5 | 19.36 | 0.001 | 1,544 | 772.0 | 15.80 |
| 473 | R52 Pain, unspecified | 3 | 19.17 | 0.001 | 525 | 262.5 | 26.86 |
| 483 | R11.0 Nausea | 4 | 18.70 | 0.001 | 1,065 | 532.5 | 18.69 |
| 485 | R00.1 Bradycardia, unspecified | 4 | 18.57 | 0.001 | 344 | 172.0 | 32.56 |
| 500 | R42 Dizziness and giddiness | 3 | 17.58 | 0.001 | 1,201 | 600.5 | 17.07 |
| 516 | R47.89 Other speech disturbances | 5 | 16.96 | 0.001 | 818 | 409.0 | 20.29 |
| 520 | R47-R49 Symptoms and signs involving speech and voice | 2 | 16.26 | 0.001 | 1,866 | 933.0 | 13.18 |
| 521 | R18 Ascites | 3 | 16.18 | 0.001 | 99 | 49.5 | 55.56 |
| 527 | R47.8 Other speech disturbances | 4 | 15.97 | 0.001 | 837 | 418.5 | 19.47 |
| 532 | R18.8 Other ascites | 4 | 15.74 | 0.001 | 98 | 49.0 | 55.10 |
| 533 | R47 Speech disturbances, not elsewhere classified | 3 | 15.70 | 0.001 | 1,241 | 620.5 | 15.87 |
| 536 | R06.82 Tachypnea, not elsewhere classified | 5 | 15.56 | 0.001 | 232 | 116.0 | 36.21 |
| 542 | R29 Other symptoms and signs involving the nervous and musculoskeletal systems | 3 | 15.39 | 0.001 | 2,480 | 1,240.0 | 11.13 |
| 543 | R60 Edema, not elsewhere classified | 3 | 15.39 | 0.001 | 229 | 114.5 | 36.24 |
| 550 | R05.1 Acute cough | 4 | 14.99 | 0.001 | 394 | 197.0 | 27.41 |
| 561 | R43.9 Unspecified disturbances of smell and taste | 4 | 14.63 | 0.001 | 36 | 18.0 | 83.33 |
| 566 | R05.3 Chronic cough | 4 | 14.47 | 0.001 | 536 | 268.0 | 23.13 |
| 569 | R73 Elevated blood glucose level | 3 | 14.43 | 0.001 | 1,030 | 515.0 | 16.70 |
| 572 | R56.9 Unspecified convulsions | 4 | 14.33 | 0.001 | 876 | 438.0 | 18.04 |
| 574 | R40 Somnolence, stupor and coma | 3 | 14.21 | 0.002 | 618 | 309.0 | 21.36 |
| 576 | R32 Unspecified urinary incontinence | 3 | 14.17 | 0.002 | 716 | 358.0 | 19.83 |
| 577 | R93 Abnormal findings on diagnostic imaging of other body structures | 3 | 14.13 | 0.002 | 833 | 416.5 | 18.37 |
| 590 | R20.0 Anesthesia of skin | 4 | 13.65 | 0.003 | 169 | 84.5 | 39.64 |
| 599 | R56.0 Febrile convulsions | 4 | 13.40 | 0.003 | 524 | 262.0 | 22.52 |
| 600 | R63.0 Anorexia | 4 | 13.36 | 0.003 | 508 | 254.0 | 22.83 |
| 613 | R60.9 Edema, unspecified | 4 | 13.05 | 0.003 | 108 | 54.0 | 48.15 |
| 624 | R45.89 Other symptoms and signs involving emotional state | 5 | 12.78 | 0.004 | 577 | 288.5 | 20.97 |
| 626 | R04 Hemorrhage from respiratory passages | 3 | 12.65 | 0.004 | 966 | 483.0 | 16.15 |
| 627 | R16 Hepatomegaly and splenomegaly, not elsewhere classified | 3 | 12.65 | 0.004 | 251 | 125.5 | 31.47 |
| 646 | R79.82 Elevated C-reactive protein (CRP) | 5 | 12.04 | 0.005 | 139 | 69.5 | 41.01 |
| 652 | R35 Polyuria | 3 | 11.91 | 0.007 | 692 | 346.0 | 18.50 |
| 653 | R82 Other and unspecified abnormal findings in urine | 3 | 11.90 | 0.007 | 650 | 325.0 | 19.08 |
| 657 | R23 Other skin changes | 3 | 11.89 | 0.007 | 782 | 391.0 | 17.39 |
| 660 | R46.8 Other symptoms and signs involving appearance and behavior | 4 | 11.83 | 0.007 | 2,646 | 1,323.0 | 9.45 |
| 663 | R57 Shock, not elsewhere classified | 3 | 11.80 | 0.007 | 123 | 61.5 | 43.09 |
| 667 | R46.89 Other symptoms and signs involving appearance and behavior | 5 | 11.75 | 0.008 | 2,601 | 1,300.5 | 9.50 |
| 669 | R29.8 Other symptoms and signs involving the nervous and musculoskeletal systems | 4 | 11.62 | 0.008 | 1,794 | 897.0 | 11.37 |
| 671 | R46 Symptoms and signs involving appearance and behavior | 3 | 11.57 | 0.008 | 2,662 | 1,331.0 | 9.32 |
| 680 | R70 | 3 | 11.35 | 0.009 | 142 | 71.0 | 39.44 |
| 682 | R09.82 Postnasal drip | 5 | 11.32 | 0.009 | 208 | 104.0 | 32.69 |
| 685 | R79.1 Abnormal coagulation profile | 4 | 11.24 | 0.011 | 186 | 93.0 | 34.41 |
| 691 | R10.31 Right lower quadrant pain | 5 | 11.14 | 0.011 | 577 | 288.5 | 19.58 |
| 697 | R10.8 Other abdominal pain | 4 | 11.06 | 0.014 | 2,587 | 1,293.5 | 9.24 |
| 700 | R13.11 Dysphagia, oral phase | 5 | 11.01 | 0.014 | 431 | 215.5 | 22.51 |
| 703 | R93.89 Abnormal findings on diagnostic imaging of other specified body structures | 5 | 10.94 | 0.014 | 254 | 127.0 | 29.13 |
| 713 | R14.0 Abdominal distension (gaseous) | 4 | 10.77 | 0.014 | 405 | 202.5 | 22.96 |
| 720 | R25 Abnormal involuntary movements | 3 | 10.63 | 0.015 | 750 | 375.0 | 16.80 |
| 728 | R53.81 Other malaise | 5 | 10.52 | 0.018 | 498 | 249.0 | 20.48 |
| 734 | R73.9 Hyperglycemia, unspecified | 4 | 10.43 | 0.021 | 266 | 133.0 | 27.82 |
| 758 | R93.8 Abnormal findings on diagnostic imaging of other specified body structures | 4 | 9.94 | 0.035 | 264 | 132.0 | 27.27 |
| 759 | R10.1 Pain localized to upper abdomen | 4 | 9.94 | 0.036 | 1,526 | 763.0 | 11.40 |
| 767 | R70.0 Elevated erythrocyte sedimentation rate | 4 | 9.88 | 0.044 | 135 | 67.5 | 37.78 |
| S00-T98 Injury, poisoning and certain other consequences of external causes | | | | | | | |
| 56 | T80-T88 Complications of surgical and medical care, not elsewhere classified | 2 | 170.74 | 0.001 | 2,916 | 1,458.0 | 33.88 |
| 69 | S00-T98 Injury, poisoning and certain other consequences of external causes | 1 | 145.62 | 0.001 | 37,509 | 18,754.5 | 8.81 |
| 155 | T82 Complications of cardiac and vascular prosthetic devices, implants and grafts | 3 | 67.79 | 0.001 | 514 | 257.0 | 50.19 |
| 161 | S00-S09 Injuries to the head | 2 | 63.35 | 0.001 | 7,726 | 3,863.0 | 12.79 |
| 178 | T86 Complications of transplanted organs and tissue | 3 | 58.39 | 0.001 | 644 | 322.0 | 41.93 |
| 301 | T82.5 Mechanical complication of other cardiac and vascular devices and implants | 4 | 31.44 | 0.001 | 173 | 86.5 | 58.38 |
| 307 | S09 Other and unspecified injuries of head | 3 | 31.05 | 0.001 | 1,927 | 963.5 | 17.90 |
| 320 | S00 Superficial injury of head | 3 | 30.07 | 0.001 | 1,639 | 819.5 | 19.10 |
| 326 | T80 Complications following infusion, transfusion and therapeutic injection | 3 | 29.41 | 0.001 | 329 | 164.5 | 41.64 |
| 343 | S09.9 Unspecified injury of face and head | 4 | 28.25 | 0.001 | 1,833 | 916.5 | 17.51 |
| 353 | T17 Foreign body in respiratory tract | 3 | 27.10 | 0.001 | 854 | 427.0 | 25.06 |
| 363 | T86.2 Complications of heart transplant | 4 | 26.12 | 0.001 | 189 | 94.5 | 51.32 |
| 370 | T85 Complications of other internal prosthetic devices, implants and grafts | 3 | 25.04 | 0.001 | 677 | 338.5 | 27.03 |
| 374 | S90-S99 Injuries to the ankle and foot | 2 | 24.98 | 0.001 | 3,406 | 1,703.0 | 12.10 |
| 422 | S09.90 Unspecified injury of head | 5 | 21.68 | 0.001 | 1,347 | 673.5 | 17.89 |
| 423 | S09.90X | 6 | 21.68 | 0.001 | 1,347 | 673.5 | 17.89 |
| 436 | T17.9 Foreign body in respiratory tract, part unspecified | 4 | 21.15 | 0.001 | 575 | 287.5 | 26.96 |
| 451 | T82.8 Other specified complications of cardiac and vascular prosthetic devices, implants and grafts | 4 | 20.43 | 0.001 | 214 | 107.0 | 42.99 |
| 452 | T80.2 Infections following infusion, transfusion and therapeutic injection | 4 | 20.29 | 0.001 | 153 | 76.5 | 50.33 |
| 460 | T15-T19 Effects of foreign body entering through natural orifice | 2 | 19.79 | 0.001 | 1,486 | 743.0 | 16.29 |
| 465 | T80.21 Infection due to central venous catheter | 5 | 19.48 | 0.001 | 151 | 75.5 | 49.67 |
| 474 | S09.90XA Unspecified injury of head, initial encounter | 7 | 19.16 | 0.001 | 1,235 | 617.5 | 17.57 |
| 486 | T85.5 Mechanical complication of gastrointestinal prosthetic devices, implants and grafts | 4 | 18.51 | 0.001 | 206 | 103.0 | 41.75 |
| 509 | T20-T32 Burns and corrosions | 2 | 17.22 | 0.001 | 806 | 403.0 | 20.60 |
| 545 | T85.52 Displacement of gastrointestinal prosthetic devices, implants and grafts | 5 | 15.29 | 0.001 | 135 | 67.5 | 46.67 |
| 547 | T80.211 Bloodstream infection due to central venous catheter | 6 | 15.25 | 0.001 | 108 | 54.0 | 51.85 |
| 552 | T85.528A Displacement of other gastrointestinal prosthetic devices, implants and grafts, initial encounter | 7 | 14.90 | 0.001 | 134 | 67.0 | 46.27 |
| 553 | T85.528 Displacement of other gastrointestinal prosthetic devices, implants and grafts | 6 | 14.90 | 0.001 | 134 | 67.0 | 46.27 |
| 579 | T82.51 Breakdown (mechanical) of other cardiac and vascular devices and implants | 5 | 14.07 | 0.002 | 46 | 23.0 | 73.91 |
| 585 | T86.4 Complications of liver transplant | 4 | 13.90 | 0.002 | 102 | 51.0 | 50.98 |
| 593 | T86.20 Unspecified complication of heart transplant | 5 | 13.57 | 0.003 | 76 | 38.0 | 57.89 |
| 632 | T80.211A Bloodstream infection due to central venous catheter, initial encounter | 7 | 12.40 | 0.005 | 101 | 50.5 | 48.51 |
| 644 | T82.89 Other specified complication of cardiac and vascular prosthetic devices, implants and grafts | 5 | 12.05 | 0.005 | 55 | 27.5 | 63.64 |
| 649 | T17.90 Unspecified foreign body in respiratory tract, part unspecified | 5 | 11.92 | 0.007 | 449 | 224.5 | 22.94 |
| 687 | T86.1 Complications of kidney transplant | 4 | 11.21 | 0.011 | 159 | 79.5 | 37.11 |
| 693 | T36-T50 Poisoning by drugs, medicaments and biological substances | 2 | 11.11 | 0.011 | 652 | 326.0 | 18.40 |
| 708 | T51-T65 Toxic effects of substances chiefly nonmedicinal as to source | 2 | 10.82 | 0.014 | 378 | 189.0 | 23.81 |
| 719 | T17.908 Unspecified foreign body in respiratory tract, part unspecified causing other injury | 6 | 10.69 | 0.015 | 391 | 195.5 | 23.27 |
| 744 | T86.21 Heart transplant rejection | 5 | 10.23 | 0.032 | 60 | 30.0 | 56.67 |
| 754 | T82.59 Other mechanical complication of other cardiac and vascular devices and implants | 5 | 10.05 | 0.034 | 87 | 43.5 | 47.13 |
| 755 | S00.8 Superficial injury of other parts of head | 4 | 10.05 | 0.034 | 650 | 325.0 | 17.54 |
| U00-U95 Codes for special purposes | | | | | | | |
| 2 | U00-U49 Provisional assignment of new diseases of uncertain etiology or emergency use | 2 | 2,305.42 | 0.001 | 3,433 | 1,716.5 | 99.36 |
| 3 | U00-U95 Codes for special purposes | 1 | 2,305.42 | 0.001 | 3,433 | 1,716.5 | 99.36 |
| 4 | U07.1 Emergency use of U07.1 \| COVID-19 | 4 | 2,109.85 | 0.001 | 3,088 | 1,544.0 | 99.74 |
| 5 | U07 | 3 | 2,075.03 | 0.001 | 3,099 | 1,549.5 | 99.29 |
| 36 | U09 Post COVID-19 condition | 3 | 231.51 | 0.001 | 334 | 167.0 | 100.00 |
| 37 | U09.9 Post COVID-19 condition, unspecified | 4 | 231.51 | 0.001 | 334 | 167.0 | 100.00 |
| V01-Y98 External causes of morbidity and mortality | | | | | | | |
| 111 | V01-Y98 External causes of morbidity and mortality | 1 | 95.86 | 0.001 | 5,175 | 2,587.5 | 19.19 |
| 152 | V00-V99 Transport accidents | 2 | 68.95 | 0.001 | 4,135 | 2,067.5 | 18.21 |
| 248 | Y40-Y84 Complications of medical and surgical care | 2 | 39.19 | 0.001 | 382 | 191.0 | 44.50 |
| 269 | Y83 Surgical operation and other surgical procedures as the cause of abnormal reaction of the patient, or of later complication, without mention of misadventure at the time of the procedure | 3 | 36.06 | 0.001 | 284 | 142.0 | 49.30 |
| 354 | V72 Bus occupant injured in collision with two- or three-wheeled motor vehicle | 3 | 26.99 | 0.001 | 532 | 266.0 | 31.58 |
| 368 | Y83.8 Other surgical procedures as the cause of abnormal reaction of the patient, or of later complication, without mention of misadventure at the time of the procedure | 4 | 25.08 | 0.001 | 156 | 78.0 | 55.13 |
| 406 | V72.2 Person on outside of bus injured in collision with two- or three-wheeled motor vehicle in nontraffic accident | 4 | 22.56 | 0.001 | 365 | 182.5 | 34.79 |
| 604 | V65 Occupant of heavy transport vehicle injured in collision with railway train or railway vehicle | 3 | 13.29 | 0.003 | 660 | 330.0 | 20.00 |
| Z00-Z99 Factors influencing health status and contact with health services | | | | | | | |
| 23 | Z00-Z99 Factors influencing health status and contact with health services | 1 | 310.96 | 0.001 | 46,922 | 23,461.0 | 11.50 |
| 39 | Z20-Z29 Persons with potential health hazards related to communicable diseases | 2 | 222.84 | 0.001 | 27,639 | 13,819.5 | 12.68 |
| 40 | Z23 | 3 | 218.62 | 0.001 | 27,469 | 13,734.5 | 12.60 |
| 184 | Z99 Dependence on enabling machines and devices, not elsewhere classified | 3 | 56.06 | 0.001 | 1,645 | 822.5 | 25.96 |
| 204 | Z99.8 Dependence on other enabling machines and devices | 4 | 51.45 | 0.001 | 1,121 | 560.5 | 30.06 |
| 239 | Z99.81 Dependence on supplemental oxygen | 5 | 41.90 | 0.001 | 629 | 314.5 | 36.09 |
| 293 | Z77-Z99 Persons with potential health hazards related to family and personal history and certain conditions influencing health status | 2 | 32.40 | 0.001 | 9,112 | 4,556.0 | 8.43 |
| 300 | Z69-Z76 Persons encountering health services in other circumstances | 2 | 31.45 | 0.001 | 3,131 | 1,565.5 | 14.15 |
| 438 | Z91.09 Other allergy status, other than to drugs and biological substances | 5 | 21.11 | 0.001 | 749 | 374.5 | 23.63 |
| 449 | Z91.0 Allergy status, other than to drugs and biological substances | 4 | 20.54 | 0.001 | 787 | 393.5 | 22.74 |
| 504 | Z91 | 3 | 17.48 | 0.001 | 823 | 411.5 | 20.53 |
| 559 | Z93 | 3 | 14.70 | 0.001 | 634 | 317.0 | 21.45 |
| 563 | Z00-Z13 Persons encountering health services for examinations | 2 | 14.53 | 0.001 | 5,078 | 2,539.0 | 7.56 |
| 568 | Z14-Z15 Genetic carrier and genetic susceptibility to disease | 2 | 14.44 | 0.001 | 693 | 346.5 | 20.35 |
| 591 | Z71 | 3 | 13.61 | 0.003 | 482 | 241.0 | 23.65 |
| 592 | Z71.1 Person with feared health complaint in whom no diagnosis is made | 4 | 13.61 | 0.003 | 482 | 241.0 | 23.65 |
| 594 | Z16 Resistance to antimicrobial drugs | 3 | 13.49 | 0.003 | 101 | 50.5 | 50.50 |
| 595 | Z16-Z16 Resistance to antimicrobial drugs | 2 | 13.49 | 0.003 | 101 | 50.5 | 50.50 |
| 633 | Z99.89 Dependence on other enabling machines and devices | 5 | 12.40 | 0.005 | 492 | 246.0 | 22.36 |
| 664 | Z90.49 Acquired absence of other specified parts of digestive tract | 5 | 11.79 | 0.007 | 325 | 162.5 | 26.77 |
| 665 | Z90.4 Acquired absence of other specified parts of digestive tract | 4 | 11.79 | 0.007 | 325 | 162.5 | 26.77 |
| 706 | Z74.0 Reduced mobility | 4 | 10.89 | 0.014 | 1,362 | 681.0 | 12.63 |
| 711 | Z15 Genetic susceptibility to disease | 3 | 10.80 | 0.014 | 648 | 324.0 | 18.21 |
| 714 | Z74.09 Other reduced mobility | 5 | 10.77 | 0.014 | 1,361 | 680.5 | 12.56 |
| 715 | Z74 Problems related to care provider dependency | 3 | 10.77 | 0.014 | 1,393 | 696.5 | 12.42 |
| 723 | Z94 Transplanted organ and tissue status | 3 | 10.56 | 0.017 | 174 | 87.0 | 34.48 |
| 732 | Z93.1 Gastrostomy status | 4 | 10.49 | 0.019 | 529 | 264.5 | 19.85 |
